# Anatomical phenotyping and staging of brain tumors

**DOI:** 10.1101/2021.03.14.21253533

**Authors:** Kevin Akeret, Flavio Vasella, Victor E. Staartjes, Julia Velz, Timothy Müller, Marian Christoph Neidert, Michael Weller, Luca Regli, Carlo Serra, Niklaus Krayenbühl

**Affiliations:** Department of Neurosurgery, Clinical Neuroscience Center, University Hospital Zurich and University of Zurich, Zurich, Switzerland; Department of Neurology, Clinical Neuroscience Center, University Hospital Zurich and University of Zurich, Zurich, Switzerland; Division of Pediatric Neurosurgery, University Children’s Hospital, Zurich, Switzerland

**Author notes:** **Corresponding author:** Kevin Akeret, MD, Department of Neurosurgery, Clinical Neuroscience Center, Universitätsspital and University of Zurich, Frauenklinikstrasse 10, CH-8091 Zurich, Switzerland.

**Keywords:** Glioma, Metastases, Phylo-ontogenetic radial units, Primary central nervous system lymphoma, Topography

## Abstract

In contrast to most other tumors, the anatomical extent of brain tumors is not objectified and quantified through staging. Staging systems are built on the understanding of the anatomical sequence of tumor progression and its relation to histopathological dedifferentiation and survival. While major advances in the understanding of primary brain tumors at a histological, cellular and molecular level have been achieved in recent decades, our understanding at a macroscopic anatomical level is limited. The aim of this study was to describe the anatomical phenotype of the most frequent brain tumor entities based on topographic probability and growth behavior analysis. The association of anatomical tumor features with survival probability was assessed and a prototypical staging system for WHO grade II-IV glioma was proposed based on the hypothesized anatomical sequence of tumor progression. The analysis is based on data from a consecutive cohort of 1000 patients with first diagnosis of a primary or secondary brain tumor. On preoperative MRI, the relative tumor density (RTD) of different topographic, phylogenetic and ontogenetic parcellation units was derived through normalization of the relative tumor prevalence to the relative volume of the respective structure. While primary central nervous system lymphoma (PCNSL) showed a high RTD along white matter tracts, the RTD in metastases was highest along terminal arterial flow areas. Neuroepithelial tumors (NT) demonstrated a high and homogeneous RTD along all sectors of the ventriculo-cortical axis, avoiding adjacent units, consistent with a transpallial behavior within phylo-ontogenetic radial units. Additionally, the topographic probability in NT correlated with morphogenetic processes of convergence and divergence of radial units during phylo- and ontogenesis. The anatomical tumor growth behavior was analyzed by comparing pre- and postoperative MRI, showing that a ventriculofugal growth dominates in NT. With progressive histopathological dedifferentiation of NT, a gradual deviation from this neuroepithelial anatomical behavior was found. By comparing survival probability, we identified prognostically critical steps in the anatomical behavior of NT. Based on a hypothesized sequence of anatomical tumor progression, we developed a three-level prototypical staging system for WHO grade II-IV glioma. This staging system proved to be accurate across histological, molecular, radiomorphological and clinical strata based on Kaplan Meier curves and multivariable survival analysis. Similar to staging systems for other tumors, a staging system such as this one may have the potential to inform stage-adapted treatment decisions.

## Introduction

Primary and secondary brain tumors cause high mortality and morbidity globally and require considerable diagnostic and therapeutic resources^1^. The basis for new and more tailored therapies is an enhanced understanding of the underlying pathophysiology of and interindividual differences between tumors^2^. On histological, cellular and molecular levels, fundamental progress has been achieved in recent decades. The current classification of brain tumors and therapeutic guidelines reflect the insights into these pathophysiological processes^3,4^. However, our understanding of the pathophysiological processes at the macroscopic anatomical level is substantially less developed. Also, this dimension is not represented in the classification of brain tumors. Although the spatial extent of a brain tumor often influences therapeutic decision making, this is based on qualitative experience-based assessment. In contrast, in most other tumors, the anatomical extent is objectified and quantified through staging^5^. The TNM system for solid tumors integrates local (T), regional (N) and systemic (M) spatial criteria based on a known sequence of anatomical tumor progression. This parallels histopathological dedifferentiation, is a fundamental determinant of prognosis and drives treatment decisions. The basis for the development of a staging system is thus an understanding of the anatomical sequence of tumor progression and its association with histopathological tumor characteristics and survival.

At least in part, the lack of a staging system for brain tumors could be attributed to the brain’s anatomical complexity and the associated obscuring of the anatomical sequence of tumor progression. The micro- and macroanatomy of the brain is determined by the histogenetic and morphogenetic processes during phylo- and ontogenesis^6–8^. On a macroscopic anatomical level, little is known about the specific interaction of tumors with the brain. It has remained uncertain where exactly brain tumors develop and how they evolve spatially. On the cellular and molecular levels, gliomagenesis displays retrograde parallels to ontogenesis^9,10^. Such similarities may also be present on the macroscopic anatomical level. Distinct differences in the anatomical patterns of neuroepithelial tumors (NT), primary central nervous system lymphoma (PCNSL) and metastases have been demonstrated^11^. A more profound understanding of brain tumor anatomy in the context of phylo- and ontogenesis and its influence on survival may provide the basis for an enhanced pathophysiological understanding and the development of a brain tumor staging system.

The aim of this study was to describe the anatomical phenotype of the most frequent brain tumor entities based on topographic probability and growth behavior analysis. The association of anatomical tumor features with survival probability was characterized and a prototypical staging system for brain tumors proposed based on the hypothesized anatomical sequence of tumor progression.

## Methods

This study was approved by the ethical review board of the Canton of Zurich, Switzerland (KEK ZH 01120). Written informed consent was obtained from all patients or their legal representatives. The results are reported in accordance with the STROBE statement^12^.

### Source population

The data were obtained from a consecutive cohort of patients with a newly diagnosed brain tumor admitted to the University Hospital Zurich during an eight-year period (January 2009 to December 2016). The eligibility criteria comprised: (i) first diagnosis with consecutive histopathologic confirmation of a primary or secondary brain tumor; (ii) no pretreatment or previous cranial surgery; (iii) intraparenchymal encephalic tumor location (supra-, infratentorial or both); (iv) availability of complete preoperative MRI data.

### Data acquisition

Supplemental Table 1 provides the demographic, clinical and histopathologic data obtained with the respective specifications. A 3-tesla Skyra VD13 (Siemens Healthcare, Erlangen, Germany) with a 32-channel receive coil was used for acquisition of T1-weighted Magnetization Prepared Rapid Acquisition Gradient Echo images with and without contrast agent, T2-weighted and fluid-attenuated inversion recovery (FLAIR) images. Histopathologic analysis was performed by the Institute of Neuropathology, University Hospital Zurich.

### Neuroanatomical parcellation

The topographic parcellation of the brain followed a recently published protocol^13^. The cerebral and cerebellar white matter was divided into sectors according to Yasargil^11,14^. The phylogenetic and ontogenetic parcellation protocols are described in Supplemental Table 2.

### Image analysis

Image analysis was performed by two blinded investigators (KA, CS), with a third serving to reach consensus in cases of disagreement (NK). On the preoperative MRI closest to surgery, each topographic, phylogenetic and ontogenetic parcellation unit was assessed for tumor involvement. Those units that were only rated displaced or edematous, but not invaded, were classified as non-affected. The consultation of the post-resection MRI study assisted in this decision. The tumor was delineated by combining the morphological sequences (T1, T1 with contrast agent, T2, FLAIR). For singular tumors, a manual segmentation and volumetric analysis was performed using 3DSlicer 4.10.2^15^. The contrast agent behavior was assessed for all tumors (contrast enhancement, central necrosis). The anatomical tumor behavior in the period before and after surgery was analyzed by comparing the MRI study undertaken immediately preoperatively with previous MRI studies of the patient and with postoperative follow-up MRI studies, respectively. A minimum interval of one month between MRI studies was required.

### Tumor probability

A parcellation unit’s relative tumor prevalence (RTP) was derived from the frequency of involvement of the respective structure in relation to the overall prevalence of the tumor entity. By normalizing the RTP to the relative volumes of the respective structures using a topographic volume-standardization atlas^13^, the relative tumor density (RTD) of a parcellation unit was calculated. The volumes of the phylogenetic and ontogenetic parcellation units (Supplemental Table 3) were generated using the reference atlas^13^ and the protocol described in Supplemental Table 2.

### Statistical analysis

All statistical analyses were performed using R 4.0.0 (R Core Team, 2020). The statistical code is provided in the Supplemental Materials, including the versions of the R-packages used. Descriptive statistics are presented as absolute numbers (n) and proportions (%) with a 95% Wilson confidence interval for categorical variables, whereas continuous variables are shown as mean and standard deviation (SD). Survival probabilities were plotted using the Kaplan Meier method and pairwise comparisons between subgroups were carried out using the log-rank test with Bonferroni-Holm correction for multiple testing^16^. For multivariable survival analysis, the Cox proportional hazards regression model was used^17,18^. The proportional hazards assumption was tested for each covariable using the scaled Schoenfeld residuals. In case the proportional hazards assumption was not met, the variable was included in the model as a stratum. The linearity assumption for continuous variables was checked using the Martingale residuals. Unadjusted models were compared to models with adjustment for tumor volume and treatment modality (type of surgery, chemotherapy, radiotherapy). Due to the exploratory character of this study, no level of statistical significance was defined: instead, the results were interpreted based on the level of evidence: p < 0.001: very strong evidence; p < 0.01: strong evidence; p < 0.05 evidence; p < 0.1 weak evidence; p > 0.1: no evidence^19^.

### Data availability

The authors confirm that the data supporting the findings of this study are available within the article and its Supplemental Materials. The statistical code and the anonymized datasets will be made available on GitHub at https://github.com/KevinAkeret/Anatomical-phenotyping-and-staging-of-brain-tumors. The use of dynamic reporting guarantees full reproducibility of the results.

## Results

Of 1051 screened patients, the study included 1000. The exclusions were due to unclear histological results (n = 25, 49%), incomplete or qualitatively insufficient MRI data (n = 23, 45.1%) or previous cranial surgery (n = 3, 5.9%). The mean age of the included patients was 53.8 years (SD 19.6) and 583 were men (58.3%). There were 657 (65.7%) patients with NT, 299 (29.9%) with metastases and 44 (4.4%) with PCNSL. The most frequent histological subtype was glioblastoma (n = 385, 60.6%), followed by lung cancer metastases (n = 143, 47.8%), WHO grade III glioma (n = 110, 17.3%; comprising WHO grade II astrocytoma, oligoastrocytoma and oligodendroglioma), WHO grade II glioma (n = 50, 7.9%;; comprising WHO grade III astrocytoma, oligoastrocytoma and oligodendroglioma) and melanoma metastases (n = 49, 16.4%). Supplemental Table 4 provides the histopathologic details of the study population. The Karnofsky Performance Status (KPS) at time of surgery is given in Supplemental Table 5. The tumor was resected in 806 (80.6%) patients and a biopsy was performed in 194 (19.4%). Radiotherapy was given to 759 patients (78.2%) and chemotherapy to 657 (68.2%).

### Topographic tumor probability identifies specific anatomical phenotypes

First, we characterized the RTP and RTD in relation to general topographic, phylogenetic and ontogenetic units (Supplemental Tables 6-8, Fig. 1A-I). The highest NT RTD was observed in the insular lobe, the archi-/paleopallium and the medial pallium. Developmental tumors, WHO grade II-III glioma and glioblastoma resembled each other in their anatomical phenotype – topographically, phylogenetically and ontogenetically. There was, however, a gradual loss of anatomical specificity along this sequence. A different anatomical phenotype was found in ependymoma, pilocytic astrocytoma and medulloblastoma. Pilocytic astrocytoma showed a high RTD in the hypothalamus and medulloblastoma in the archi-/paleocerebellum. The highest RTD for PCNSL was found in the mesencephalon. Metastases had the highest RTD in the neocerebellum, followed by the occipital and the central lobes. Comparable topographic, phylogenetic and ontogenetic phenotypes were found with respect to the different origins of metastases. The exception was for melanoma metastases, which showed a poorer topographic specification and a proportionally high RTD in the archi-/paleopallium and medial pallium.

**Fig 1.**
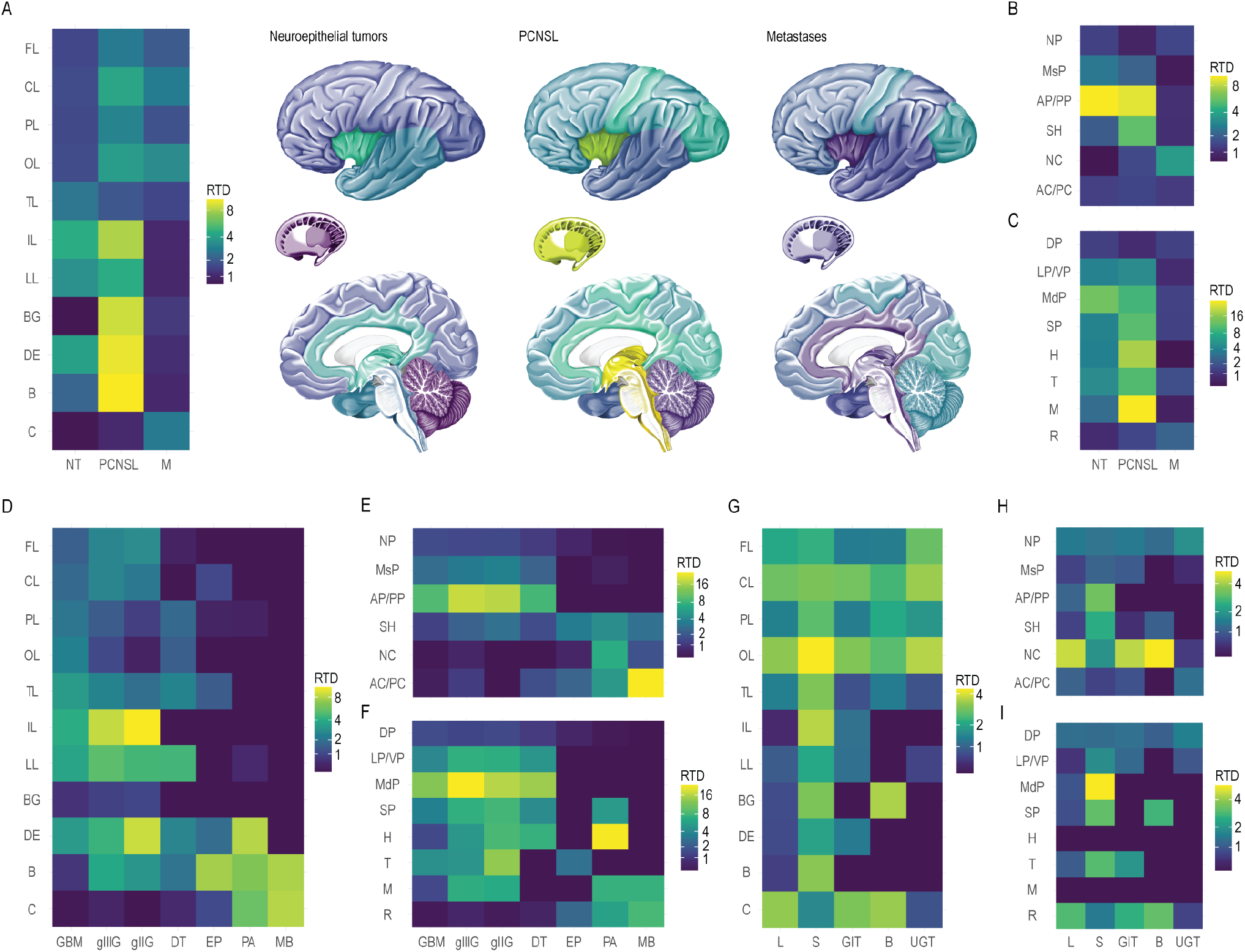
Relative tumor density within general topographic, phylogenetic and ontogenetic parcellation units: **A-C**. Relative tumor density (RTD) of neuroepithelial tumors (NT), primary CNS lymphoma (PCNSL) and metastases (M) stratified to their general (**A**.) topographic anatomy (cerebral lobes, basal ganglia, diencephalon, brainstem and cerebellum), (**B**.) phylogenetic anatomy or (**C**.) ontogenetic anatomy. *AC/PC = archicerebellum/paleocerebellum; AP/PP = archipallium/paleopallium; BG = basal ganglia; C = cerebellum; CL = central lobe; DE = diencephalon; DP = dorsal pallium; FL = frontal lobe; H = hypothalamus; IL = insular lobe; LL = limbic lobe; LP/VP = lateral pallium / ventral pallium; M = mesencephalon; MdP = medial pallium; MsP = mesopallium; NC = neocerebellum; NP = neopallium; OL = occipital lobe; PL = parietal lobe; R = rhombencephalon; SH = stammhirn; SP = subpallium; T = thalamus; TL = temporal lobe*. **D-F**. RTD of histopathologic neuroepithelial subtypes stratified to their (**D**.) topographic anatomy, (**E**.) phylogenetic anatomy or (**F**.) ontogenetic anatomy. *DT = developmental tumors; EP = ependymoma; gIIG = WHO grade II glioma (astrocytoma, oligoastrocytoma, oligodendroglioma); gIIIG = WHO grade III glioma (astrocytoma, oligoastrocytoma, oligodendroglioma); GBM = glioblastoma; MB = medulloblastoma; PA = pilocytic astrocytoma*. **G-I**. RTD of metastases with respect to the organ of origin stratified to their (**G**.) topographic anatomy, (**H**.) phylogenetic anatomy or (**I**.) ontogenetic anatomy. *B = breast; GIT = gastrointestinal tract; L = lung; S = skin; UGT = urogenital tract*.

Second, we characterized the RTP and RTD of brain tumors on a gyral level. Gyral RTP and RTD were compared between NT and metastases (Supplemental Table 9, Fig. 2A), between different NT entities (Supplemental Table 10, Fig. 2B) and between metastases of different origin (Supplemental Table 11, Fig. 2C). Due to extensive (>10) gyral involvement, 11 NT (1.7%) and 9 metastases (3%) were excluded from this analysis. PCNSL were not included due to the limited number of cases. The highest NT RTD was found in the insular, opercular, mediofrontal and temporomesial gyri. This was true for developmental tumors, WHO grade II-III glioma and glioblastoma, with a loss of anatomical specificity along this sequence. In metastases, the highest RTD was found in the inferior temporal gyrus, the middle occipital gyrus and the cuneus. In contrast to metastases of other origins, melanoma metastases were characterized by high RTD in the amygdala and hippocampus.

**Fig 2.**
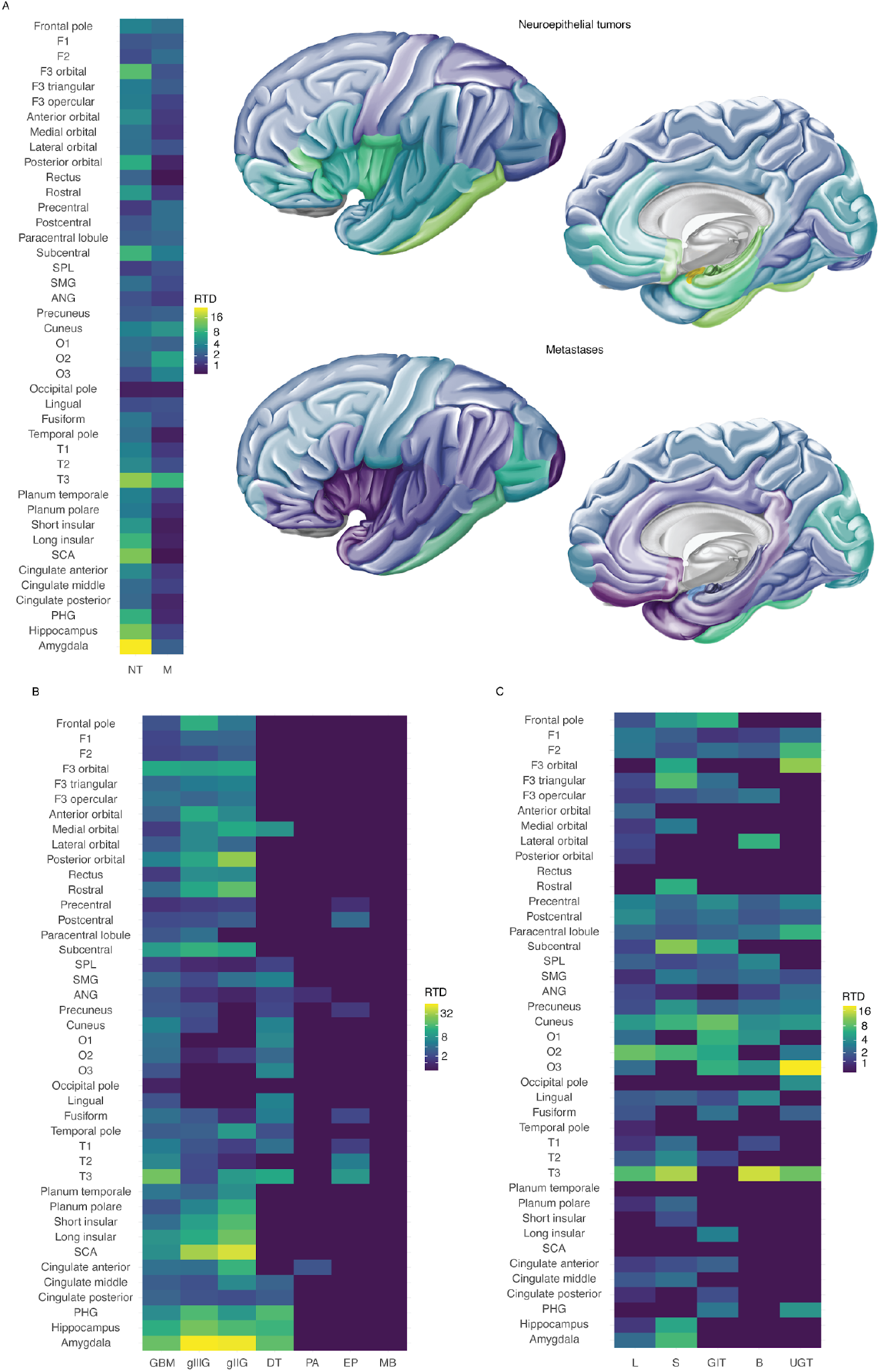
Relative tumor density within gyral parcellation units: **A**. Relative tumor density (RTD) of neuroepithelial tumors (NT) and metastases (M) stratified to their gyral anatomy. *ANG = angular gyrus; F1 = superior frontal gyrus; F2 = middle frontal gyrus; F3 = inferior frontal gyrus; O1 = superior occipital gyrus; O2 = middle occipital gyrus; O3 = inferior occipital gyrus; PHG = parahippocampal gyrus; SCA = subcallosal area; SMG = supramarginal gyrus; SPL = superior parietal lobule; T1 = superior temporal gyrus; T2 = middle temporal gyrus; T3 = inferior temporal gyrus*. **B**. RTD of histopathologic neuroepithelial subtypes stratified to their gyral anatomy. *DT = developmental tumors; EP = ependymoma; gIIG = WHO grade II glioma (astrocytoma, oligoastrocytoma, oligodendroglioma); gIIIG = WHO grade III glioma (astrocytoma, oligoastrocytoma, oligodendroglioma); GBM = glioblastoma; MB = medulloblastoma; PA = pilocytic astrocytoma*. **C**. RTD of metastases with respect to the organ of origin stratified to their gyral anatomy. *B = breast; GIT = gastrointestinal tract; L = lung; S = skin; UGT = urogenital tract*.

Third, the central supratentorial and infratentorial RTP and RTD were studied (Supplemental Tables 12-17, Supplemental Fig. 1). NT RTD was generally low in the central supratentorium and brainstem, with the internal capsule showing the lowest RTD. Pilocytic astrocytoma, however, demonstrated a high RTD in the hypothalamus. There was a loss of anatomical specificity along the sequence from developmental tumors, WHO grade II-III glioma to glioblastoma. In cerebellar NT, a higher RTD was observed in the vermis than in the hemisphere. Comparing cerebellar lobes, the flocculonodular lobe demonstrated the highest NT RTD. Among NT entities, medulloblastoma and pilocytic astrocytoma dominated in the cerebellum. PCNSL showed a high RTD in the central supratentorium and the brainstem, especially the globus pallidum, internal capsule and mesencephalon. PCNSL were not analyzed in relation to detailed cerebellar anatomy due to the limited number of cases. Metastases were almost non-existent in the central supratentorium and the brainstem. Cerebellar metastases presented with comparable RTD in the vermis and hemispheres. The highest RTD for metastases was found in the posterior cerebellar lobe. The same patterns were observed after stratification according to the origin of the metastases. The flocculonodular lobe remained spared across all strata.

Next, we characterized brain tumor anatomy in relation to the ventricular system (Fig. 3). First, the RTP and RTD along the different ventricular segments were investigated (Supplemental Tables 18-19, Fig. 3A-B). NT demonstrated an anatomical specificity for the temporal horn of the lateral ventricle and the lateral recess of the fourth ventricle. Developmental tumors, WHO grade II-III glioma and glioblastoma showed association with the temporal horn. A decrease in anatomical specificity along this sequence was observed. Pilocytic astrocytoma, ependymoma and medulloblastoma were associated with the lateral recess. The third ventricle showed the strongest association with pilocytic astrocytoma. PCNSL demonstrated a high RTD along the wall of all ventricular segments, highest in the fastigium of the fourth ventricle. The RTD of metastases was very low along all segments. Second, we compared the RTP and RTD between univentriculosegmental and multiventriculosegmental NT, highlighting the anatomical specificity of univentriculosegmental NT for the temporal horn and lateral recess (Supplemental Table 20, Fig. 3C). Third, we characterized the topological ventriculo-parenchymal phenotype, i.e. the relationship between ventricular and parenchymal tumor anatomy (Supplemental Table 21, Fig. 3D, Supplemental Fig. 2). Insular tumors were found to interrelate topologically with all segments of the lateral ventricle.

**Fig 3.**
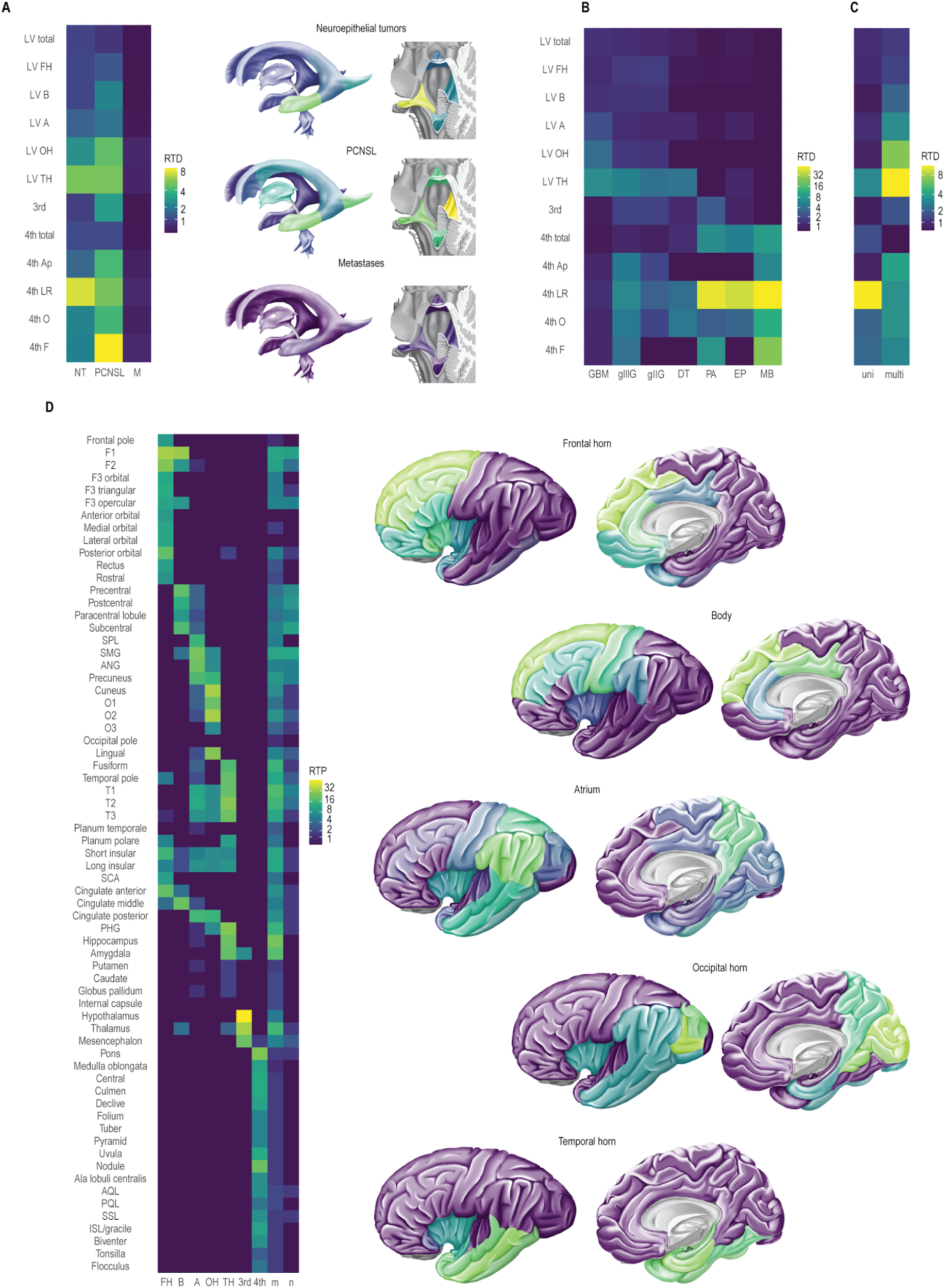
Relative tumor density along the ventricles and ventriculo-parenchymal topological relationship in neuroepithelial tumors: **A**. Relative tumor density (RTD) of neuroepithelial tumors (NT), primary CNS lymphoma (PCNSL) and metastases (M) stratified to their ventricular anatomy. *3rd = third ventricle; 4th = fourth ventricle; A = atrium; Ap = apex; B = body; F = fastigium; FH = frontal horn; LR = lateral recess; LV = lateral ventricle; O = obex; OH = occipital horn; TH = temporal horn*. **B**. RTD of histopathologic neuroepithelial subtypes stratified to their ventricular anatomy. *DT = developmental tumors; EP = ependymoma; gIIG = WHO grade II glioma (astrocytoma, oligoastrocytoma, oligodendroglioma); gIIIG = WHO grade III glioma (astrocytoma, oligoastrocytoma, oligodendroglioma); GBM = glioblastoma; MB = medulloblastoma; PA = pilocytic astrocytoma*. **C**. RTD of NT with contact with one (univentriculosegmental = uni) or multiple (multiventriculosegmental = multi) anatomical segments of the ventricular system stratified to their ventricular anatomy. **D**. Ventriculo-parenchymal topological relationship of NT, providing the tumor prevalence in each anatomical parcellation unit relative to the ventricular contact. *ANG = angular gyrus; AQL = anterior quadrangular lobule; F1 = superior frontal gyrus; F2 = middle frontal gyrus; F3 = inferior frontal gyrus; ISL = inferior semilunar lobule; m = multi; n = none; O1 = superior occipital gyrus; O2 = middle occipital gyrus; O3 = inferior occipital gyrus; PHG = parahippocampal gyrus; PQL = posterior quadrangular lobule; RTP = relative tumor prevalence; SCA = subcallosal area; SMG = supramarginal gyrus; SPL = superior parietal lobule; SSL = superior semilunar lobule; T1 = superior temporal gyrus; T2 = middle temporal gyrus; T3 = inferior temporal gyrus*.

To complement the analyses with respect to a radial axis, we examined the RTP within ventriculocortical parcellation units (Supplemental Tables 22-24, Fig. 4). In NT, the anatomical sectors between cortex and ventricle (cortex, subcortical, subgyral, gyral and lobar white matter sector, lateral ventricle ependyma) showed a high and homogeneous RTP. Diffuse ependymal involvement or a subependymal spread along the corpus callosum was associated with an intermediate RTP. The central corpus callosum, and especially the internal capsule, were found to have a very low NT RTP. The comparison of WHO grade II-III glioma and glioblastoma revealed similar phenotypes; however, diffuse ependymal as well as subependymal spreading along the corpus callosum increased along this sequence. An involvement of the internal capsule was exclusively found in glioblastoma (1.1% [0.4-2.8%]). The same applied to the central corpus callosum (6.2% [4.2-9.2%]), with the exception of a single case of WHO grade III glioma (1% [0.2-5.5%]). PCNSL spared the cortex, showed a high RTP across all white matter sectors, the internal capsule and the central corpus callosum, as well as exclusively diffuse ependymal involvement. Metastases remained limited to the cortex, subcortical and subgyral white matter. We also determined the RTP along the radial units in the cerebellum. Again, NT demonstrated a high and homogeneous RTP along all ventriculocortical radial parcellation units, while cerebellar nuclei and peduncles remained spared. PCNSL had a low RTP in the cortex, while it was high in the cerebellar nuclei and peduncles. Like cerebral metastases, cerebellar metastases demonstrated a high RTP in the cortex and the superficial white matter sectors.

**Fig 4.**
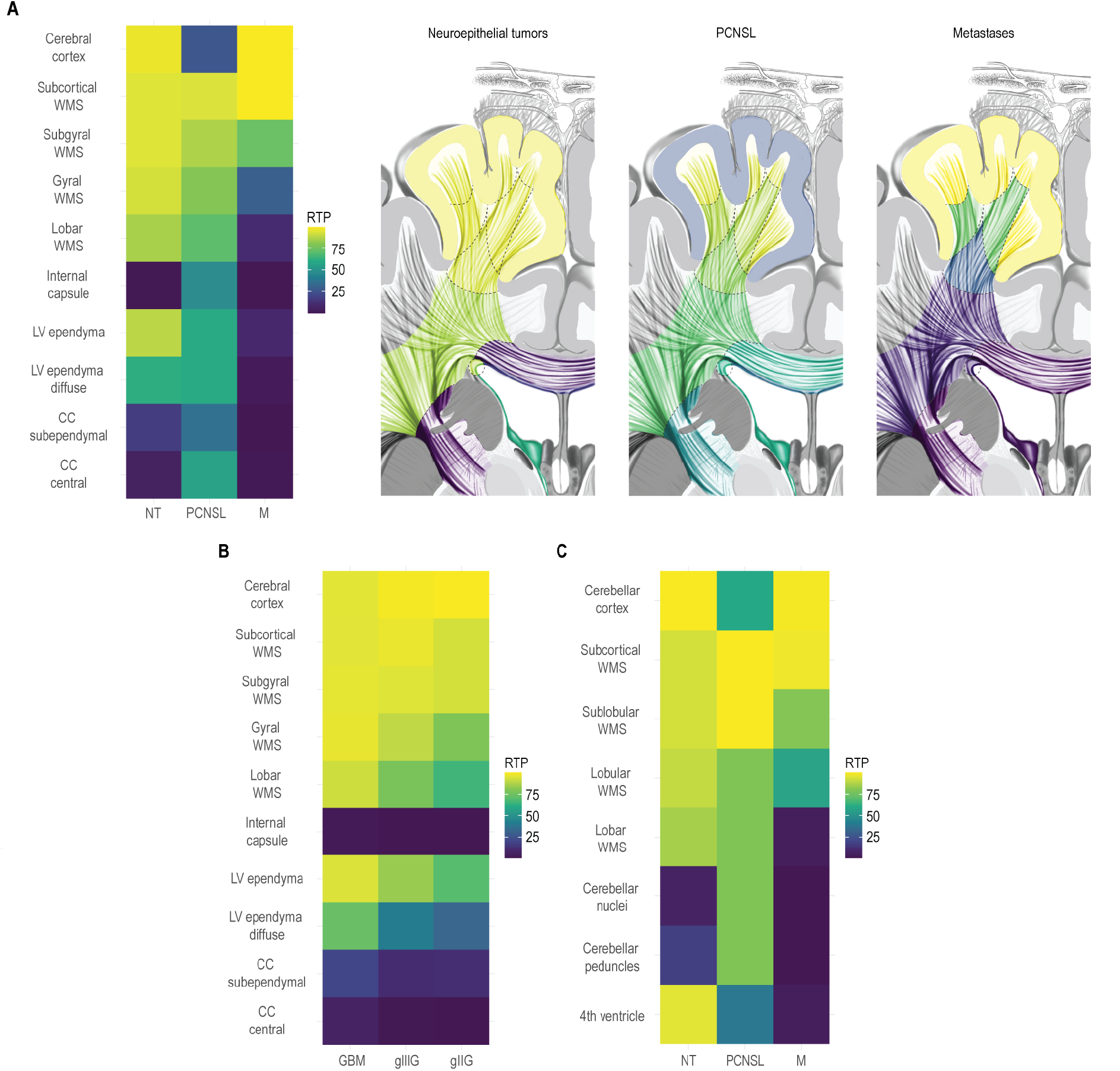
Relative tumor prevalence within cerebral and cerebellar radial sectors: **A**. Relative tumor prevalence (RTP) of neuroepithelial tumors (NT), primary CNS lymphoma (PCNSL) and metastases (M) stratified to their radial cerebral anatomy. *CC = corpus callosum; LV = lateral ventricle; WMS = white matter sector*. **B**. RTP of glioblastoma (GBM), WHO III (gIIIG) and WHO grade II (gIIG) glioma stratified to their radial cerebral anatomy. **C**. RTP of NT, PCNSL and M stratified to their radial cerebellar anatomy.

In summary, NT, PCNSL and metastases demonstrated specific anatomical phenotypes. In addition, differences in the anatomical phenotypes were found depending on the NT entity. With progressive histopathological dedifferentiation, a decrease in anatomical specificity was observed.

### Tumor growth follows distinct anatomical patterns

We added a temporal dimension to the analysis of the anatomical behavior of brain tumors by examining both preoperative and postoperative tumor growth (Supplemental Tables 25-27, Fig. 5). In NT, the dominant growth was ventriculofugal. Spherical growth was observed in developmental tumors, ependymoma and pilocytic astrocytoma. Subependymal growth was found in particular in WHO grade III glioma and glioblastoma. PCNSL generally appeared to grow along white matter tracts. In metastases, the growth was predominantly spherical or new independent lesions were found. Thus, NT, PCNSL and metastases differed in their anatomical growth patterns.

**Fig 5.**
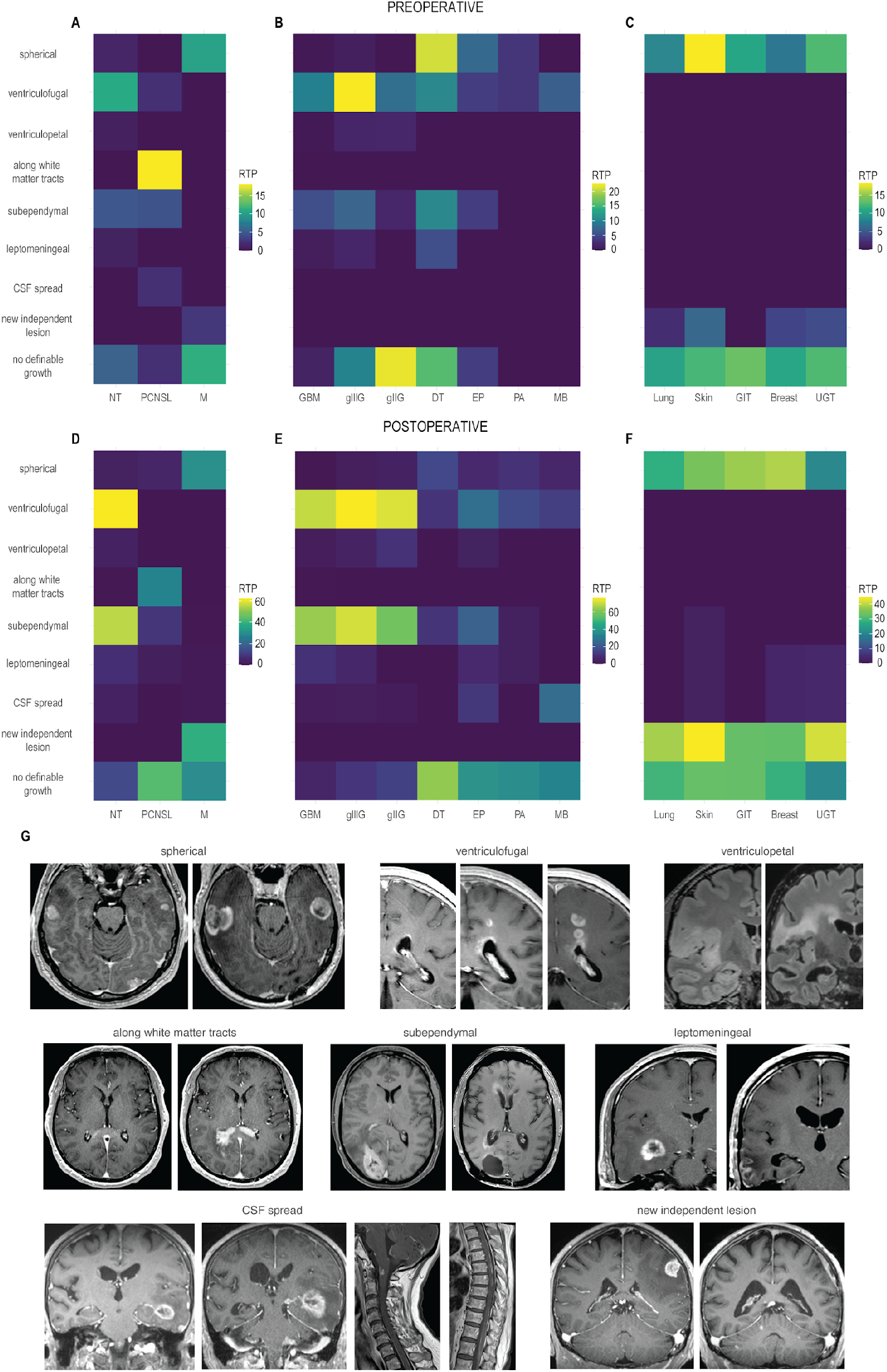
Brain tumor growth patterns: Relative tumor prevalence (RTP) of different tumor growth patterns on pre- (**A**., **B**., **C**.) and postoperative (**D**., **E**., **F**.) follow-up in (**A**., **D**.) neuroepithelial tumors (NT), primary CNS lymphoma (PCNSL) and metastases (M). *CSF = cerebrospinal fluid*; in (**B**., **E**.) different histopathologic neuroepithelial subtypes. *DT = developmental tumors; EP = ependymoma; gIIG = WHO grade II glioma (astrocytoma, oligoastrocytoma, oligodendroglioma); gIIIG = WHO grade III glioma (astrocytoma, oligoastrocytoma, oligodendroglioma); GBM = glioblastoma; MB = medulloblastoma; PA = pilocytic astrocytoma*; in (**C**., **F**.) metastases with respect to the organ of origin. *B = breast; GIT = gastrointestinal tract; L = lung; S = skin; UGT = urogenital tract*. **G**. Exemplified MR images of tumor growth patterns.

### Survival analysis determines prognostically critical steps in the anatomical behavior of neuroepithelial tumors

Considering statistical validity, the analyses regarding the association between anatomical tumor features and survival probability were limited to general topographic, ventricular and radial anatomical tumor features (Supplemental Tables 28-32, Fig. 6). NT, in their entirety, were compared to glioblastoma, the most common NT entity. There was evidence for survival differences in NT associated with general topographic anatomical tumor features (Supplemental Table 28, Fig. 6A). Within glioblastoma, however, no evidence for such an association remained. Additionally, associations in univentriculosegmental tumors between survival and ventricular topography, which were identified in NT, were not confirmed in the analysis of glioblastoma (Supplemental Table 29, Fig. 6B). However, the analysis provided very strong to strong evidence for a difference in survival between NT without ventricular contact, NT with unisegmental ventricular contact and NT with multisegmental ventricular contact (Supplemental Table 30, Fig. 6C). Within glioblastoma, there was no evidence for a difference between tumors without ventricular contact and those with unisegmental ventricular contact (p = 0.19). However, there was strong to very strong evidence of a survival difference compared to multisegmental glioblastoma. Both, NT and glioblastoma, showed no association between survival probability and the ventriculocortical sectors involved (Supplemental Table 31, Fig. 6D). In NT and in glioblastoma, there was strong evidence for a difference in survival between tumors with focal contact with the ependyma and those with diffuse ependymal involvement, subependymal spread along the corpus callosum, central involvement of the corpus callosum or involvement of the internal capsule (Supplemental Table 32, Fig. 6E). In addition, there was evidence for a difference in survival between NT with diffuse ependymal involvement and those with central corpus callosum involvement. This is consistent with the grouping for NT in the Kaplan Meier curves: best survival with focal ependymal involvement; intermediate survival with diffuse ependymal or subependymal corpus callosum pattern; and poorest survival with internal capsule or central corpus callosum involvement. The same pattern, although less pronounced, was observed in glioblastoma. In summary, certain specific anatomical tumor features, e.g. the nature of ventricular or white matter tract involvement, appear to be associated with survival probability in neuroepithelial tumors.

**Fig 6.**
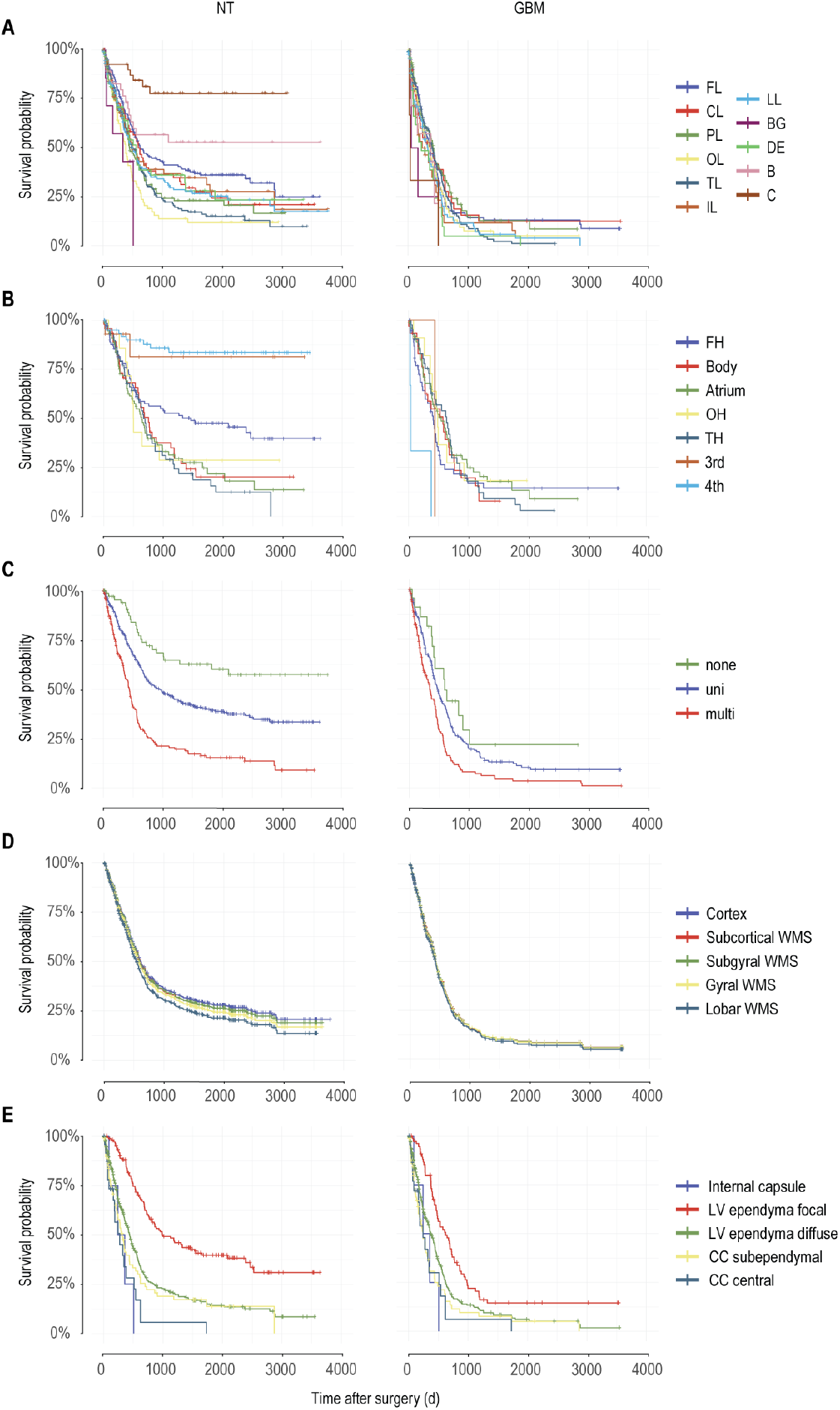
Association between anatomical features of neuroepithelial tumors and survival: **A**. Kaplan-Meier (KM) curves showing the survival probability for neuroepithelial tumors (NT, left) and glioblastoma (GBM, right) stratified to their topographic anatomy. *B = basal ganglia; C = cerebellum; CL = central lobe; d = days; DE = diencephalon; FL = frontal lobe; H = hypothalamus; IL = insular lobe; LL = limbic lobe; M = mesencephalon; OL = occipital lobe; PL = parietal lobe; T = thalamus; TL = temporal lobe*. **B**. KM curves showing the survival probability for univentriculosegmental NT (left) and GBM (right) stratified to their ventricular contact. *3rd = third ventricle; 4th = fourth ventricle; A = atrium; B = body; FH = frontal horn; LV = lateral ventricle; OH = occipital horn; TH = temporal horn*. **C**. KM curves showing the survival probability for NT (left) and GBM (right) stratified to their ventriculosegmentality. *multi = multiventriculosegmental, none = no ventricular contact*; *uni = univentriculosegmental*. **D**. KM curves showing the survival probability for NT (left) and GBM (right) stratified to the involvement of the radial lobar sectors. *WMS = white matter sector*. **D**. KM curves showing the survival probability for NT (left) and GBM (right) stratified to the invasion of the internal capsule, focal or diffuse involvement of the lateral ventricular (LV) ependyma and subependymal or central involvement of the corpus callosum (CC).

### Prototypical staging of neuroepithelial tumors by anatomical differentiation

Lobar WHO grade II-III glioma and glioblastoma were categorized into three different anatomical stages of neuroepithelial differentiation, based on anatomical criteria determined by the pathophysiological rationale derived from the findings for tumor probability and survival analyses (Fig. 7A): an invasion of the central corpus callosum or internal capsule was considered the criterion for *Stage III*. If this was not met, but multiple ventricular segments were involved or a diffuse ventricular or subependymal corpus callosum spread was present, then it was considered a *Stage II*. If neither of the above anatomical dedifferentiation criteria was met, the tumor was defined as *Stage I*. Along the sequence of WHO grade II-III glioma and glioblastoma, a progression towards dedifferentiated anatomical behavior was found (Supplemental Table 33). The staging system was internally evaluated for accuracy within different histological, molecular, radiomorphological and clinical strata using Kaplan Meier curves and multivariable survival analysis (Fig. 7B-G). The choice between including the variables as covariables or stratifying variables in the Cox proportional hazards model was made based on scaled Schoenfeld residuals testing (Supplemental Fig. 3). In each case, an unadjusted model was compared with an adjusted model. The unadjusted model included only the proposed tumor stages as covariables and the respective histological, molecular, radiomorphological or clinical categories as stratifying variables. The adjusted model additionally included tumor volume as a continuous covariable and the type of surgical intervention (biopsy or resection), chemotherapy or radiotherapy as stratifying binary variables. In both the Kaplan Meier curves and the multivariable survival analyses, tumor staging proved plausible and consistent across the different strata. Within the multivariable survival analyses, the adjustment for tumor volume and treatment modality did not lead to a relevant change in the estimated hazard ratio for the different stages, although the confidence intervals partially widened. With Stage II as reference, the hazard ratio of Stage I tumors ranged from 0.39 to 0.73, that of Stage III tumors from 1.63 to 2.58. Collectively, this staging based on specific anatomical criteria was found to be consistent with the degree of histological dedifferentiation and accurate in terms of survival probability within different histological, molecular, radiomorphological and clinical strata.

**Fig 7.**
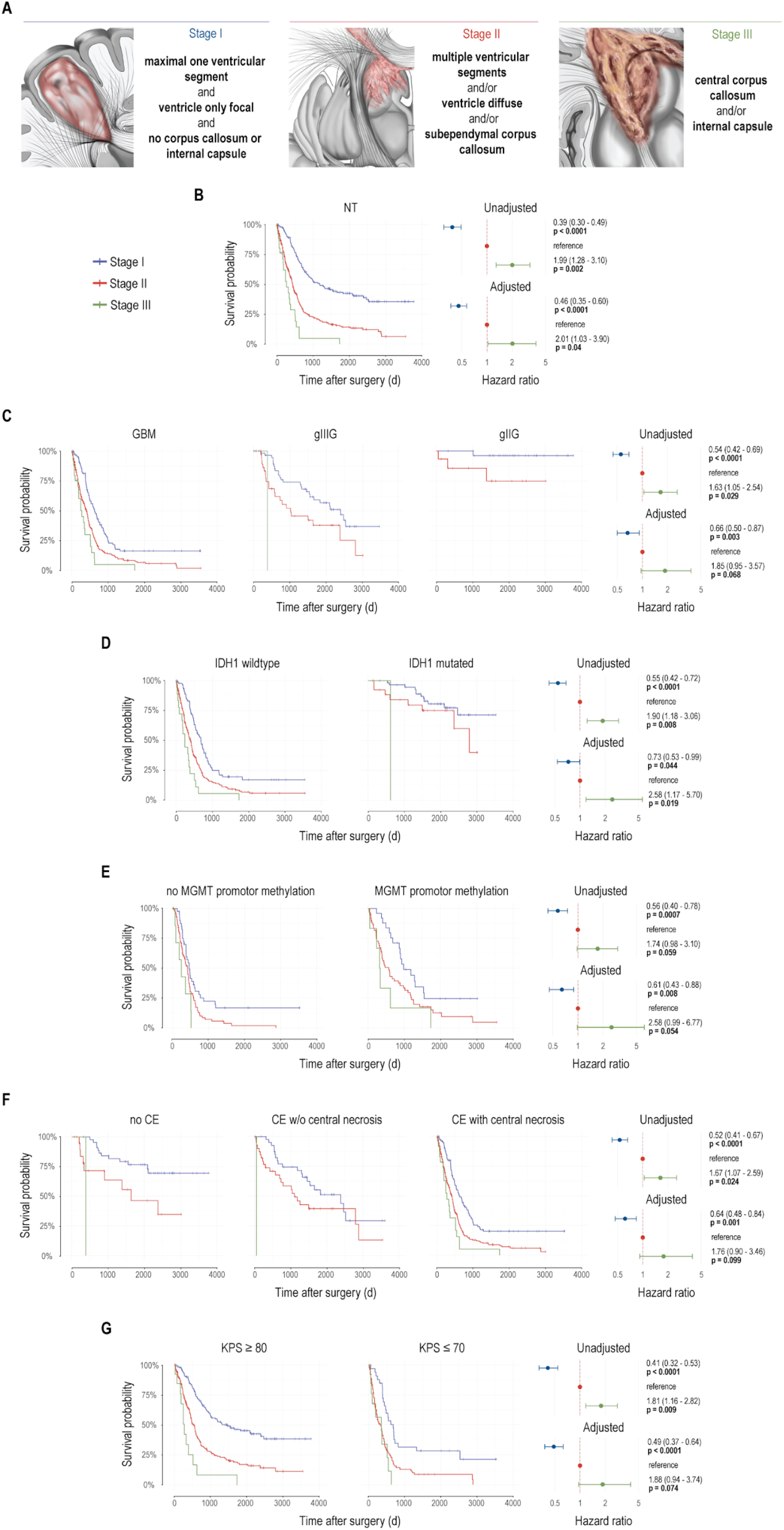
Stages of neuroepithelial dedifferentiation and association with survival: **A**. Schematic illustration of the proposed prototypical tumor stages based on topographic anatomical criteria. **B**. Left: Kaplan-Meier (KM) curves showing the survival probability for neuroepithelial tumors (NT) stratified to the anatomical tumor stages. *d = days*. Right: Forest plots providing the hazard ratios for Stage I and III tumors in relation to Stage II tumors with the respective 95% confidence intervals. In the adjusted model, tumor volume was used as a continuous covariable and type of surgery (biopsy or resection), chemotherapy and radiotherapy as binary stratifying variables. **B**. KM curves showing the survival probability for glioblastoma (GBM), WHO grade III (gIIIG) and WHO grade II (gIIIG) glioma (astrocytoma, oligoastrocytoma and oligodendroglioma) stratified to the anatomical tumor stages. Right: Forest plots providing the hazard ratios for Stage I and III tumors in relation to Stage II tumors with the respective 95% confidence intervals. In the unadjusted model, the histological neuroepithelial entity was used as a stratifying variable. In the adjusted model, tumor volume was used as a continuous covariable and type of surgery (biopsy or resection), chemotherapy and radiotherapy as binary stratifying variables. **C**. Left: KM curves showing the survival probability for IDH1 wildtype and IDH1 mutated NT stratified to the anatomical tumor stages. Right: Forest plots providing the hazard ratios for Stage I and III tumors in relation to Stage II tumors with the respective 95% confidence intervals. In the unadjusted model, the IDH1 status was used as a stratifying variable. In the adjusted models, tumor volume was used as a continuous covariable and type of surgery (biopsy or resection), chemotherapy and radiotherapy as binary stratifying variables. **D**. Left: KM curves showing the survival probability for NT with and without MGMT promoter methylation stratified to the anatomical tumor stages. Right: Forest plots providing the hazard ratios for Stage I and III tumors in relation to Stage II tumors with the respective 95% confidence intervals. In the unadjusted model, the MGMT promoter methylation status was used as a stratifying variable. In the adjusted models, tumor volume was used as a continuous covariable and type of surgery (biopsy or resection), chemotherapy and radiotherapy as binary stratifying variables. **E**. Left: KM curves showing the survival probability for neuroepithelial tumors without contrast enhancement, contrast enhancement without central necrosis and contrast enhancement with central necrosis, stratified to the anatomical tumor stages. Right: Forest plots providing the hazard ratios for Stage I and III tumors in relation to Stage II tumors with the respective 95% confidence intervals. In the unadjusted model, tumor contrast behavior was used as a stratifying variable. In the adjusted models, tumor volume was used as a continuous covariable and type of surgery (biopsy or resection), chemotherapy and radiotherapy as binary stratifying variables. **F**. Left: KM curves showing the survival probability for KPS ≥80 and ≤70 stratified to the anatomical tumor stages. Right: Forest plots providing the hazard ratios for Stage I and III tumors in relation to Stage II tumors with the respective 95% confidence intervals. In the unadjusted model, KPS was used as a stratifying variable. In the adjusted models, tumor volume was used as a continuous covariable and type of surgery (biopsy or resection), chemotherapy and radiotherapy as binary stratifying variables.

## Discussion

In this study, we identified specific anatomical phenotypes in different brain tumor entities through topographic probability and growth pattern assessment. Survival analysis determined prognostically critical steps in the anatomical behavior of neuroepithelial tumors. We developed a prototypical staging system for lobar WHO grade II-III glioma and glioblastoma based on the hypothesized anatomical sequence of tumor progression, which was found to be accurate across histological, molecular, radiomorphological and clinical strata.

The brain features a phylo- and ontogenetically determined natural coordinate system^20,21^. Perpendicular to the longitudinal neuraxis there are radially oriented units that relate the ventricular to the pial surface of the neural tube (Fig. 8A). A defining cell type within these units is radial glia cells (RGC); these have a periventricular cell body, a short projection to the ventricular surface and a long projection to the pial surface^22,23^. RGC are believed to serve as radially oriented migration scaffolds between the subventricular germinal zone and superficial brain regions during ontogenesis. RGC are also drivers of phylogenesis^24,25^: the evolutionary enlargement of brain regions, especially the surface of the neopallium and the neocerebellum, are attributable to a modification of the periventricular proliferation of RGC and an increase in the number of radial units^21,26–28^. Due to morphogenetic processes during phylo- and ontogenesis, the radial units become distorted^6–8^, their character as phylo-ontogenetic entities, however, likely persists (Fig. 8B)^29–31^.

**Fig 8.**
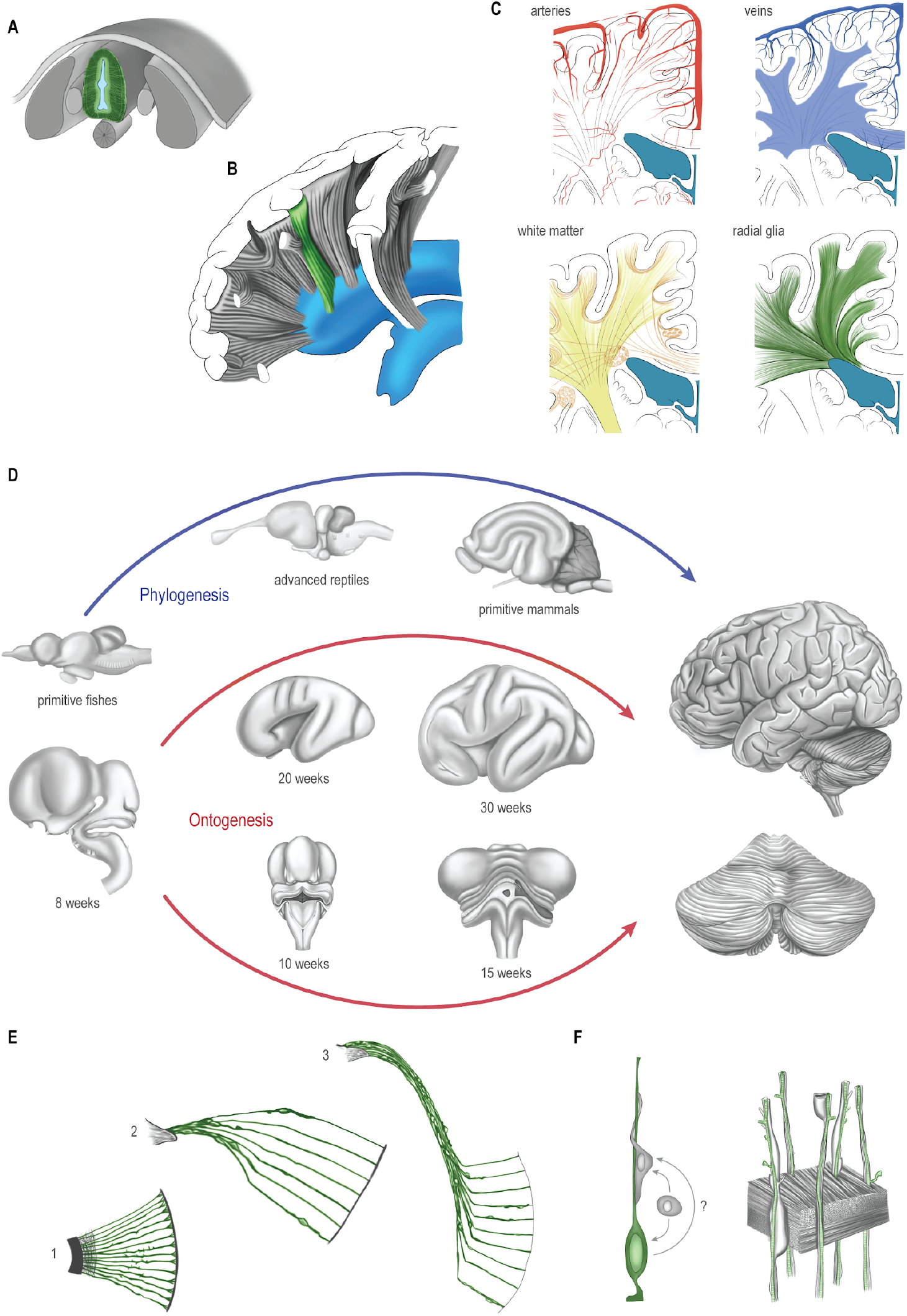
Architecture of the human brain: **A**. The natural coordinate system of the brain: neural tube with radially oriented units perpendicular to the neuraxis, relating the ventricular to the pial surface. Radial glia cells (RGCs) are shown in green. **B**. The concept of phylo-ontogenetic radial units persisting into adulthood. **C**. The radial cerebral anatomy of different potential tumor scaffolds: cerebral arteries, cerebral veins, white matter fiber systems, radial glia cells. **D**. Morphogenetic changes during phylo- and ontogenesis. A pronounced convergence occurs in the insular and opercular as well as in the vermal and paravermal areas. **E**. Schematic representation of the distortion of radial glial fibers during morphogenetic alterations (1-3), resulting in a varying density of radial glial fibers and associated stem cells. **F**. RGC-like cells may persist into adulthood and serve as tumor-stem cells and migration scaffold, or cells sharing characteristics of RGCs may emerge during gliomagenesis.

The radial anatomical tumor phenotypes described in this study can be compared to the orientation of possible scaffolds (Fig. 8C). NT showed a transpallial pattern characterized by a high RTP in all ventriculo-cortical sectors. Adjacent structures, however, such as the corpus callosum, basal ganglia or internal capsule, had a very low RTD. Such a transpallial pattern was a specific and consistent feature of NT and corresponds to an orientation along the phylo-ontogenetic radial unit. In contrast, PCNSL usually spared the cortex and often involved the corpus callosum, basal ganglia or internal capsule. This pattern can best be explained by an orientation along the white matter tracts and the ependyma. Metastases had a strong tendency to cortex, subcortical and subgyral sectors, while deep sectors were spared. This is compatible with the arterial angio-anatomy.

We also characterized the ventriculo-parenchymal topological relationship in NT. A comprehensive comparison of these with the orientation of the radial phylo-ontogenetic units is not possible, since a sufficient description of the latter for the adult human brain is not given^32^. Insular NT were associated with all segments of the lateral ventricle. This finding can be explained by the early phylo- and ontogenetic position of the insula in the lateral aspect of the pallium and hence its contact with all future segments of the lateral ventricle (Fig. 8D). The expansion of the neocortex and the involution of the insula likely result in a distortion of the radial units (Fig. 8E). This might explain the parabolic course of the ventricular contacts of insular tumors (Fig. 5G). Additionally, the trajectory of the ventricular contact in NT resembles the transmantle sign in focal cortical dysplasia type II^33^. Both patterns may be determined by the orientation of the radial phylo-ontogenetic units. The focal cortical dysplasia with a transmantle sign is likely the result of disturbed migration along the radial unit during ontogenesis. In NT, this might reflect neuroepitheloid behavior with neoplastic reactivation of a migration pathway of the same orientation, which simulates ventriculo-cortical migration during ontogenesis.

This study identified differences in parenchymal NT RTD along the longitudinal neuraxis. The highest NT RTD was found in the insular and opercular, mediofrontal and temporomesial structures. Phylogenetically, archi- and palleopallium demonstrated the highest NT RTD, followed by the mesopallium. Ontogenetically, the medial pallium showed the highest RTD. Infratentorially, the vermis and flocculonodular lobe had the highest NT RTD. The differences in parenchymal RTD could be attributable to morphogenetic processes (Fig. 8D). During phylo- and ontogenesis, there may be convergence of radial units in some regions of the brain and a divergence in others. A convergence of radial units potentially results in a local increase in stem cell density. A divergence may result in the opposite. Such a difference in stem cell density may cause a different stochastic risk for neoplasia. The parenchymal regions with high NT RTD (insula, operculum, mediofrontal and temporomesial structures; vermis and flocculonodular lobe) correspond to a converging morphogenesis (Fig. 8D)^32^. Brain areas with diverging character during phylo- or ontogenesis – for instance the neopallium, neocerebellum or dorsal pallium (Fig. 8D) – showed a low RTD.

In addition, along the ventricular wall, we identified differences in NT RTD. Supratentorially, the temporal horn showed the highest RTD, and infratentorially, the lateral recess had the highest RTD. NT entities differed in their RTD: developmental tumors, WHO II-III glioma and glioblastoma were associated with the temporal horn; pilocytic astrocytoma, ependymoma and medulloblastoma showed the highest RTD in the wall of the lateral recess of the fourth ventricle; and the third ventricle was associated with pilocytic astrocytoma. The ventricular specificity decreased along the sequence of histopathological dedifferentiation from developmental tumors, WHO grade II-III glioma to glioblastoma. The differences in NT RTD along the ventricular wall may, again, reflect the density of radial units and thus of stem cells (Fig. 8D). The differences between NT entities, however, imply additional intrinsic variability in the differentiation potential of stem cell niches, possibly also related to aging. Stem cell niches are thought to be present in the adult brain – in a similar way to most other organs^34^ – and to be responsible for ongoing neuro- and gliogenesis^35,36^. Such niches were found in the dentate gyrus^37^ or the subventricular zone of the lateral ventricles^38^. A subventricular germinal zone of the third ventricle has been described in children, but appears to be absent in adults^39^. Such periventricular stem cell niches are a likely origin of brain tumors^34,40–42^. The subventricular zone of the lateral ventricles has been associated with glioblastoma^40^, and the germinal zone of the third ventricle with pilocytic astrocytoma^34,42^. In this study, we identified ventriculofugal growth as the dominant growth pattern in NT, consistent with a periventricular origin of primary brain tumors.

We hypothesize that the similarities between the anatomical phenotype of NT and known phylo-ontogenetic patterns are attributable to a neuroepitheloid tumor behavior. RGC-like geno- and cellular phenotypes have been described in NT of different entities, providing a connection between gliomagenesis and both phylo- and ontogenesis^34,42,43^. It is conceivable that RGC-like cells persist into adulthood and serve as tumor-stem cells^22,44^ and migration scaffold, or that cells sharing characteristics of RGCs emerge during gliomagenesis (Fig. 8E)^34,42,43^. PCSNL and metastases, on the other hand, did not exhibit neuroepitheloid behavior. In PCNSL, the identified anatomical pattern along the longitudinal neuraxis and radial axis rather appears to result from an orientation along white matter tracts, ependyma and leptomeninges. Pre- and postoperative growth patterns support this. PCNSL, believed to be of lymphoid origin, seem not to respect the neuroepithelial phylo-ontogenetic units. The anatomical phenotype of metastases along the longitudinal neuraxis is best explained by a seeding in watershed areas of large cerebral vessels. In conjunction with the observed tendency to metastasize at the corticomedullary border, this emphasizes a vascular, especially arterial, anatomical pattern. The predominant growth behavior of metastases was spherical, implying absence of a specific interaction between tumor and brain. Although the power of this study is limited in this respect, the anatomical phenotype of melanoma metastases appeared to be an exception and resembled NT in certain respects. This may be attributable to the neural crest origin of melanocytes.

With progressive histopathological dedifferentiation of NT, a staged deviation from their neuroepitheloid behavior was observed. Along the sequence of WHO grade II-III glioma and glioblastoma, this was demonstrated by a loss of anatomical specificity; phylo- and ontogenetic barriers were increasingly overcome and an increasing frequency of diffuse ependymal or subependymal corpus callosum spread was observed. An involvement of the internal capsule or the central corpus callosum was found in some WHO grade III glioma and glioblastoma. The anatomical tumor behavior may reflect the dedifferentiation of neoplastic stem cells, while the histological entity corresponds to the differentiation capability of neoplastic non-stem cells. A greater degree of dedifferentiation is likely due to a higher number of mutations and the expression of more “atypical”, i.e. non-neuroepithelial, genes by neoplastic stem cells. In hematopoietic neoplasia, oncogenesis has been shown to follow the steps of hematopoiesis in a retrograde way^45–47^. Analogously, it is conceivable that gliomagenesis also follows the steps of neurogliogenesis in a retrograde way^9,10^. In high-grade glioma, molecular subclasses have been identified that resemble the steps of gliomagenesis, correlating with both tumor grade and survival^9,48,49^. Tumors expressing neuronal lineage markers had lower grades and a longer survival than tumors with neuronal stem cell markers^9,48,49^. The expression of mesenchymal markers was linked to shortest survival and tumor recurrence^9,48,49^.

The association between survival probability and anatomical tumor features in this study also reflects such gradual anatomical behavior. There was no difference in survival depending on the white matter sector involved. Although this may seem counterintuitive, it is compatible with neuroepitheloid behavior along the phylo-ontogenetic radial unit. In contrast, diffuse ependymal involvement was associated with an inferior survival compared to focal involvement, explainable with a switch from neuroepitheloid to non-neuroepitheloid anatomical behavior, reflecting an increasingly dedifferentiated state. A subependymal spread along the corpus callosum was associated with survival comparable to that for diffuse subependymal involvement. A lower survival probability was, however, found in the case of invasion of the internal capsule or the central corpus callosum. This behavior might be due to an even more advanced stage of dedifferentiation and continued divergence from neuroepitheloid behavior.

We developed a prototypical staging system for lobar WHO grade II-III glioma and glioblastoma based on the pathophysiological rationale of a stepwise anatomical dedifferentiation. This proved to be accurate across numerous histological, molecular, radiomorphological and clinical strata. A staging system such as the one proposed in this study may inform stage-adapted treatment decisions. Additional studies are needed to validate this staging system, make adjustments, and evaluate its clinical impact. An advantage of such an anatomical staging system over histopathological methods of tumor characterization – which are typically based on single histological samples and thus associated with the risk of sampling bias – is the reflection of the tumor biology in its entirety.

Despite an overall high number of patients, with 1000 included subjects, this study is limited in its statistical power regarding conclusions for rarer tumors. Another intrinsic limitation of this study relates to the partially subjective nature of the assessment of tumor involvement or noninvolvement of anatomical parcellation units. In addition, the effect of volume-normalization should be considered when interpreting the results of the RTP analysis. In some cases, volume-normalization changes the impression drastically compared to absolute or non-volume-normalized RTP.

## Conclusions

In this study, we identified specific anatomical phenotypes in different brain tumor entities through topographic probability and growth pattern assessment. Survival analysis determined prognostically critical steps in the anatomical behavior of neuroepithelial tumors. We developed a prototypical staging system for lobar WHO grade II-III glioma and glioblastoma based on the hypothesized anatomical sequence of tumor progression, which was found to be accurate across histological, molecular, radiomorphological and clinical strata.

## Data Availability

The authors confirm that the data supporting the findings of this study are available within the article and its Supplemental Materials. The statistical code and the anonymized datasets will be made available on GitHub at https://github.com/KevinAkeret/Anatomical-phenotyping-and-staging-of-brain-tumors.

## Acknowledgements

KA is supported by the Prof. Dr. med. Karl und Rena Theiler-Haag foundation. NK is supported by the Swiss National Science Foundation. We thank Miller Rasmy for his contribution to the artistic work.

## Disclosure

None of the authors has any conflict of interest to disclose. All procedures performed in studies involving human participants were in accordance with the ethical standards of the national research committee (Cantonal Ethics Committee Zürich, KEK 01120) and with the 1964 Helsinki declaration and its later amendments.

## Abbreviations

FLAIR: Fluid-attenuated inversion recovery
KPS: Karnofsky Performance Status
MRI: Magnetic resonance imaging
NT: Neuroepithelial tumors
PCNSL: Primary central nervous system lymphoma
RTD: Relative tumor density
RTP: Relative tumor prevalence
SD: Standard deviation

---

**Supplemental Fig. 1.**
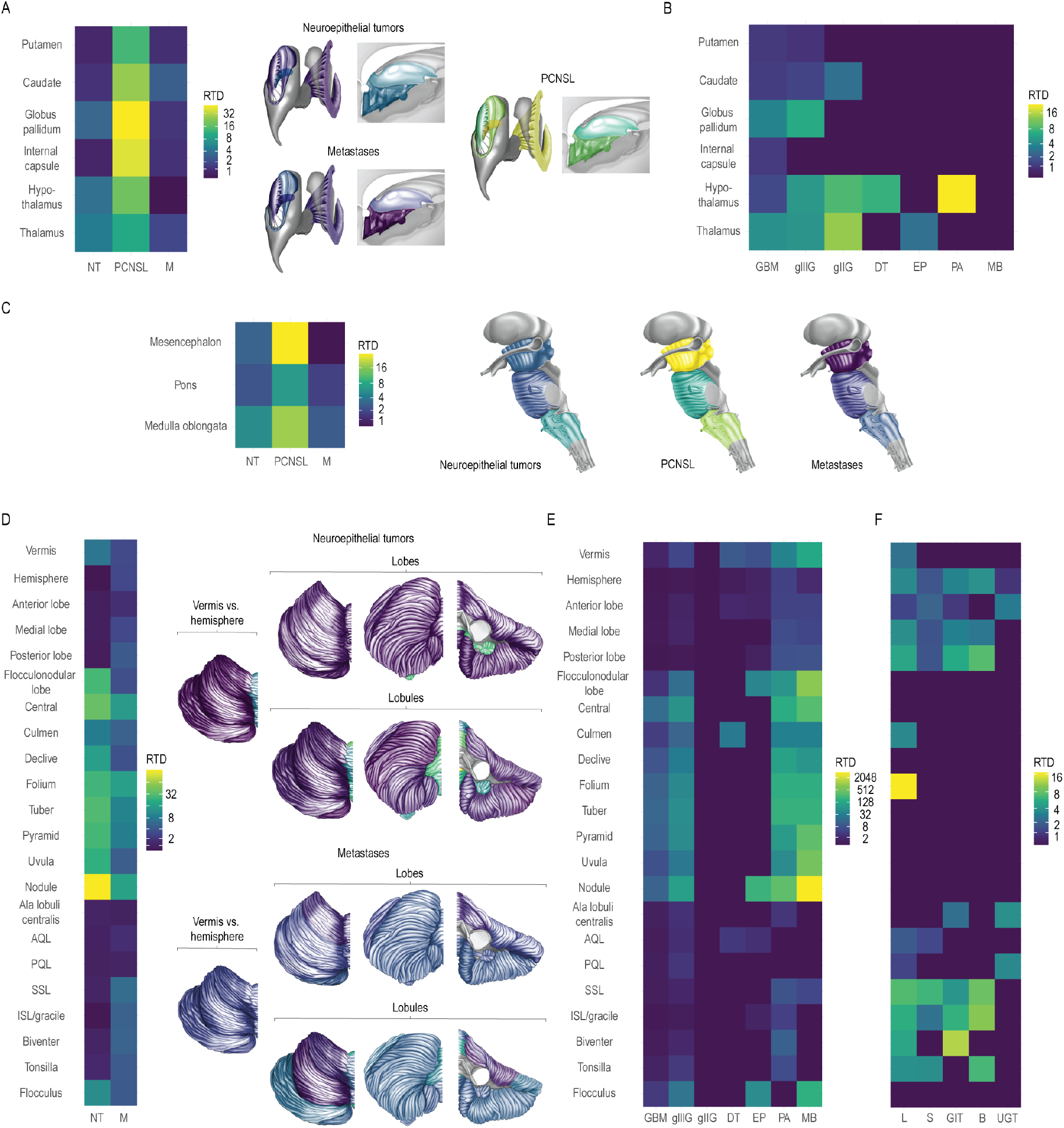
Relative tumor density within central supratentorial and infratentorial parcellation units: **A**. Relative tumor density (RTD) of neuroepithelial tumors (NT) and metastases (M) stratified to their central supratentorial anatomy. **B**. RTD of histopathologic neuroepithelial subtypes stratified to their central supratentorial anatomy. *DT = developmental tumors; EP = ependymoma; gIIG = WHO grade II glioma (astrocytoma, oligoastrocytoma, oligodendroglioma); gIIIG = WHO grade III glioma (astrocytoma, oligoastrocytoma, oligodendroglioma); GBM = glioblastoma; MB = medulloblastoma; PA = pilocytic astrocytoma*. **C**. RTD of NT, PCNSL and M stratified to their brainstem anatomy. **D**. RTD of NT, PCNSL and M stratified to their cerebellar anatomy comparing between vermis and hemisphere, between the individual cerebellar lobes and between the individual cerebellar lobules. *AQL = anterior quadrangular lobule; ISL = inferior semilunar lobule; PQL = posterior quadrangular lobule; SSL = superior semilunar lobule*. **E**. RTD of histopathologic neuroepithelial subtypes stratified to their cerebellar anatomy. **F**. RTD of metastases with respect to the organ of origin stratified to their cerebellar anatomy. *B = breast; GIT = gastrointestinal tract; L = lung; S = skin; UGT = urogenital tract*.

**Supplemental Fig. 2.**
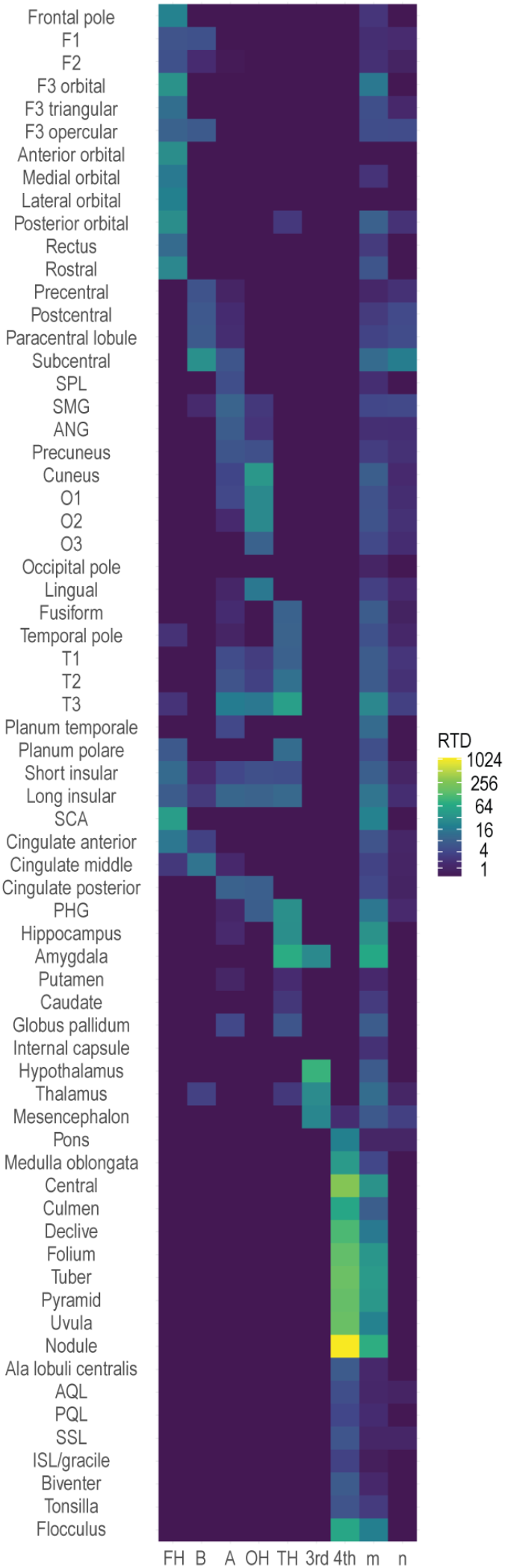
Volume-normalized ventriculo-parenchymal topological relationship in neuroepithelial tumors: Ventriculo-parenchymal topology of neuroepithelial tumors, providing the relative tumor density (RTD) in each anatomical parcellation unit relative to the ventricular contact. *3rd = third ventricle, 4th = fourth ventricle; A = atrium; ANG = angular gyrus; AQL = anterior quadrangular lobule; B = body; F1 = superior frontal gyrus; F2 = middle frontal gyrus; F3 = inferior frontal gyrus; FH = frontal horn; ISL = inferior semilunar lobule; m = multi; n = none; O1 = superior occipital gyrus; O2 = middle occipital gyrus; O3 = inferior occipital gyrus; OH = occipital horn; PHG = parahippocampal gyrus; PQL = posterior quadrangular lobule; RTP = relative tumor prevalence; SCA = subcallosal area; SMG = supramarginal gyrus; SPL = superior parietal lobule; SSL = superior semilunar lobule; T1 = superior temporal gyrus; T2 = middle temporal gyrus; T3 = inferior temporal gyrus*.

**Supplemental Fig. 3.**
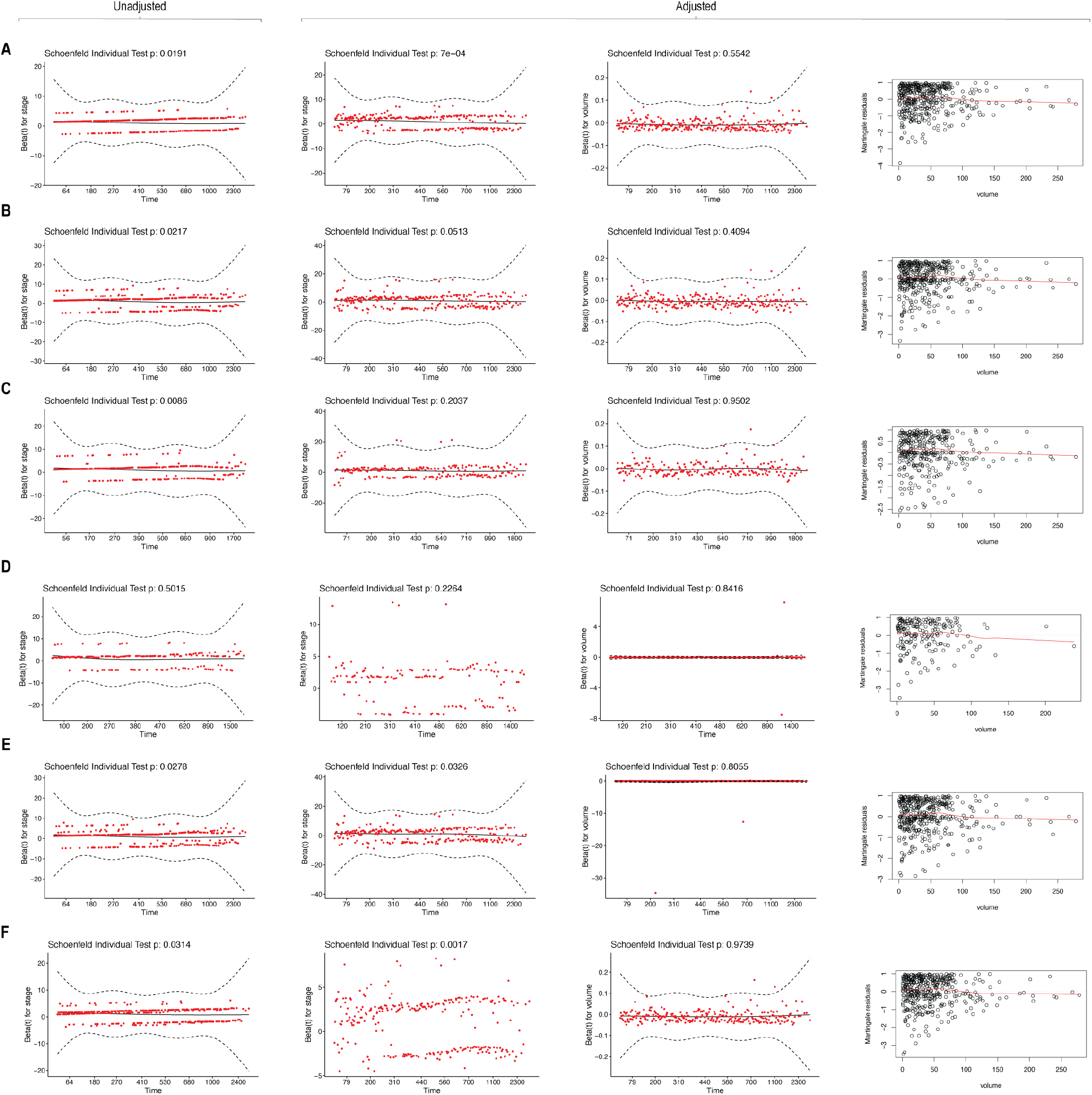
Schoenfeld and Martingale residuals of the multivariable survival analysis: Analysis of the scaled Schoenfeld and Martingale residuals to assess the proportional hazards and the linearity assumptions of the unadjusted and adjusted Cox proportional hazards regression models (Cox PHM) with regard to the proposed anatomical tumor stages. *Unadjusted:* no additional adjustment of the Cox PHM for tumor volume and treatment modality. *Adjusted:* Considering tumor volume as continuous covariable and type of surgery (biopsy or resection), chemotherapy and radiotherapy as binary stratifying variables. **A**. Cox PHM for all neuroepithelial tumors. **B**. Cox PHM considering histological neuroepithelial entity as stratifying variable. **C**. Cox PHM considering IDH1 status as stratifying variable. **D**. Cox PHM considering MGMT methylation status as stratifying variable. **E**. Cox PHM considering contrast behavior as stratifying variable. **F**. Cox PHM considering Karnofsky Performance Score as stratifying variable.

## Notes

### Competing Interest Statement

The authors have declared no competing interest.

### Author Declarations

This study was approved by the ethical review board of the Canton of Zurich, Switzerland (KEK ZH 01120).

## References

1. Patel AP, Fisher JL, Nichols E, et al. Global, regional, and national burden of brain and other CNS cancer, 1990–2016: a systematic analysis for the Global Burden of Disease Study 2016. Lancet Neurol 2019;18(4):376–93.

2. Aldape K, Brindle KM, Chesler L, et al. Challenges to curing primary brain tumours. Nat Rev Clin Oncol 2019;16(8):509–20.

3. Louis DN, Perry A, Reifenberger G, et al. The 2016 World Health Organization Classification of Tumors of the Central Nervous System: a summary. Acta Neuropathol 2016;131(6):803–20.

4. Bates A, Gonzalez-Viana E, Cruickshank G, Roques T. Primary and metastatic brain tumours in adults: summary of NICE guidance. BMJ. 2018;k2924.

5. Brierley J, O’Sullivan B, Asamura H, et al. Global Consultation on Cancer Staging: promoting consistent understanding and use. Nat Rev Clin Oncol 2019;16(12):763–71.

6. Borrell V, Reillo I. Emerging roles of neural stem cells in cerebral cortex development and evolution. Dev Neurobiol 2012;72(7):955–71.

7. Lohmann G, von Cramon DY, Colchester ACF. Deep sulcal landmarks provide an organizing framework for human cortical folding. Cereb Cortex 2008;18(6):1415–20.

8. Lohmann G, von Cramon DY, Steinmetz H. Sulcal variability of twins. Cereb Cortex 1999;9(7):754–63.

9. Phillips HS, Kharbanda S, Chen R, et al. Molecular subclasses of high-grade glioma predict prognosis, delineate a pattern of disease progression, and resemble stages in neurogenesis. Cancer Cell 2006;9(3):157– 73.

10. Verhaak RGW, Hoadley KA, Purdom E, et al. Integrated genomic analysis identifies clinically relevant subtypes of glioblastoma characterized by abnormalities in PDGFRA, IDH1, EGFR, and NF1. Cancer Cell 2010;17(1):98–110.

11. Akeret K, Staartjes VE, Vasella F, et al. Distinct topographic-anatomical patterns in primary and secondary brain tumors and their therapeutic potential. J Neurooncol 2020;149(1):73–85.

12. von Elm E, Altman DG, Egger M, et al. The Strengthening the Reporting of Observational Studies in Epidemiology (STROBE) statement: guidelines for reporting observational studies. Lancet 2007;370(9596):1453–7.

13. Akeret K, van Niftrik B, Seboek M, et al. Topographic volume-standardization atlas of the human brain. medRxiv 2021.

14. Yasargil MG. Microneurosurgery. Vol IVA. Stuttgart, Germany: Georg Thieme; 1994.

15. Kikinis R, Pieper SD, Vosburgh KG. 3D Slicer: A Platform for Subject-Specific Image Analysis, Visualization, and Clinical Support. Intraoperative Imaging and Image-Guided Therapy. 2014;277–89.

16. Kassambara A, Kosinski M, Biecek P, Fabian S. survminer: Drawing Survival Curves using “ggplot2”. 2019. R package version 0 4 2019;4.

17. Grambsch PM, Therneau TM. Modeling survival data: extending the Cox model. Stat Biol Health 2000;

18. Therneau T. A Package for Survival Analysis in R. R package version 3.1--12 (2020). 2020;

19. Bland M. An Introduction to Medical Statistics. Oxford University Press; 2015.

20. Nieuwenhuys R. Histogenesis. In: Nieuwenhuys R, ten Donkelaar HJ, Nicholson C, editors. The Central Nervous System of Vertebrates: Volume 1 / Volume 2 / Volume 3. Berlin, Heidelberg: Springer Berlin Heidelberg; 1998. p. 229–71.

21. Rakic P. Specification of cerebral cortical areas. Science. 1988;241(4862):170–6.

22. Rakic P. Developmental and evolutionary adaptations of cortical radial glia. Cereb Cortex 2003;13(6):541–9.

23. Schmechel DE, Rakic P. A Golgi study of radial glial cells in developing monkey telencephalon: morphogenesis and transformation into astrocytes. Anat Embryol 1979;156(2):115–52.

24. Kuida K, Zheng TS, Na S, et al. Decreased apoptosis in the brain and premature lethality in CPP32-deficient mice. Nature 1996;384(6607):368–72.

25. Chenn A, Walsh CA. Regulation of cerebral cortical size by control of cell cycle exit in neural precursors. Science 2002;297(5580):365–9.

26. Kornack DR, Rakic P. Changes in cell-cycle kinetics during the development and evolution of primate neocortex. Proc Natl Acad Sci U S A 1998;95(3):1242–6.

27. Rakic P. A small step for the cell, a giant leap for mankind: a hypothesis of neocortical expansion during evolution. Trends Neurosci 1995;18(9):383–8.

28. Franco SJ, Gil-Sanz C, Martinez-Garay I, et al. Fate-restricted neural progenitors in the mammalian cerebral cortex. Science 2012;337(6095):746–9.

29. De Juan Romero C, Borrell V. Coevolution of radial glial cells and the cerebral cortex. Glia 2015;63(8):1303– 19.

30. Smart IH, McSherry GM. Gyrus formation in the cerebral cortex of the ferret. II. Description of the internal histological changes. J Anat 1986;147:27–43.

31. Pardal R, Clarke MF, Morrison SJ. Applying the principles of stem-cell biology to cancer. Nat Rev Cancer 2003;3(12):895–902.

32. Nieuwenhuys R. Principles of Current Vertebrate Neuromorphology. Brain Behav Evol 2017;90(2):117–30.

33. Barkovich AJ, Kuzniecky RI, Bollen AW, Grant PE. Focal transmantle dysplasia: a specific malformation of cortical development. Neurology 1997;49(4):1148–52.

34. Tchoghandjian A, Fernandez C, Colin C, et al. Pilocytic astrocytoma of the optic pathway: a tumour deriving from radial glia cells with a specific gene signature. Brain 2009;132(Pt 6):1523–35.

35. Goldman SA, Kirschenbaum B, Harrison-Restelli C, Thaler HT. Neuronal precursors of the adult rat subependymal zone persist into senescence, with no decline in spatial extent or response to BDNF. J Neurobiol 1997;32(6):554–66.

36. Kuhn HG, Dickinson-Anson H, Gage FH. Neurogenesis in the dentate gyrus of the adult rat: age-related decrease of neuronal progenitor proliferation. J Neurosci 1996;16(6):2027–33.

37. Gage FH, Kempermann G, Palmer TD, Peterson DA, Ray J. Multipotent progenitor cells in the adult dentate gyrus. J Neurobiol 1998;36(2):249–66.

38. García-Verdugo Jm, Doetsch F, Wichterle H, Lim DA, Alvarez-Buylla A. Architecture and cell types of the adult subventricular zone: in search of the stem cells. J Neurobiol 1998;36(2):234–48.

39. Dahiya S, Da Yong L, Gutmann DH. Comparative Characterization of the Human and Mouse Third Ventricle Germinal Zones. Journal of Neuropathology & Experimental Neurology. 2011;70(7):622–33.

40. Lee JH, Lee JE, Kahng JY, et al. Human glioblastoma arises from subventricular zone cells with low-level driver mutations. Nature 2018;560(7717):243–7.

41. Lan X, Jörg DJ, Cavalli FMG, et al. Fate mapping of human glioblastoma reveals an invariant stem cell hierarchy. Nature. 2017;549(7671):227–32.

42. Taylor MD, Poppleton H, Fuller C, et al. Radial glia cells are candidate stem cells of ependymoma. Cancer Cell 2005;8(4):323–35.

43. De Rosa A, Pellegatta S, Rossi M, et al. A radial glia gene marker, fatty acid binding protein 7 (FABP7), is involved in proliferation and invasion of glioblastoma cells. PLoS One 2012;7(12):e52113.

44. Barry DS, Pakan JMP, McDermott KW. Radial glial cells: key organisers in CNS development. Int J Biochem Cell Biol 2014;46:76–9.

45. Bonnet D, Dick JE. Human acute myeloid leukemia is organized as a hierarchy that originates from a primitive hematopoietic cell. Nat Med 1997;3(7):730–7.

46. Lapidot T, Sirard C, Vormoor J, et al. A cell initiating human acute myeloid leukaemia after transplantation into SCID mice. Nature 1994;367(6464):645–8.

47. Castor A, Nilsson L, Astrand-Grundström I, et al. Distinct patterns of hematopoietic stem cell involvement in acute lymphoblastic leukemia. Nat Med 2005;11(6):630–7.

48. Tso C-L, Shintaku P, Chen J, et al. Primary glioblastomas express mesenchymal stem-like properties. Mol Cancer Res 2006;4(9):607–19.

49. Tso C-L, Freije WA, Day A, et al. Distinct transcription profiles of primary and secondary glioblastoma subgroups. Cancer Res 2006;66(1):159–67.

