## Supplemental Table for "Anatomical phenotyping and staging of brain tumors"

### Supplemental Table 1. Acquired demographic, clinical and histopathologic data

The acquired demographic, clinical and Histopathologic parameters with the respective specification.

| ACQUIRED PARAMETERS | DEFINITION/EXPLANATION |
|---|---|
| **Demographic** | |
| Age | At time of surgery (in years) |
| Sex | Female, male |
| **Clinical** | |
| Karnofsky Performance Status (KPS) | At time of presurgical hospital admission |
| Type of surgery | Resection or biopsy |
| Adjuvant therapy | Radiotherapy and/or chemotherapy |
| Overall survival (OS) | Time (in days) from surgery until death or last follow-up |
| **Histopathologic** | |
| Histological entity | Until 2016 according to the 2007 WHO Classification[1] of Tumours of the Central Nervous System, since 2016 according to the 2016 update[2] |
| WHO grade | Only with neuroepithelial tumors. Until 2016 according to the 2007 WHO Classification[1] of Tumours of the Central Nervous System, since 2016 according to the 2016 update[2]. |

### Supplemental Table 2. Phylogenetic and ontogenetic parcellation protocol

The individual topographic parcellation units[3] are listed with the corresponding relative volume (in relation to the total individual encephalic volume)[3] and its phylogenetic and ontogenetic affiliation[4–10]. *AC/PC = Archi- and Paleocerebellum; AP/PP = Archi- and Paleopallium; DPa = dorsal pallium; H = hypothalamus; LPa/VPa = lateral/ventral pallium; M = mesencephalon; MsP = Mesopallium; MdP = medial pallium; NC = Neocerebellum; NP = Neopallium; R = rhombencephalon; SH = Stammhirn; SPa = Subpallium; T = thalamus.*

| TOPOGRAPHIC PARCELLATION UNIT | RELATIVE VOLUME | PHYLOGENY | ONTOGENY |
|---|---|---|---|
| **Cerebral gyral segments** | | | |
| Frontal pole | 0.430816128 | NP | DPa |
| Superior frontal gyrus | 6.279798428 | NP | DPa |
| Middle frontal gyrus | 5.337649747 | NP | DPa |
| Inferior frontal gyrus, orbital part | 0.358035003 | NP | DPa |
| Inferior frontal gyrus, triangular part | 1.071281027 | NP | DPa |
| Inferior frontal gyrus, opercular part | 1.278079325 | NP | DPa |
| Anterior orbital gyrus | 0.305823941 | NP | DPa |
| Medial orbital gyrus | 0.690815598 | NP | DPa |
| Lateral orbital gyrus | 0.553600509 | NP | DPa |
| Posterior orbital gyrus | 0.677565662 | MsP | LVPa |
| Gyrus rectus | 0.913437544 | NP | DPa |
| Rostral gyrus | 0.329349815 | NP | DPa |
| Subcallosal area | 0.217079725 | AP/PP | DPa |
| Precentral gyrus | 3.587020729 | NP | DPa |
| Postcentral gyrus | 2.037574286 | NP | DPa |
| Paracentral lobule | 1.287248954 | NP | DPa |
| Subcentral gyrus | 0.5533886 | NP | DPa |
| Superior parietal lobule | 3.051229984 | NP | DPa |
| Supramarginal gyrus | 2.805470234 | NP | DPa |
| Angular gyrus | 3.140150843 | NP | DPa |
| Precuneus | 2.818478929 | NP | DPa |
| Cuneus | 0.921807593 | NP | DPa |
| Superior occipital gyrus | 0.836334771 | NP | DPa |
| Middle occipital gyrus | 1.441203685 | NP | DPa |
| Inferior occipital gyrus | 0.845945253 | NP | DPa |
| Occipital pole | 1.040137147 | NP | DPa |
| Lingual gyrus | 1.861172912 | NP | DPa |
| Fusiform gyrus | 2.031391571 | NP | DPa |
| Superior temporal gyrus | 2.489971123 | NP | DPa |

| | | | |
|---|---|---|---|
| Middle temporal gyrus | 2.171034008 | NP | DPa |
| Inferior temporal gyrus | 0.457353845 | NP | DPa |
| Planum temporale | 0.280539509 | NP | DPa |
| Planum polare | 0.911025095 | MsP | LVPa |
| Temporal pole | 2.316330704 | MsP | DPa |
| Short insular gyri | 1.414900052 | MsP | LVPa |
| Long insular gyri | 0.832453891 | NP | DPa |
| Parahippocampal gyrus | 0.956311824 | MsP | LVPa |
| Amygdala | 0.286452663 | AP/PP | SPa; LVPa |
| Hippocampus | 0.744687497 | AP/PP | MdP |
| Cingulate anterior | 1.317694878 | MsP | DPa |
| Cingulate middle | 1.563786755 | MsP | DPa |
| Cingulate posterior | 1.841331053 | MsP | DPa |
| **Central supratentorial structures** | | | |
| Caudate nucleus | 0.713849498 | SH | SPa |
| Putamen | 1.034494995 | SH | SPa |
| Globus pallidum | 0.280506077 | SH | SPa |
| Claustrum | 0.122849802 | MsP | LVPa |
| Hypothalamus | 0.728936893 | SH | H |
| Thalamus | 1.340071726 | SH | T |
| Innominate substance | 0.247721762 | - | SPa |
| **Brainstem** | | | |
| Mesencephalon | 0.920762329 | SH | M |
| Pons | 1.400562507 | SH | R |
| Medulla oblongata | 0.252406131 | SH | R |
| **Vermian lobules** | | | |
| Central | 0.044497901 | AC/PC | R |
| Culmen | 0.200655643 | AC/PC | R |
| Declive | 0.094555295 | NC | R |
| Folium | 0.04051762 | NC | R |
| Tuber | 0.035534754 | NC | R |
| Pyramid | 0.038985895 | AC/PC | R |
| Uvula | 0.072222968 | AC/PC | R |
| Nodule | 0.019623856 | AC/PC | R |
| **Cerebellar hemispheric lobules** | | | |
| Ala lobuli centralis | 1.104029712 | AC/PC | R |
| Anterior quadrangular lobule | 1.621911333 | AC/PC | R |
| Posterior quadrangular lobule | 1.202306311 | NC | R |
| Superior semilunar lobule | 1.304405853 | NC | R |
| Inferior semilunar / gracile lobule | 3.220567307 | NC | R |

| | | | |
|---|---|---|---|
| Biventer lobule | 1.107047752 | NC | R |
| Tonsilla | 0.546270333 | NC | R |
| Flocculus | 0.078694963 | AC/PC | R |

### Supplemental Table 3. Volumes of the phylogenetic and ontogenetic parcellation units

The individual phylo- and ontogenetic parcellation units are listed with the corresponding relative volume (in relation to the total individual encephalic volume).

| ANATOMICAL UNIT | RELATIVE VOLUME |
|---|---|
| **Phylogeny** | |
| **Neopallium** | 52.03859493 |
| **Mesopallium** | 11.12179583 |
| **Achi-/Paleopallium** | 1.248219885 |
| **Stammhirn** | 6.671590156 |
| **Neocerebellum** | 7.551205226 |
| **Archi-/Paleocerebellum** | 3.180622271 |
| **Ontogeny** | |
| **Dorsal pallium** | 59.29481805 |
| **Lateral/ventral pallium** | 4.3691051 |
| **Medial pallium** | 0.744687497 |
| **Subpallium** | 2.563024996 |
| **Hypothalamus** | 0.728936893 |
| **Thalamus** | 1.340071726 |
| **Mesencephalon** | 0.920762329 |
| **Rhombencephalon** | 12.30610117 |

### Supplemental Table 4. Histopathology

**A.** Main histopathologic results of the study population. *PCNSL = primary central nervous system lymphoma.* **B.** Details on the histopathologic subtypes identified in the study population.

| SUPPLEMENTAL TABLE 4A | n (%) / mean (SD) |
|---|---|
| **Total** | 1000 |
| **Metastases** | 299 (29.9) |
| **Neuroepithelial tumor** | 657 (65.7) |
| **PCNSL** | 44 (4.4) |
| **Neuroepithelial subentities (main)** | |
| **Glioblastoma** | 385 (60.6) |
| **WHO grade III glioma** (astrocytoma, oligoastrocytoma, oligodendroglioma) | 110 (17.3) |
| **WHO grade II glioma** (astrocytoma, oligoastrocytoma, oligodendroglioma) | 50 (7.9) |
| **Pilocytic astrocytoma** | 30 (4.7) |
| **Ependymoma** | 26 (4.1) |
| **Developmental tumors** | 19 (3.0) |
| **Medulloblastoma** | 15 (2.4) |
| **WHO grade** | |
| **I** | 56 (5.6) |
| **II** | 70 (7.0) |
| **III** | 128 (12.8) |
| **IV** | 402 (40.2) |
| **none** | 344 (34.4) |
| **IDH1 mutation** | 89 (19.5) |
| **MGMT promoter methylation** | 97 (39.0) |
| **Metastases subentities (main)** | |
| **Lung** | 143 (47.8) |
| **Melanoma** | 49 (16.4) |
| **Gastrointestinal tract*** | 37 (12.4) |
| **Breast** | 28 (9.4) |
| **Urogenital tract**** | 24 (8.0) |
| **Miscellaneous***** | 18 (6.0) |

*Mouth, tonsil, parotid, esophagus, stomach, gallbladder, pancreas, colorectal cancer.
**Kidney, bladder; ovary, tube, uterus; testes, prostate.
***Cancer of unknown primary, adrenal, leukemia, sarcoma, mesothelial, thyroid.

| SUPPLEMENTAL TABLE 4B | n (%) |
|---|---|
| Adrenocortical carcinoma metastases | 1 (0.1) |
| Astrocytoma | 99 (9.9) |
| AT/RT (atypical teratoid rhabdoid tumor) | 1 (0.1) |
| Bronchus carcinoma metastases (adenocarcinoma) | 93 (9.3) |
| Bronchus carcinoma metastases (carcinosarcoma) | 1 (0.1) |
| Bronchus carcinoma metastases (combined small cell lung cancer) | 3 (0.3) |
| Bronchus carcinoma metastases (large cell lung cancer) | 8 (0.8) |
| Bronchus carcinoma metastases (non small cell lung cancer NOD) | 3 (0.3) |
| Bronchus carcinoma metastases (small cell lung cancer) | 18 (1.8) |
| Bronchus carcinoma metastases (squamous cell carcinoma) | 17 (1.7) |
| Central neurocytoma | 3 (0.3) |
| CML (chronic myeloid leukemia) metastases | 1 (0.1) |
| CRC (colorectal cancer) metastases | 21 (2.1) |
| CUP (cancer of unknown primary) | 4 (0.4) |
| DNET (dysembryoplastic neuroepithelial tumor) | 4 (0.4) |
| Ependymoma | 26 (2.6) |
| Esophageal carcinoma metastases | 6 (0.6) |
| Gall bladder cancer | 1 (0.1) |
| Ganglioglioma | 15 (1.5) |
| Gastric carcinoma metastases | 2 (0.2) |
| Glioblastoma | 385 (38.5) |
| Germ cell cancer metastases | 6 (0.6) |
| Leiyomyosarcoma metastases | 1 (0.1) |
| Lymphoma | 47 (4.7) |
| Mamma carcinoma metastases | 28 (2.8) |
| Medulloblastoma | 15 (1.5) |
| Melanoma metastases | 49 (4.9) |
| Myxofibrosarcoma metastases | 1 (0.1) |
| Neuroendocrine carcinoma metastases | 2 (0.2) |
| Oligoastrocytoma | 22 (2.2) |
| Oligodendroglioma | 39 (3.9) |
| Oral carcinoma metastases | 3 (0.3) |
| Ovarian carcinoma metastases | 1 (0.1) |
| Pancreas carcinoma metastases | 1 (0.1) |
| Parotid carcinoma metastases | 2 (0.2) |
| Peritoneal carcinoma metastases | 1 (0.1) |
| Pilocytic astrocytoma | 30 (3.0) |
| Pilomyxoid astrocytoma | 4 (0.4) |
| Pleomorphic xanthastrocytoma | 2 (0.2) |
| Pleural mesiothelioma metastases | 1 (0.1) |
| Plexus papilloma | 5 (0.5) |
| PNET (primitive neuroectodermal tumors) | 1 (0.1) |
| Prostate carcinoma metastases | 3 (0.3) |

| | |
|---|---|
| **Renal cell cancer metastases** | 8 (0.8) |
| **RGNT (rosette-forming glioneuronal tumor)** | 1 (0.1) |
| **Phabdomyosarcoma metastases** | 1 (0.1) |
| **Subependymal giant cell astrocytoma** | 1 (0.1) |
| **Subependymoma** | 4 (0.4) |
| **Thyroid carcinoma metastases** | 2 (0.2) |
| **Tonsillar squamous cell carcinoma metastases** | 1 (0.1) |
| **Tubal carcinoma** | 1 (0.1) |
| **Urothelial carcinoma metastases** | 4 (0.4) |
| **Uterus carcinoma metastases** | 1 (0.1) |

### Supplemental Table 5. Karnofsky Performance Status

Karnofsky Performance Status (KPS) at the time of surgery.

| KPS | n (%) |
|---|---|
| **100** | 38 (4.0) |
| **90** | 307 (31.9) |
| **80** | 301 (31.3) |
| **70** | 173 (18.0) |
| **60** | 90 (9.4) |
| **50** | 42 (4.4) |
| **40** | 9 (0.9) |
| **20** | 1 (0.1) |

**Supplemental Table 6. General anatomical patterns of neuroepithelial tumors, primary central nervous system lymphoma and brain metastases**

The involvement of general topographic, phylogenetic and ontogenetic parcellation units by brain tumors in total, neuroepithelial tumors (NT), primary central nervous system lymphoma (PCNSL) or metastases is given in absolute numbers (n), relative tumor prevalence (RTP) and relative tumor density (RTD). *AC/PC = Archi- and Paleocerebellum; AP/PP = Archi- and Paleopallium; DPa = dorsal pallium; H = hypothalamus; LPa/VPa = lateral/ventral pallium; M = mesencephalon; MsP = Mesopallium; MdP = medial pallium; NC = Neocerebellum; NP = Neopallium; R = rhombencephalon; SH = Stammhirn; SPa = Subpallium; T = thalamus.*

| | Total (n, RTP [95% CI], RTD [95% CI]) | NT (n, RTP [95% CI], RTD [95% CI]) | PCNSL (n, RTP [95% CI], RTD [95% CI]) | Metastases (n, RTP [95% CI], RTD [95% CI]) |
|---|---|---|---|---|
| **Total** | 1000, 100 (99.6 to 100), 1 (1 to 1) | 657, 100 (99.4 to 100), 1 (1 to 1) | 44, 100 (92 to 100), 1 (0.9 to 1) | 299, 100 (98.7 to 100), 1 (1 to 1) |
| **TOPOGRAPHY** | | | | |
| **Frontal lobe** | 316, 31.6 (28.8 to 34.5), 1.7 (1.6 to 1.9) | 186, 28.3 (25 to 31.9), 1.6 (1.4 to 1.7) | 22, 50 (35.8 to 64.2), 2.7 (2 to 3.5) | 108, 36.1 (30.9 to 41.7), 2 (1.7 to 2.3) |
| **Central lobe** | 159, 15.9 (13.8 to 18.3), 2.1 (1.8 to 2.5) | 82, 12.5 (10.2 to 15.2), 1.7 (1.4 to 2) | 14, 31.8 (20 to 46.6), 4.3 (2.7 to 6.2) | 63, 21.1 (16.8 to 26), 2.8 (2.3 to 3.5) |
| **Parietal lobe** | 204, 20.4 (18 to 23), 1.7 (1.5 to 1.9) | 125, 19 (16.2 to 22.2), 1.6 (1.4 to 1.9) | 16, 36.4 (23.8 to 51.1), 3.1 (2 to 4.3) | 63, 21.1 (16.8 to 26), 1.8 (1.4 to 2.2) |
| **Occipital lobe** | 156, 15.6 (13.5 to 18), 2.2 (1.9 to 2.6) | 76, 11.6 (9.3 to 14.2), 1.7 (1.3 to 2) | 12, 27.3 (16.3 to 41.8), 3.9 (2.4 to 6) | 68, 22.7 (18.4 to 27.8), 3.3 (2.6 to 4) |
| **Temporal lobe** | 244, 24.4 (21.8 to 27.2), 2.3 (2 to 2.5) | 186, 28.3 (25 to 31.9), 2.7 (2.3 to 3) | 9, 20.5 (11.2 to 34.5), 1.9 (1 to 3.2) | 49, 16.4 (12.6 to 21), 1.5 (1.2 to 2) |
| **Insular lobe** | 83, 8.3 (6.7 to 10.2), 3.7 (3 to 4.5) | 68, 10.4 (8.2 to 12.9), 4.6 (3.7 to 5.7) | 8, 18.2 (9.5 to 32), 8.1 (4.2 to 14.2) | 7, 2.3 (1.1 to 4.8), 1 (0.5 to 2.1) |
| **Limbic lobe** | 195, 19.5 (17.2 to 22.1), 2.8 (2.5 to 3.2) | 160, 24.4 (21.2 to 27.8), 3.5 (3.1 to 4) | 14, 31.8 (20 to 46.6), 4.6 (2.9 to 6.7) | 21, 7 (4.6 to 10.5), 1 (0.7 to 1.5) |
| **Basal ganglia** | 24, 2.4 (1.6 to 3.5), 1.2 (0.8 to 1.7) | 8, 1.2 (0.6 to 2.4), 0.6 (0.3 to 1.2) | 8, 18.2 (9.5 to 32), 9 (4.7 to 15.8) | 8, 2.7 (1.4 to 5.2), 1.3 (0.7 to 2.6) |
| **Diencephalon** | 71, 7.1 (5.7 to 8.9), 3.4 (2.7 to 4.3) | 55, 8.4 (6.5 to 10.7), 4 (3.1 to 5.2) | 9, 20.5 (11.2 to 34.5), 9.9 (5.4 to 16.7) | 7, 2.3 (1.1 to 4.8), 1.1 (0.6 to 2.3) |
| **Brainstem** | 58, 5.8 (4.5 to 7.4), 2.3 (1.8 to 2.9) | 38, 5.8 (4.2 to 7.8), 2.2 (1.6 to 3) | 12, 27.3 (16.3 to 41.8), 10.6 (6.4 to 16.3) | 8, 2.7 (1.4 to 5.2), 1 (0.5 to 2) |
| **Cerebellum** | 136, 13.6 (11.6 to 15.9), 1.3 (1.1 to 1.5) | 43, 6.5 (4.9 to 8.7), 0.6 (0.5 to 0.8) | 5, 11.4 (5 to 24), 1.1 (0.5 to 2.2) | 88, 29.4 (24.6 to 34.8), 2.7 (2.3 to 3.2) |
| **PHYLOGENY** | | | | |
| **NP** | 707, 70.7 (67.8 to 73.4), 1.4 (1.3 to 1.4) | 463, 70.5 (66.9 to 73.8), 1.4 (1.3 to 1.4) | 19, 43.2 (29.7 to 57.8), 0.8 (0.6 to 1.1) | 225, 75.3 (70.1 to 79.8), 1.4 (1.3 to 1.5) |
| **MsP** | 218, 21.8 (19.4 to 24.5), 2 (1.7 to 2.2) | 187, 28.5 (25.1 to 32), 2.6 (2.3 to 2.9) | 10, 22.7 (12.8 to 37), 2 (1.2 to 3.3) | 21, 7 (4.6 to 10.5), 0.6 (0.4 to 0.9) |
| **AP/PP** | 91, 9.1 (7.5 to 11), 7.3 (6 to 8.8) | 82, 12.5 (10.2 to 15.2), 10 (8.1 to 12.2) | 5, 11.4 (5 to 24), 9.1 (4 to 19.2) | 4, 1.3 (0.5 to 3.4), 1.1 (0.4 to 2.7) |
| **SH** | 121, 12.1 (10.2 to 14.3), 1.8 (1.5 to 2.1) | 84, 12.8 (10.4 to 15.6), 1.9 (1.6 to 2.3) | 16, 36.4 (23.8 to 51.1), 5.5 (3.6 to 7.7) | 21, 7 (4.6 to 10.5), 1.1 (0.7 to 1.6) |
| **NC** | 114, 11.4 (9.6 to 13.5), 1.5 (1.3 to 1.8) | 26, 4 (2.7 to 5.7), 0.5 (0.4 to 0.8) | 5, 11.4 (5 to 24), 1.5 (0.7 to 3.2) | 83, 27.8 (23 to 33.1), 3.7 (3 to 4.4) |
| **AC/PC** | 42, 4.2 (3.1 to 5.6), 1.3 (1 to 1.8) | 28, 4.3 (3 to 6.1), 1.3 (0.9 to 1.9) | 2, 4.5 (1.3 to 15.1), 1.4 (0.4 to 4.8) | 12, 4 (2.3 to 6.9), 1.3 (0.7 to 2.2) |
| **ONTOGENY** | | | | |
| **DP** | 754, 75.4 (72.6 to 78), 1.3 (1.2 to 1.3) | 502, 76.4 (73 to 79.5), 1.3 (1.2 to 1.3) | 20, 45.5 (31.7 to 59.9), 0.8 (0.5 to 1) | 232, 77.6 (72.5 to 82), 1.3 (1.2 to 1.4) |
| **LPa/VPa** | 138, 13.8 (11.8 to 16.1), 3.2 (2.7 to 3.7) | 119, 18.1 (15.4 to 21.2), 4.1 (3.5 to 4.9) | 9, 20.5 (11.2 to 34.5), 4.7 (2.6 to 7.9) | 10, 3.3 (1.8 to 6), 0.8 (0.4 to 1.4) |
| **MdP** | 69, 6.9 (5.5 to 8.6), 9.3 (7.4 to 11.6) | 63, 9.6 (7.6 to 12.1), 12.9 (10.2 to 16.2) | 3, 6.8 (2.3 to 18.2), 9.2 (3.2 to 24.5) | 3, 1 (0.3 to 2.9), 1.3 (0.5 to 3.9) |
| **SPa** | 93, 9.3 (7.7 to 11.3), 3.6 (3 to 4.4) | 71, 10.8 (8.7 to 13.4), 4.2 (3.4 to 5.2) | 12, 27.3 (16.3 to 41.8), 10.6 (6.4 to 16.3) | 10, 3.3 (1.8 to 6), 1.3 (0.7 to 2.4) |
| **H** | 25, 2.5 (1.7 to 3.7), 3.4 (2.3 to 5) | 19, 2.9 (1.9 to 4.5), 4 (2.6 to 6.1) | 6, 13.6 (6.4 to 26.7), 18.7 (8.8 to 36.6) | 0, 0 (0 to 1.3), 0 (0 to 1.7) |
| **T** | 54, 5.4 (4.2 to 7), 4 (3.1 to 5.2) | 41, 6.2 (4.6 to 8.4), 4.7 (3.5 to 6.2) | 6, 13.6 (6.4 to 26.7), 10.2 (4.8 to 19.9) | 7, 2.3 (1.1 to 4.8), 1.7 (0.8 to 3.5) |
| **M** | 31, 3.1 (2.2 to 4.4), 3.4 (2.4 to 4.7) | 18, 2.7 (1.7 to 4.3), 3 (1.9 to 4.7) | 12, 27.3 (16.3 to 41.8), 29.6 (17.8 to 45.5) | 1, 0.3 (0.1 to 1.9), 0.4 (0.1 to 2) |
| **R** | 164, 16.4 (14.2 to 18.8), 1.3 (1.2 to 1.5) | 62, 9.4 (7.4 to 11.9), 0.8 (0.6 to 1) | 8, 18.2 (9.5 to 32), 1.5 (0.8 to 2.6) | 94, 31.4 (26.4 to 36.9), 2.6 (2.1 to 3) |

**Supplemental Table 7. General anatomical patterns of neuroepithelial tumor subentities**

The involvement of general topographic, phylogenetic and ontogenetic parcellation units by neuroepithelial tumors (NT) in total, glioblastoma (GBM), WHO grade III (gIIIG) and II (gIIG) glioma, developmental tumors (DT), pilocytic astrocytoma (PA), ependymoma (EP) or medulloblastoma (MB) is given in absolute numbers (n), relative tumor prevalence (RTP) and relative tumor density (RTD). *AC/PC = Archi- and Paleocerebellum; AP/PP = Archi- and Paleopallium; DPa = dorsal pallium; H = hypothalamus; LPa/VPa = lateral/ventral pallium; M = mesencephalon; MsP = Mesopallium; MdP = medial pallium; NC = Neocerebellum; NP = Neopallium; R = rhombencephalon; SH = Stammhirn; SPa = Subpallium; T = thalamus.*

| | NT total (n, RTP [95% CI], RTD [95% CI]) | GBM (n, RTP [95% CI], RTD [95% CI]) | gIIIG (n, RTP [95% CI], RTD [95% CI]) | gIIG (n, RTP [95% CI], RTD [95% CI]) | DT (n, RTP [95% CI], RTD [95% CI]) | PA (n, RTP [95% CI], RTD [95% CI]) | EP (n, RTP [95% CI], RTD [95% CI]) | MB (n, RTP [95% CI], RTD [95% CI]) |
|---|---|---|---|---|---|---|---|---|
| **Total** | 635, 100 (99.4 to 100), 1 (1 to 1) | 385, 100 (99 to 100), 1 (1 to 1) | 110, 100 (96.6 to 100), 1 (1 to 1) | 50, 100 (92.9 to 100), 1 (0.9 to 1) | 19, 100 (83.2 to 100), 1 (0.8 to 1) | 30, 100 (88.6 to 100), 1 (0.9 to 1) | 26, 100 (87.1 to 100), 1 (0.9 to 1) | 15, 100 (79.6 to 100), 1 (0.8 to 1) |
| **TOPOGRAPHY** | | | | | | | | |
| **Frontal lobe** | 185, 29.1 (25.7 to 32.8), 1.6 (1.4 to 1.8) | 108, 28.1 (23.8 to 32.7), 1.5 (1.3 to 1.8) | 51, 46.4 (37.3 to 55.6), 2.5 (2 to 3.1) | 25, 50 (36.6 to 63.4), 2.7 (2 to 3.5) | 1, 5.3 (0.9 to 24.6), 0.3 (0.1 to 1.4) | 0, 0 (0 to 11.4), 0 (0 to 0.6) | 0, 0 (0 to 12.9), 0 (0 to 0.7) | 0, 0 (0 to 20.4), 0 (0 to 1.1) |
| **Central lobe** | 82, 12.9 (10.5 to 15.7), 1.7 (1.4 to 2.1) | 51, 13.2 (10.2 to 17), 1.8 (1.4 to 2.3) | 21, 19.1 (12.8 to 27.4), 2.6 (1.7 to 3.7) | 8, 16 (8.3 to 28.5), 2.1 (1.1 to 3.8) | 0, 0 (0 to 16.8), 0 (0 to 2.3) | 0, 0 (0 to 11.4), 0 (0 to 1.5) | 2, 7.7 (2.1 to 24.1), 1 (0.3 to 3.2) | 0, 0 (0 to 20.4), 0 (0 to 2.7) |
| **Parietal lobe** | 125, 19.7 (16.8 to 23), 1.7 (1.4 to 1.9) | 95, 24.7 (20.6 to 29.2), 2.1 (1.7 to 2.5) | 19, 17.3 (11.3 to 25.4), 1.5 (1 to 2.2) | 5, 10 (4.3 to 21.4), 0.8 (0.4 to 1.8) | 4, 21.1 (8.5 to 43.3), 1.8 (0.7 to 3.7) | 1, 3.3 (0.6 to 16.7), 0.3 (0.1 to 1.4) | 1, 3.8 (0.7 to 18.9), 0.3 (0.1 to 1.6) | 0, 0 (0 to 20.4), 0 (0 to 1.7) |
| **Occipital lobe** | 75, 11.8 (9.5 to 14.6), 1.7 (1.4 to 2.1) | 64, 16.6 (13.2 to 20.7), 2.4 (1.9 to 3) | 8, 7.3 (3.7 to 13.7), 1 (0.5 to 2) | 1, 2 (0.4 to 10.5), 0.3 (0.1 to 1.5) | 2, 10.5 (2.9 to 31.4), 1.5 (0.4 to 4.5) | 0, 0 (0 to 11.4), 0 (0 to 1.6) | 0, 0 (0 to 12.9), 0 (0 to 1.9) | 0, 0 (0 to 20.4), 0 (0 to 2.9) |
| **Temporal lobe** | 184, 29 (25.6 to 32.6), 2.7 (2.4 to 3.1) | 136, 35.3 (30.7 to 40.2), 3.3 (2.9 to 3.8) | 29, 26.4 (19 to 35.3), 2.5 (1.8 to 3.3) | 10, 20 (11.2 to 33), 1.9 (1.1 to 3.1) | 5, 26.3 (11.8 to 48.8), 2.5 (1.1 to 4.6) | 0, 0 (0 to 11.4), 0 (0 to 1.1) | 4, 15.4 (6.2 to 33.5), 1.4 (0.6 to 3.1) | 0, 0 (0 to 20.4), 0 (0 to 1.9) |
| **Insular lobe** | 68, 10.7 (8.5 to 13.4), 4.8 (3.8 to 5.9) | 35, 9.1 (6.6 to 12.4), 4 (2.9 to 5.5) | 21, 19.1 (12.8 to 27.4), 8.5 (5.7 to 12.2) | 12, 24 (14.3 to 37.4), 10.7 (6.4 to 16.6) | 0, 0 (0 to 16.8), 0 (0 to 7.5) | 0, 0 (0 to 11.4), 0 (0 to 5.1) | 0, 0 (0 to 12.9), 0 (0 to 5.7) | 0, 0 (0 to 20.4), 0 (0 to 9.1) |
| **Limbic lobe** | 160, 25.2 (22 to 28.7), 3.6 (3.2 to 4.1) | 98, 25.5 (21.4 to 30), 3.7 (3.1 to 4.3) | 39, 35.5 (27.1 to 44.7), 5.1 (3.9 to 6.5) | 16, 32 (20.8 to 45.8), 4.6 (3 to 6.6) | 6, 31.6 (15.4 to 54), 4.6 (2.2 to 7.8) | 1, 3.3 (0.6 to 16.7), 0.5 (0.1 to 2.4) | 0, 0 (0 to 12.9), 0 (0 to 1.9) | 0, 0 (0 to 20.4), 0 (0 to 2.9) |
| **Basal ganglia** | 8, 1.3 (0.6 to 2.5), 0.6 (0.3 to 1.2) | 5, 1.3 (0.6 to 3), 0.6 (0.3 to 1.5) | 2, 1.8 (0.5 to 6.4), 0.9 (0.2 to 3.1) | 1, 2 (0.4 to 10.5), 1 (0.2 to 5.2) | 0, 0 (0 to 16.8), 0 (0 to 8.3) | 0, 0 (0 to 11.4), 0 (0 to 5.6) | 0, 0 (0 to 12.9), 0 (0 to 6.3) | 0, 0 (0 to 20.4), 0 (0 to 10) |
| **Diencephalon** | 52, 8.2 (6.3 to 10.6), 4 (3 to 5.1) | 26, 6.8 (4.6 to 9.7), 3.3 (2.2 to 4.7) | 10, 9.1 (5 to 15.9), 4.4 (2.4 to 7.7) | 9, 18 (9.8 to 30.8), 8.7 (4.7 to 14.9) | 1, 5.3 (0.9 to 24.6), 2.5 (0.5 to 11.9) | 5, 16.7 (7.3 to 33.6), 8.1 (3.5 to 16.2) | 1, 3.8 (0.7 to 18.9), 1.9 (0.3 to 9.1) | 0, 0 (0 to 20.4), 0 (0 to 9.9) |
| **Brainstem** | 36, 5.7 (4.1 to 7.7), 2.2 (1.6 to 3) | 7, 1.8 (0.9 to 3.7), 0.7 (0.3 to 1.4) | 11, 10 (5.7 to 17), 3.9 (2.2 to 6.6) | 4, 8 (3.2 to 18.8), 3.1 (1.2 to 7.3) | 1, 5.3 (0.9 to 24.6), 2 (0.4 to 9.6) | 5, 16.7 (7.3 to 33.6), 6.5 (2.9 to 13) | 5, 19.2 (8.5 to 37.9), 7.5 (3.3 to 14.7) | 3, 20 (7 to 45.2), 7.8 (2.7 to 17.6) |
| **Cerebellum** | 42, 6.6 (4.9 to 8.8), 0.6 (0.5 to 0.8) | 3, 0.8 (0.3 to 2.3), 0.1 (0 to 0.2) | 4, 3.6 (1.4 to 9), 0.3 (0.1 to 0.8) | 0, 0 (0 to 7.1), 0 (0 to 0.7) | 1, 5.3 (0.9 to 24.6), 0.5 (0.1 to 2.3) | 18, 60 (42.3 to 75.4), 5.6 (3.9 to 7) | 3, 11.5 (4 to 29), 1.1 (0.4 to 2.7) | 13, 86.7 (62.1 to 96.3), 8.1 (5.8 to 9) |
| **PHYLOGENY** | | | | | | | | |
| **NP** | 458, 72.1 (68.5 to 75.5), 1.4 (1.3 to 1.5) | 319, 82.9 (78.8 to 86.3), 1.6 (1.5 to 1.7) | 82, 74.5 (65.7 to 81.8), 1.4 (1.3 to 1.6) | 38, 76 (62.6 to 85.7), 1.5 (1.2 to 1.6) | 11, 57.9 (36.3 to 76.9), 1.1 (0.7 to 1.5) | 1, 3.3 (0.6 to 16.7), 0.1 (0 to 0.3) | 7, 26.9 (13.7 to 46.1), 0.5 (0.3 to 0.9) | 0, 0 (0 to 20.4), 0 (0 to 0.4) |

| | | | | | | | | |
|---|---|---|---|---|---|---|---|---|
| **MsP** | 187, 29.4 (26 to 33.1), 2.6 (2.3 to 3) | 118, 30.6 (26.3 to 35.4), 2.8 (2.4 to 3.2) | 42, 38.2 (29.6 to 47.5), 3.4 (2.7 to 4.3) | 21, 42 (29.4 to 55.8), 3.8 (2.6 to 5) | 5, 26.3 (11.8 to 48.8), 2.4 (1.1 to 4.4) | 1, 3.3 (0.6 to 16.7), 0.3 (0.1 to 1.5) | 0, 0 (0 to 12.9), 0 (0 to 1.2) | 0, 0 (0 to 20.4), 0 (0 to 1.8) |
| **AP/PP** | 82, 12.9 (10.5 to 15.7), 10.3 (8.4 to 12.6) | 43, 11.2 (8.4 to 14.7), 8.9 (6.7 to 11.8) | 26, 23.6 (16.7 to 32.4), 18.9 (13.4 to 25.9) | 11, 22 (12.8 to 35.2), 17.6 (10.2 to 28.2) | 2, 10.5 (2.9 to 31.4), 8.4 (2.4 to 25.2) | 0, 0 (0 to 11.4), 0 (0 to 9.1) | 0, 0 (0 to 12.9), 0 (0 to 10.3) | 0, 0 (0 to 20.4), 0 (0 to 16.3) |
| **SH** | 79, 12.4 (10.1 to 15.2), 1.9 (1.5 to 2.3) | 32, 8.3 (5.9 to 11.5), 1.2 (0.9 to 1.7) | 17, 15.5 (9.9 to 23.4), 2.3 (1.5 to 3.5) | 10, 20 (11.2 to 33), 3 (1.7 to 5) | 2, 10.5 (2.9 to 31.4), 1.6 (0.4 to 4.7) | 9, 30 (16.7 to 47.9), 4.5 (2.5 to 7.2) | 6, 23.1 (11 to 42.1), 3.5 (1.7 to 6.3) | 3, 20 (7 to 45.2), 3 (1.1 to 6.8) |
| **NC** | 26, 4.1 (2.8 to 5.9), 0.5 (0.4 to 0.8) | 3, 0.8 (0.3 to 2.3), 0.1 (0 to 0.3) | 4, 3.6 (1.4 to 9), 0.5 (0.2 to 1.2) | 0, 0 (0 to 7.1), 0 (0 to 0.9) | 0, 0 (0 to 16.8), 0 (0 to 2.2) | 16, 53.3 (36.1 to 69.8), 7.1 (4.8 to 9.2) | 1, 3.8 (0.7 to 18.9), 0.5 (0.1 to 2.5) | 2, 13.3 (3.7 to 37.9), 1.8 (0.5 to 5) |
| **AC/PC** | 27, 4.3 (2.9 to 6.1), 1.3 (0.9 to 1.9) | 3, 0.8 (0.3 to 2.3), 0.2 (0.1 to 0.7) | 4, 3.6 (1.4 to 9), 1.1 (0.4 to 2.8) | 0, 0 (0 to 7.1), 0 (0 to 2.2) | 1, 5.3 (0.9 to 24.6), 1.7 (0.3 to 7.7) | 5, 16.7 (7.3 to 33.6), 5.2 (2.3 to 10.6) | 2, 7.7 (2.1 to 24.1), 2.4 (0.7 to 7.6) | 12, 80 (54.8 to 93), 25.2 (17.2 to 29.2) |
| **ONTOGENY** | | | | | | | | |
| **DP** | 497, 78.3 (74.9 to 81.3), 1.3 (1.3 to 1.4) | 348, 90.4 (87 to 92.9), 1.5 (1.5 to 1.6) | 86, 78.2 (69.6 to 84.9), 1.3 (1.2 to 1.4) | 42, 84 (71.5 to 91.7), 1.4 (1.2 to 1.5) | 12, 63.2 (41 to 80.9), 1.1 (0.7 to 1.4) | 2, 6.7 (1.8 to 21.3), 0.1 (0 to 0.4) | 7, 26.9 (13.7 to 46.1), 0.5 (0.2 to 0.8) | 0, 0 (0 to 20.4), 0 (0 to 0.3) |
| **LPa/VPa** | 119, 18.7 (15.9 to 22), 4.3 (3.6 to 5) | 66, 17.1 (13.7 to 21.2), 3.9 (3.1 to 4.9) | 34, 30.9 (23 to 40.1), 7.1 (5.3 to 9.2) | 15, 30 (19.1 to 43.8), 6.9 (4.4 to 10) | 4, 21.1 (8.5 to 43.3), 4.8 (1.9 to 9.9) | 0, 0 (0 to 11.4), 0 (0 to 2.6) | 0, 0 (0 to 12.9), 0 (0 to 2.9) | 0, 0 (0 to 20.4), 0 (0 to 4.7) |
| **MdP** | 63, 9.9 (7.8 to 12.5), 13.3 (10.5 to 16.8) | 36, 9.4 (6.8 to 12.7), 12.6 (9.2 to 17) | 19, 17.3 (11.3 to 25.4), 23.2 (15.2 to 34.1) | 6, 12 (5.6 to 23.8), 16.1 (7.5 to 32) | 2, 10.5 (2.9 to 31.4), 14.1 (3.9 to 42.2) | 0, 0 (0 to 11.4), 0 (0 to 15.2) | 0, 0 (0 to 12.9), 0 (0 to 17.3) | 0, 0 (0 to 20.4), 0 (0 to 27.4) |
| **SPa** | 71, 11.2 (9 to 13.9), 4.4 (3.5 to 5.4) | 35, 9.1 (6.6 to 12.4), 3.5 (2.6 to 4.8) | 20, 18.2 (12.1 to 26.4), 7.1 (4.7 to 10.3) | 10, 20 (11.2 to 33), 7.8 (4.4 to 12.9) | 2, 10.5 (2.9 to 31.4), 4.1 (1.1 to 12.2) | 4, 13.3 (5.3 to 29.7), 5.2 (2.1 to 11.6) | 0, 0 (0 to 12.9), 0 (0 to 5) | 0, 0 (0 to 20.4), 0 (0 to 8) |
| **H** | 17, 2.7 (1.7 to 4.2), 3.7 (2.3 to 5.8) | 4, 1 (0.4 to 2.6), 1.4 (0.6 to 3.6) | 4, 3.6 (1.4 to 9), 5 (2 to 12.3) | 3, 6 (2.1 to 16.2), 8.2 (2.8 to 22.2) | 1, 5.3 (0.9 to 24.6), 7.2 (1.3 to 33.8) | 5, 16.7 (7.3 to 33.6), 22.9 (10.1 to 46) | 0, 0 (0 to 12.9), 0 (0 to 17.7) | 0, 0 (0 to 20.4), 0 (0 to 28) |
| **T** | 40, 6.3 (4.7 to 8.5), 4.7 (3.5 to 6.3) | 23, 6 (4 to 8.8), 4.5 (3 to 6.6) | 7, 6.4 (3.1 to 12.6), 4.7 (2.3 to 9.4) | 9, 18 (9.8 to 30.8), 13.4 (7.3 to 23) | 0, 0 (0 to 16.8), 0 (0 to 12.6) | 0, 0 (0 to 11.4), 0 (0 to 8.5) | 1, 3.8 (0.7 to 18.9), 2.9 (0.5 to 14.1) | 0, 0 (0 to 20.4), 0 (0 to 15.2) |
| **M** | 18, 2.8 (1.8 to 4.4), 3.1 (2 to 4.8) | 5, 1.3 (0.6 to 3), 1.4 (0.6 to 3.3) | 7, 6.4 (3.1 to 12.6), 6.9 (3.4 to 13.6) | 3, 6 (2.1 to 16.2), 6.5 (2.2 to 17.6) | 0, 0 (0 to 16.8), 0 (0 to 18.3) | 2, 6.7 (1.8 to 21.3), 7.2 (2 to 23.2) | 0, 0 (0 to 12.9), 0 (0 to 14) | 1, 6.7 (1.2 to 29.8), 7.2 (1.3 to 32.4) |
| **R** | 59, 9.3 (7.3 to 11.8), 0.8 (0.6 to 1) | 6, 1.6 (0.7 to 3.4), 0.1 (0.1 to 0.3) | 7, 6.4 (3.1 to 12.6), 0.5 (0.3 to 1) | 1, 2 (0.4 to 10.5), 0.2 (0 to 0.9) | 1, 5.3 (0.9 to 24.6), 0.4 (0.1 to 2) | 21, 70 (52.1 to 83.3), 5.7 (4.2 to 6.8) | 8, 30.8 (16.5 to 50), 2.5 (1.3 to 4.1) | 15, 100 (79.6 to 100), 8.1 (6.5 to 8.1) |

**Supplemental Table 8. General anatomical patterns of metastases subentities**

The involvement of general topographic, phylogenetic and ontogenetic parcellation units by metastases in total and by organ of origin is given in absolute numbers (n), relative tumor prevalence (RTP) and relative tumor density (RTD). *AC/PC = Archi- and Paleocerebellum; AP/PP = Archi- and Paleopallium; DPa = dorsal pallium; GIT = gastrointestinal tract; H = hypothalamus; LPa/VPa = lateral/ventral pallium; M = mesencephalon; MsP = Mesopallium; MdP = medial pallium; NC = Neocerebellum; NP = Neopallium; R = rhombencephalon; SH = Stammhirn; SPa = Subpallium; T = thalamus; UGT = urogenital tract.*

| | Metastases total (n, % [95% CI], Vnorm [95% CI]) | Lung (n, % [95% CI], Vnorm [95% CI]) | Skin (n, % [95% CI], Vnorm [95% CI]) | GIT (n, % [95% CI], Vnorm [95% CI]) | Breast (n, % [95% CI], Vnorm [95% CI]) | UGT (n, % [95% CI], Vnorm [95% CI]) | Miscellaneous (n, % [95% CI], Vnorm [95% CI]) |
|---|---|---|---|---|---|---|---|
| **Total** | 299, 100 (98.7 to 100), 1 (1 to 1) | 143, 100 (97.4 to 100), 1 (1 to 1) | 49, 100 (92.7 to 100), 1 (0.9 to 1) | 37, 100 (90.6 to 100), 1 (0.9 to 1) | 28, 100 (87.9 to 100), 1 (0.9 to 1) | 24, 100 (86.2 to 100), 1 (0.9 to 1) | 18, 100 (82.4 to 100), 1 (0.8 to 1) |
| **TOPOGRAPHY** | | | | | | | |
| **Frontal lobe** | 108, 36.1 (30.9 to 41.7), 2 (1.7 to 2.3) | 54, 37.8 (30.2 to 45.9), 2.1 (1.7 to 2.5) | 20, 40.8 (28.2 to 54.8), 2.2 (1.5 to 3) | 9, 24.3 (13.4 to 40.1), 1.3 (0.7 to 2.2) | 7, 25 (12.7 to 43.4), 1.4 (0.7 to 2.4) | 12, 50 (31.4 to 68.6), 2.7 (1.7 to 3.8) | 6, 33.3 (16.3 to 56.3), 1.8 (0.9 to 3.1) |
| **Central lobe** | 63, 21.1 (16.8 to 26), 2.8 (2.3 to 3.5) | 30, 21 (15.1 to 28.4), 2.8 (2 to 3.8) | 11, 22.4 (13 to 35.9), 3 (1.7 to 4.8) | 8, 21.6 (11.4 to 37.2), 2.9 (1.5 to 5) | 5, 17.9 (7.9 to 35.6), 2.4 (1.1 to 4.8) | 6, 25 (12 to 44.9), 3.3 (1.6 to 6) | 3, 16.7 (5.8 to 39.2), 2.2 (0.8 to 5.3) |
| **Parietal lobe** | 63, 21.1 (16.8 to 26), 1.8 (1.4 to 2.2) | 24, 16.8 (11.5 to 23.8), 1.4 (1 to 2) | 15, 30.6 (19.5 to 44.5), 2.6 (1.7 to 3.8) | 6, 16.2 (7.7 to 31.1), 1.4 (0.6 to 2.6) | 7, 25 (12.7 to 43.4), 2.1 (1.1 to 3.7) | 5, 20.8 (9.2 to 40.5), 1.8 (0.8 to 3.4) | 6, 33.3 (16.3 to 56.3), 2.8 (1.4 to 4.8) |
| **Occipital lobe** | 68, 22.7 (18.4 to 27.8), 3.3 (2.6 to 4) | 32, 22.4 (16.3 to 29.9), 3.2 (2.3 to 4.3) | 15, 30.6 (19.5 to 44.5), 4.4 (2.8 to 6.4) | 8, 21.6 (11.4 to 37.2), 3.1 (1.6 to 5.4) | 5, 17.9 (7.9 to 35.6), 2.6 (1.1 to 5.1) | 6, 25 (12 to 44.9), 3.6 (1.7 to 6.5) | 2, 11.1 (3.1 to 32.8), 1.6 (0.4 to 4.7) |
| **Temporal lobe** | 49, 16.4 (12.6 to 21), 1.5 (1.2 to 2) | 21, 14.7 (9.8 to 21.4), 1.4 (0.9 to 2) | 16, 32.7 (21.2 to 46.6), 3.1 (2 to 4.4) | 3, 8.1 (2.8 to 21.3), 0.8 (0.3 to 2) | 4, 14.3 (5.7 to 31.5), 1.3 (0.5 to 3) | 2, 8.3 (2.3 to 25.8), 0.8 (0.2 to 2.4) | 3, 16.7 (5.8 to 39.2), 1.6 (0.5 to 3.7) |
| **Insular lobe** | 7, 2.3 (1.1 to 4.8), 1 (0.5 to 2.1) | 1, 0.7 (0.1 to 3.9), 0.3 (0.1 to 1.7) | 4, 8.2 (3.2 to 19.2), 3.6 (1.4 to 8.5) | 1, 2.7 (0.5 to 13.8), 1.2 (0.2 to 6.2) | 0, 0 (0 to 12.1), 0 (0 to 5.4) | 0, 0 (0 to 13.8), 0 (0 to 6.1) | 1, 5.6 (1 to 25.8), 2.5 (0.4 to 11.5) |
| **Limbic lobe** | 21, 7 (4.6 to 10.5), 1 (0.7 to 1.5) | 9, 6.3 (3.3 to 11.5), 0.9 (0.5 to 1.7) | 7, 14.3 (7.1 to 26.7), 2.1 (1 to 3.8) | 3, 8.1 (2.8 to 21.3), 1.2 (0.4 to 3.1) | 0, 0 (0 to 12.1), 0 (0 to 1.7) | 1, 4.2 (0.7 to 20.2), 0.6 (0.1 to 2.9) | 1, 5.6 (1 to 25.8), 0.8 (0.1 to 3.7) |
| **Basal ganglia** | 8, 2.7 (1.4 to 5.2), 1.3 (0.7 to 2.6) | 2, 1.4 (0.4 to 5), 0.7 (0.2 to 2.4) | 3, 6.1 (2.1 to 16.5), 3 (1 to 8.1) | 0, 0 (0 to 9.4), 0 (0 to 4.6) | 2, 7.1 (2 to 22.6), 3.5 (1 to 11.2) | 0, 0 (0 to 13.8), 0 (0 to 6.8) | 1, 5.6 (1 to 25.8), 2.7 (0.5 to 12.7) |
| **Diencephalon** | 7, 2.3 (1.1 to 4.8), 1.1 (0.6 to 2.3) | 2, 1.4 (0.4 to 5), 0.7 (0.2 to 2.4) | 2, 4.1 (1.1 to 13.7), 2 (0.5 to 6.6) | 1, 2.7 (0.5 to 13.8), 1.3 (0.2 to 6.7) | 0, 0 (0 to 12.1), 0 (0 to 5.8) | 0, 0 (0 to 13.8), 0 (0 to 6.7) | 2, 11.1 (3.1 to 32.8), 5.4 (1.5 to 15.9) |
| **Brainstem** | 8, 2.7 (1.4 to 5.2), 1 (0.5 to 2) | 2, 1.4 (0.4 to 5), 0.5 (0.1 to 1.9) | 4, 8.2 (3.2 to 19.2), 3.2 (1.3 to 7.5) | 0, 0 (0 to 9.4), 0 (0 to 3.7) | 0, 0 (0 to 12.1), 0 (0 to 4.7) | 0, 0 (0 to 13.8), 0 (0 to 5.4) | 2, 11.1 (3.1 to 32.8), 4.3 (1.2 to 12.7) |
| **Cerebellum** | 88, 29.4 (24.6 to 34.8), 2.7 (2.3 to 3.2) | 50, 35 (27.6 to 43.1), 3.3 (2.6 to 4) | 8, 16.3 (8.5 to 29), 1.5 (0.8 to 2.7) | 12, 32.4 (19.6 to 48.5), 3 (1.8 to 4.5) | 10, 35.7 (20.7 to 54.2), 3.3 (1.9 to 5) | 2, 8.3 (2.3 to 25.8), 0.8 (0.2 to 2.4) | 6, 33.3 (16.3 to 56.3), 3.1 (1.5 to 5.2) |
| **PHYLOGENY** | | | | | | | |
| **NP** | 225, 75.3 (70.1 to 79.8), 1.4 (1.3 to 1.5) | 110, 76.9 (69.4 to 83.1), 1.5 (1.3 to 1.6) | 35, 71.4 (57.6 to 82.2), 1.4 (1.1 to 1.6) | 28, 75.7 (59.9 to 86.6), 1.5 (1.2 to 1.7) | 18, 64.3 (45.8 to 79.3), 1.2 (0.9 to 1.5) | 23, 95.8 (79.8 to 99.3), 1.8 (1.5 to 1.9) | 11, 61.1 (38.6 to 79.7), 1.2 (0.7 to 1.5) |
| **MsP** | 21, 7 (4.6 to 10.5), 0.6 (0.4 to 0.9) | 10, 7 (3.8 to 12.4), 0.6 (0.3 to 1.1) | 6, 12.2 (5.7 to 24.2), 1.1 (0.5 to 2.2) | 4, 10.8 (4.3 to 24.7), 1 (0.4 to 2.2) | 0, 0 (0 to 12.1), 0 (0 to 1.1) | 1, 4.2 (0.7 to 20.2), 0.4 (0.1 to 1.8) | 0, 0 (0 to 17.6), 0 (0 to 1.6) |
| **AP/PP** | 4, 1.3 (0.5 to 3.4), 1.1 (0.4 to 2.7) | 2, 1.4 (0.4 to 5), 1.1 (0.3 to 4) | 2, 4.1 (1.1 to 13.7), 3.3 (0.9 to 11) | 0, 0 (0 to 9.4), 0 (0 to 7.5) | 0, 0 (0 to 12.1), 0 (0 to 9.7) | 0, 0 (0 to 13.8), 0 (0 to 11.1) | 0, 0 (0 to 17.6), 0 (0 to 14.1) |
| **SH** | 21, 7 (4.6 to 10.5), 1.1 (0.7 to 1.6) | 6, 4.2 (1.9 to 8.9), 0.6 (0.3 to 1.3) | 8, 16.3 (8.5 to 29), 2.4 (1.3 to 4.4) | 1, 2.7 (0.5 to 13.8), 0.4 (0.1 to 2.1) | 2, 7.1 (2 to 22.6), 1.1 (0.3 to 3.4) | 0, 0 (0 to 13.8), 0 (0 to 2.1) | 4, 22.2 (9 to 45.2), 3.3 (1.3 to 6.8) |

| | | | | | | | |
|---|---|---|---|---|---|---|---|
| **NC** | 83, 27.8 (23 to 33.1), 3.7 (3 to 4.4) | 48, 33.6 (26.3 to 41.6), 4.4 (3.5 to 5.5) | 7, 14.3 (7.1 to 26.7), 1.9 (0.9 to 3.5) | 12, 32.4 (19.6 to 48.5), 4.3 (2.6 to 6.4) | 11, 39.3 (23.6 to 57.6), 5.2 (3.1 to 7.6) | 1, 4.2 (0.7 to 20.2), 0.6 (0.1 to 2.7) | 4, 22.2 (9 to 45.2), 2.9 (1.2 to 6) |
| **AC/PC** | 12, 4 (2.3 to 6.9), 1.3 (0.7 to 2.2) | 5, 3.5 (1.5 to 7.9), 1.1 (0.5 to 2.5) | 1, 2 (0.4 to 10.7), 0.6 (0.1 to 3.4) | 1, 2.7 (0.5 to 13.8), 0.8 (0.2 to 4.3) | 0, 0 (0 to 12.1), 0 (0 to 3.8) | 1, 4.2 (0.7 to 20.2), 1.3 (0.2 to 6.4) | 4, 22.2 (9 to 45.2), 7 (2.8 to 14.2) |
| **ONTOGENY** | | | | | | | |
| **DP** | 232, 77.6 (72.5 to 82), 1.3 (1.2 to 1.4) | 113, 79 (71.6 to 84.9), 1.3 (1.2 to 1.4) | 37, 75.5 (61.9 to 85.4), 1.3 (1 to 1.4) | 30, 81.1 (65.8 to 90.5), 1.4 (1.1 to 1.5) | 18, 64.3 (45.8 to 79.3), 1.1 (0.8 to 1.3) | 23, 95.8 (79.8 to 99.3), 1.6 (1.3 to 1.7) | 11, 61.1 (38.6 to 79.7), 1 (0.7 to 1.3) |
| **LPa/VPa** | 10, 3.3 (1.8 to 6), 0.8 (0.4 to 1.4) | 3, 2.1 (0.7 to 6), 0.5 (0.2 to 1.4) | 4, 8.2 (3.2 to 19.2), 1.9 (0.7 to 4.4) | 2, 5.4 (1.5 to 17.7), 1.2 (0.3 to 4.1) | 0, 0 (0 to 12.1), 0 (0 to 2.8) | 1, 4.2 (0.7 to 20.2), 1 (0.2 to 4.6) | 0, 0 (0 to 17.6), 0 (0 to 4) |
| **MdP** | 3, 1 (0.3 to 2.9), 1.3 (0.5 to 3.9) | 1, 0.7 (0.1 to 3.9), 0.9 (0.2 to 5.2) | 2, 4.1 (1.1 to 13.7), 5.5 (1.5 to 18.4) | 0, 0 (0 to 9.4), 0 (0 to 12.6) | 0, 0 (0 to 12.1), 0 (0 to 16.2) | 0, 0 (0 to 13.8), 0 (0 to 18.5) | 0, 0 (0 to 17.6), 0 (0 to 23.6) |
| **SPa** | 10, 3.3 (1.8 to 6), 1.3 (0.7 to 2.4) | 3, 2.1 (0.7 to 6), 0.8 (0.3 to 2.3) | 4, 8.2 (3.2 to 19.2), 3.2 (1.3 to 7.5) | 0, 0 (0 to 9.4), 0 (0 to 3.7) | 2, 7.1 (2 to 22.6), 2.8 (0.8 to 8.8) | 0, 0 (0 to 13.8), 0 (0 to 5.4) | 1, 5.6 (1 to 25.8), 2.2 (0.4 to 10) |
| **H** | 0, 0 (0 to 1.3), 0 (0 to 1.7) | 0, 0 (0 to 2.6), 0 (0 to 3.6) | 0, 0 (0 to 7.3), 0 (0 to 10) | 0, 0 (0 to 9.4), 0 (0 to 12.9) | 0, 0 (0 to 12.1), 0 (0 to 16.6) | 0, 0 (0 to 13.8), 0 (0 to 18.9) | 0, 0 (0 to 17.6), 0 (0 to 24.1) |
| **T** | 7, 2.3 (1.1 to 4.8), 1.7 (0.8 to 3.5) | 2, 1.4 (0.4 to 5), 1 (0.3 to 3.7) | 2, 4.1 (1.1 to 13.7), 3 (0.8 to 10.2) | 1, 2.7 (0.5 to 13.8), 2 (0.4 to 10.3) | 0, 0 (0 to 12.1), 0 (0 to 9) | 0, 0 (0 to 13.8), 0 (0 to 10.3) | 2, 11.1 (3.1 to 32.8), 8.3 (2.3 to 24.5) |
| **M** | 1, 0.3 (0.1 to 1.9), 0.4 (0.1 to 2) | 0, 0 (0 to 2.6), 0 (0 to 2.8) | 0, 0 (0 to 7.3), 0 (0 to 7.9) | 0, 0 (0 to 9.4), 0 (0 to 10.2) | 0, 0 (0 to 12.1), 0 (0 to 13.1) | 0, 0 (0 to 13.8), 0 (0 to 15) | 1, 5.6 (1 to 25.8), 6 (1.1 to 28) |
| **R** | 94, 31.4 (26.4 to 36.9), 2.6 (2.1 to 3) | 52, 36.4 (28.9 to 44.5), 3 (2.4 to 3.6) | 9, 18.4 (10 to 31.4), 1.5 (0.8 to 2.5) | 12, 32.4 (19.6 to 48.5), 2.6 (1.6 to 3.9) | 11, 39.3 (23.6 to 57.6), 3.2 (1.9 to 4.7) | 2, 8.3 (2.3 to 25.8), 0.7 (0.2 to 2.1) | 8, 44.4 (24.6 to 66.3), 3.6 (2 to 5.4) |

**Supplemental Table 9. Gyral patterns of neuroepithelial tumors and brain metastases**

The involvement of the gyral segments by brain tumors in total, neuroepithelial tumors (NT) or metastases is given in absolute numbers (n), relative tumor prevalence (RTP) and relative tumor density (RTD). *ANG = angular gyrus; F1 = superior frontal gyrus; F2 = middle frontal gyrus; F3 = inferior frontal gyrus; O1 = superior occipital gyrus; O2 = middle occipital gyrus; O3 = inferior occipital gyrus; PHG = parahippocampal gyrus; SCA = subcallosal area; SMG = supramarginal gyrus; SPL = superior parietal lobule; T1 = superior temporal gyrus; T2 = middle temporal gyrus; T3 = inferior temporal gyrus.*

| | Total (n, RTP [95% CI], RTD [95% CI]) | NT (n, RTP [95% CI], RTD [95% CI]) | Metastases (n, RTP [95% CI], RTD [95% CI]) |
|---|---|---|---|
| **Total** | 1000, 100 (99.6 to 100), 1 (1 to 1) | 657, 100 (99.4 to 100), 1 (1 to 1) | 299, 100 (98.7 to 100), 1 (1 to 1) |
| **Frontal pole** | 17, 1.7 (1.1 to 2.7), 3.9 (2.5 to 6.3) | 11, 1.7 (0.9 to 3), 3.9 (2.2 to 6.9) | 4, 1.3 (0.5 to 3.4), 3.1 (1.2 to 7.9) |
| **F1** | 131, 13.1 (11.1 to 15.3), 2.1 (1.8 to 2.4) | 81, 12.3 (10 to 15.1), 2 (1.6 to 2.4) | 42, 14 (10.6 to 18.4), 2.2 (1.7 to 2.9) |
| **F2** | 108, 10.8 (9 to 12.9), 2 (1.7 to 2.4) | 57, 8.7 (6.8 to 11.1), 1.6 (1.3 to 2.1) | 46, 15.4 (11.7 to 19.9), 2.9 (2.2 to 3.7) |
| **F3 orbital** | 29, 2.9 (2 to 4.1), 8.1 (5.7 to 11.5) | 23, 3.5 (2.3 to 5.2), 9.8 (6.5 to 14.5) | 2, 0.7 (0.2 to 2.4), 1.9 (0.5 to 6.7) |
| **F3 triangular** | 36, 3.6 (2.6 to 4.9), 3.4 (2.4 to 4.6) | 25, 3.8 (2.6 to 5.6), 3.6 (2.4 to 5.2) | 7, 2.3 (1.1 to 4.8), 2.2 (1.1 to 4.4) |
| **F3 opercular** | 41, 4.1 (3 to 5.5), 3.2 (2.4 to 4.3) | 31, 4.7 (3.3 to 6.6), 3.7 (2.6 to 5.2) | 5, 1.7 (0.7 to 3.9), 1.3 (0.6 to 3) |
| **Anterior orbital** | 11, 1.1 (0.6 to 2), 3.6 (2 to 6.4) | 9, 1.4 (0.7 to 2.6), 4.5 (2.4 to 8.4) | 1, 0.3 (0.1 to 1.9), 1.1 (0.2 to 6.1) |
| **Medial orbital** | 18, 1.8 (1.1 to 2.8), 2.6 (1.7 to 4.1) | 14, 2.1 (1.3 to 3.5), 3.1 (1.8 to 5.1) | 2, 0.7 (0.2 to 2.4), 1 (0.3 to 3.5) |
| **Lateral orbital** | 15, 1.5 (0.9 to 2.5), 2.7 (1.6 to 4.4) | 11, 1.7 (0.9 to 3), 3 (1.7 to 5.4) | 3, 1 (0.3 to 2.9), 1.8 (0.6 to 5.3) |
| **Posterior orbital** | 36, 3.6 (2.6 to 4.9), 5.3 (3.9 to 7.3) | 33, 5 (3.6 to 7), 7.4 (5.3 to 10.3) | 1, 0.3 (0.1 to 1.9), 0.5 (0.1 to 2.8) |
| **Rectus** | 16, 1.6 (1 to 2.6), 1.8 (1.1 to 2.8) | 15, 2.3 (1.4 to 3.7), 2.5 (1.5 to 4.1) | 0, 0 (0 to 1.3), 0 (0 to 1.4) |
| **Rostral** | 13, 1.3 (0.8 to 2.2), 3.9 (2.3 to 6.7) | 12, 1.8 (1 to 3.2), 5.5 (3.2 to 9.6) | 1, 0.3 (0.1 to 1.9), 1 (0.2 to 5.7) |
| **Precentral** | 61, 6.1 (4.8 to 7.8), 1.7 (1.3 to 2.2) | 26, 4 (2.7 to 5.7), 1.1 (0.8 to 1.6) | 32, 10.7 (7.7 to 14.7), 3 (2.1 to 4.1) |
| **Postcentral** | 48, 4.8 (3.6 to 6.3), 2.4 (1.8 to 3.1) | 26, 4 (2.7 to 5.7), 1.9 (1.3 to 2.8) | 18, 6 (3.8 to 9.3), 3 (1.9 to 4.6) |
| **Paracentral lobule** | 32, 3.2 (2.3 to 4.5), 2.5 (1.8 to 3.5) | 19, 2.9 (1.9 to 4.5), 2.2 (1.4 to 3.5) | 10, 3.3 (1.8 to 6), 2.6 (1.4 to 4.7) |
| **Subcentral** | 38, 3.8 (2.8 to 5.2), 6.9 (5 to 9.3) | 30, 4.6 (3.2 to 6.4), 8.3 (5.8 to 11.6) | 6, 2 (0.9 to 4.3), 3.6 (1.7 to 7.8) |
| **SPL** | 43, 4.3 (3.2 to 5.7), 1.4 (1.1 to 1.9) | 24, 3.7 (2.5 to 5.4), 1.2 (0.8 to 1.8) | 17, 5.7 (3.6 to 8.9), 1.9 (1.2 to 2.9) |
| **SMG** | 70, 7 (5.6 to 8.8), 2.5 (2 to 3.1) | 55, 8.4 (6.5 to 10.7), 3 (2.3 to 3.8) | 13, 4.3 (2.6 to 7.3), 1.5 (0.9 to 2.6) |
| **ANG** | 49, 4.9 (3.7 to 6.4), 1.6 (1.2 to 2) | 37, 5.6 (4.1 to 7.7), 1.8 (1.3 to 2.4) | 11, 3.7 (2.1 to 6.5), 1.2 (0.7 to 2.1) |
| **Precuneus** | 61, 6.1 (4.8 to 7.8), 2.2 (1.7 to 2.8) | 38, 5.8 (4.2 to 7.8), 2.1 (1.5 to 2.8) | 20, 6.7 (4.4 to 10.1), 2.4 (1.6 to 3.6) |
| **Cuneus** | 39, 3.9 (2.9 to 5.3), 4.2 (3.1 to 5.7) | 23, 3.5 (2.3 to 5.2), 3.8 (2.5 to 5.6) | 14, 4.7 (2.8 to 7.7), 5.1 (3 to 8.4) |
| **O1** | 23, 2.3 (1.5 to 3.4), 2.8 (1.8 to 4.1) | 16, 2.4 (1.5 to 3.9), 2.9 (1.8 to 4.7) | 6, 2 (0.9 to 4.3), 2.4 (1.1 to 5.2) |
| **O2** | 52, 5.2 (4 to 6.8), 3.6 (2.8 to 4.7) | 25, 3.8 (2.6 to 5.6), 2.6 (1.8 to 3.9) | 27, 9 (6.3 to 12.8), 6.3 (4.4 to 8.9) |
| **O3** | 19, 1.9 (1.2 to 2.9), 2.2 (1.4 to 3.5) | 9, 1.4 (0.7 to 2.6), 1.6 (0.9 to 3.1) | 10, 3.3 (1.8 to 6), 4 (2.2 to 7.1) |
| **Occipital pole** | 3, 0.3 (0.1 to 0.9), 0.3 (0.1 to 0.8) | 2, 0.3 (0.1 to 1.1), 0.3 (0.1 to 1.1) | 1, 0.3 (0.1 to 1.9), 0.3 (0.1 to 1.8) |
| **Lingual** | 31, 3.1 (2.2 to 4.4), 1.7 (1.2 to 2.3) | 20, 3 (2 to 4.7), 1.6 (1.1 to 2.5) | 10, 3.3 (1.8 to 6), 1.8 (1 to 3.2) |
| **Fusiform** | 53, 5.3 (4.1 to 6.9), 2.6 (2 to 3.4) | 43, 6.5 (4.9 to 8.7), 3.2 (2.4 to 4.3) | 10, 3.3 (1.8 to 6), 1.6 (0.9 to 3) |
| **Temporal pole** | 47, 4.7 (3.6 to 6.2), 2 (1.5 to 2.7) | 45, 6.8 (5.2 to 9), 3 (2.2 to 3.9) | 2, 0.7 (0.2 to 2.4), 0.3 (0.1 to 1) |
| **T1** | 70, 7 (5.6 to 8.8), 2.8 (2.2 to 3.5) | 62, 9.4 (7.4 to 11.9), 3.8 (3 to 4.8) | 8, 2.7 (1.4 to 5.2), 1.1 (0.5 to 2.1) |
| **T2** | 73, 7.3 (5.8 to 9.1), 3.4 (2.7 to 4.2) | 62, 9.4 (7.4 to 11.9), 4.3 (3.4 to 5.5) | 11, 3.7 (2.1 to 6.5), 1.7 (1 to 3) |

| | | | |
|---|---|---|---|
| **T3** | 56, 5.6 (4.3 to 7.2), 12.2 (9.5 to 15.7) | 45, 6.8 (5.2 to 9), 15 (11.3 to 19.8) | 11, 3.7 (2.1 to 6.5), 8 (4.5 to 14.1) |
| **Planum temporale** | 8, 0.8 (0.4 to 1.6), 2.9 (1.4 to 5.6) | 7, 1.1 (0.5 to 2.2), 3.8 (1.8 to 7.8) | 1, 0.3 (0.1 to 1.9), 1.2 (0.2 to 6.7) |
| **Planum polare** | 23, 2.3 (1.5 to 3.4), 2.5 (1.7 to 3.8) | 21, 3.2 (2.1 to 4.8), 3.5 (2.3 to 5.3) | 2, 0.7 (0.2 to 2.4), 0.7 (0.2 to 2.6) |
| **Short insular** | 51, 5.1 (3.9 to 6.6), 3.6 (2.8 to 4.7) | 49, 7.5 (5.7 to 9.7), 5.3 (4 to 6.9) | 1, 0.3 (0.1 to 1.9), 0.2 (0 to 1.3) |
| **Long insular** | 47, 4.7 (3.6 to 6.2), 5.6 (4.3 to 7.4) | 45, 6.8 (5.2 to 9), 8.2 (6.2 to 10.9) | 1, 0.3 (0.1 to 1.9), 0.4 (0.1 to 2.2) |
| **SCA** | 21, 2.1 (1.4 to 3.2), 9.7 (6.3 to 14.7) | 19, 2.9 (1.9 to 4.5), 13.3 (8.6 to 20.6) | 0, 0 (0 to 1.3), 0 (0 to 5.8) |
| **Cingulate anterior** | 44, 4.4 (3.3 to 5.9), 3.3 (2.5 to 4.4) | 38, 5.8 (4.2 to 7.8), 4.4 (3.2 to 5.9) | 4, 1.3 (0.5 to 3.4), 1 (0.4 to 2.6) |
| **Cingulate middle** | 35, 3.5 (2.5 to 4.8), 2.2 (1.6 to 3.1) | 28, 4.3 (3 to 6.1), 2.7 (1.9 to 3.9) | 6, 2 (0.9 to 4.3), 1.3 (0.6 to 2.8) |
| **Cingulate posterior** | 36, 3.6 (2.6 to 4.9), 2 (1.4 to 2.7) | 32, 4.9 (3.5 to 6.8), 2.6 (1.9 to 3.7) | 3, 1 (0.3 to 2.9), 0.5 (0.2 to 1.6) |
| **PHG** | 52, 5.2 (4 to 6.8), 5.4 (4.2 to 7.1) | 49, 7.5 (5.7 to 9.7), 7.8 (5.9 to 10.2) | 2, 0.7 (0.2 to 2.4), 0.7 (0.2 to 2.5) |
| **Hippocampus** | 69, 6.9 (5.5 to 8.6), 9.3 (7.4 to 11.6) | 63, 9.6 (7.6 to 12.1), 12.9 (10.2 to 16.2) | 3, 1 (0.3 to 2.9), 1.3 (0.5 to 3.9) |
| **Amygdala** | 55, 5.5 (4.2 to 7.1), 19.2 (14.8 to 24.8) | 50, 7.6 (5.8 to 9.9), 26.6 (20.3 to 34.5) | 2, 0.7 (0.2 to 2.4), 2.3 (0.6 to 8.4) |

**Supplemental Table 10. Gyral patterns of neuroepithelial tumor subentities**

The involvement of the gyral segments by neuroepithelial tumors (NT) in total, glioblastoma (GBM), WHO grade III (gIIIG) and II (gIIG) glioma, developmental tumors (DT), pilocytic astrocytoma (PA), ependymoma (EP), or medulloblastoma (MB) is given in absolute numbers (n), relative tumor prevalence (RTP) and relative tumor density (RTD). *ANG = angular gyrus; F1 = superior frontal gyrus; F2 = middle frontal gyrus; F3 = inferior frontal gyrus; O1 = superior occipital gyrus; O2 = middle occipital gyrus; O3 = inferior occipital gyrus; PHG = parahippocampal gyrus; SCA = subcallosal area; SMG = supramarginal gyrus; SPL = superior parietal lobule; T1 = superior temporal gyrus; T2 = middle temporal gyrus; T3 = inferior temporal gyrus.*

| | **Total (n, RTP [95% CI], RTD [95% CI])** | **GBM (n, RTP [95% CI], RTD [95% CI])** | **gIIIG (n, RTP [95% CI], RTD [95% CI])** | **gIIG (n, RTP [95% CI], RTD [95% CI])** | **DT (n, RTP [95% CI], RTD [95% CI])** | **PA (n, RTP [95% CI], RTD [95% CI])** | **EP (n, RTP [95% CI], RTD [95% CI])** | **MB (n, RTP [95% CI], RTD [95% CI])** |
|---|---|---|---|---|---|---|---|---|
| **Total** | 635, 100 (99.4 to 100), 1 (1 to 1) | 385, 100 (99 to 100), 1 (1 to 1) | 110, 100 (96.6 to 100), 1 (1 to 1) | 50, 100 (92.9 to 100), 1 (0.9 to 1) | 19, 100 (83.2 to 100), 1 (0.8 to 1) | 30, 100 (88.6 to 100), 1 (0.9 to 1) | 26, 100 (87.1 to 100), 1 (0.9 to 1) | 15, 100 (79.6 to 100), 1 (0.8 to 1) |
| **Frontal pole** | 11, 1.7 (1 to 3.1), 4 (2.3 to 7.1) | 4, 1 (0.4 to 2.6), 2.4 (0.9 to 6.1) | 6, 5.5 (2.5 to 11.4), 12.7 (5.9 to 26.4) | 1, 2 (0.4 to 10.5), 4.6 (0.8 to 24.4) | 0, 0 (0 to 16.8), 0 (0 to 39) | 0, 0 (0 to 11.4), 0 (0 to 26.3) | 0, 0 (0 to 12.9), 0 (0 to 29.9) | 0, 0 (0 to 20.4), 0 (0 to 47.3) |
| **F1** | 80, 12.6 (10.2 to 15.4), 2 (1.6 to 2.5) | 44, 11.4 (8.6 to 15), 1.8 (1.4 to 2.4) | 25, 22.7 (15.9 to 31.4), 3.6 (2.5 to 5) | 11, 22 (12.8 to 35.2), 3.5 (2 to 5.6) | 0, 0 (0 to 16.8), 0 (0 to 2.7) | 0, 0 (0 to 11.4), 0 (0 to 1.8) | 0, 0 (0 to 12.9), 0 (0 to 2) | 0, 0 (0 to 20.4), 0 (0 to 3.2) |
| **F2** | 56, 8.8 (6.9 to 11.3), 1.7 (1.3 to 2.1) | 36, 9.4 (6.8 to 12.7), 1.8 (1.3 to 2.4) | 12, 10.9 (6.4 to 18.1), 2 (1.2 to 3.4) | 8, 16 (8.3 to 28.5), 3 (1.6 to 5.3) | 0, 0 (0 to 16.8), 0 (0 to 3.2) | 0, 0 (0 to 11.4), 0 (0 to 2.1) | 0, 0 (0 to 12.9), 0 (0 to 2.4) | 0, 0 (0 to 20.4), 0 (0 to 3.8) |
| **F3 orbital** | 22, 3.5 (2.3 to 5.2), 9.7 (6.4 to 14.5) | 16, 4.2 (2.6 to 6.6), 11.6 (7.2 to 18.6) | 4, 3.6 (1.4 to 9), 10.2 (4 to 25.1) | 2, 4 (1.1 to 13.5), 11.2 (3.1 to 37.6) | 0, 0 (0 to 16.8), 0 (0 to 47) | 0, 0 (0 to 11.4), 0 (0 to 31.7) | 0, 0 (0 to 12.9), 0 (0 to 36) | 0, 0 (0 to 20.4), 0 (0 to 56.9) |
| **F3 triangular** | 24, 3.8 (2.6 to 5.6), 3.5 (2.4 to 5.2) | 15, 3.9 (2.4 to 6.3), 3.6 (2.2 to 5.9) | 6, 5.5 (2.5 to 11.4), 5.1 (2.4 to 10.6) | 3, 6 (2.1 to 16.2), 5.6 (1.9 to 15.1) | 0, 0 (0 to 16.8), 0 (0 to 15.7) | 0, 0 (0 to 11.4), 0 (0 to 10.6) | 0, 0 (0 to 12.9), 0 (0 to 12) | 0, 0 (0 to 20.4), 0 (0 to 19) |
| **F3 opercular** | 30, 4.7 (3.3 to 6.7), 3.7 (2.6 to 5.2) | 22, 5.7 (3.8 to 8.5), 4.5 (3 to 6.7) | 5, 4.5 (2 to 10.2), 3.6 (1.5 to 8) | 3, 6 (2.1 to 16.2), 4.7 (1.6 to 12.7) | 0, 0 (0 to 16.8), 0 (0 to 13.2) | 0, 0 (0 to 11.4), 0 (0 to 8.9) | 0, 0 (0 to 12.9), 0 (0 to 10.1) | 0, 0 (0 to 20.4), 0 (0 to 16) |
| **Anterior orbital** | 9, 1.4 (0.7 to 2.7), 4.6 (2.4 to 8.7) | 4, 1 (0.4 to 2.6), 3.4 (1.3 to 8.6) | 4, 3.6 (1.4 to 9), 11.9 (4.7 to 29.4) | 1, 2 (0.4 to 10.5), 6.5 (1.2 to 34.3) | 0, 0 (0 to 16.8), 0 (0 to 55) | 0, 0 (0 to 11.4), 0 (0 to 37.1) | 0, 0 (0 to 12.9), 0 (0 to 42.1) | 0, 0 (0 to 20.4), 0 (0 to 66.7) |
| **Medial orbital** | 14, 2.2 (1.3 to 3.7), 3.2 (1.9 to 5.3) | 4, 1 (0.4 to 2.6), 1.5 (0.6 to 3.8) | 5, 4.5 (2 to 10.2), 6.6 (2.8 to 14.8) | 4, 8 (3.2 to 18.8), 11.6 (4.6 to 27.3) | 1, 5.3 (0.9 to 24.6), 7.6 (1.4 to 35.7) | 0, 0 (0 to 11.4), 0 (0 to 16.4) | 0, 0 (0 to 12.9), 0 (0 to 18.6) | 0, 0 (0 to 20.4), 0 (0 to 29.5) |

| | | | | | | | | |
|---|---|---|---|---|---|---|---|---|
| **Lateral orbital** | 11, 1.7 (1 to 3.1), 3.1 (1.8 to 5.6) | 6, 1.6 (0.7 to 3.4), 2.8 (1.3 to 6.1) | 4, 3.6 (1.4 to 9), 6.6 (2.6 to 16.2) | 1, 2 (0.4 to 10.5), 3.6 (0.6 to 19) | 0, 0 (0 to 16.8), 0 (0 to 30.4) | 0, 0 (0 to 11.4), 0 (0 to 20.5) | 0, 0 (0 to 12.9), 0 (0 to 23.3) | 0, 0 (0 to 20.4), 0 (0 to 36.8) |
| **Posterior orbital** | 33, 5.2 (3.7 to 7.2), 7.7 (5.5 to 10.6) | 15, 3.9 (2.4 to 6.3), 5.8 (3.5 to 9.3) | 8, 7.3 (3.7 to 13.7), 10.7 (5.5 to 20.2) | 10, 20 (11.2 to 33), 29.5 (16.6 to 48.8) | 0, 0 (0 to 16.8), 0 (0 to 24.8) | 0, 0 (0 to 11.4), 0 (0 to 16.8) | 0, 0 (0 to 12.9), 0 (0 to 19) | 0, 0 (0 to 20.4), 0 (0 to 30.1) |
| **Rectus** | 15, 2.4 (1.4 to 3.9), 2.6 (1.6 to 4.2) | 5, 1.3 (0.6 to 3), 1.4 (0.6 to 3.3) | 7, 6.4 (3.1 to 12.6), 7 (3.4 to 13.7) | 3, 6 (2.1 to 16.2), 6.6 (2.3 to 17.8) | 0, 0 (0 to 16.8), 0 (0 to 18.4) | 0, 0 (0 to 11.4), 0 (0 to 12.4) | 0, 0 (0 to 12.9), 0 (0 to 14.1) | 0, 0 (0 to 20.4), 0 (0 to 22.3) |
| **Rostral** | 12, 1.9 (1.1 to 3.3), 5.7 (3.3 to 9.9) | 5, 1.3 (0.6 to 3), 3.9 (1.7 to 9.1) | 4, 3.6 (1.4 to 9), 11 (4.3 to 27.3) | 3, 6 (2.1 to 16.2), 18.2 (6.3 to 49.2) | 0, 0 (0 to 16.8), 0 (0 to 51.1) | 0, 0 (0 to 11.4), 0 (0 to 34.5) | 0, 0 (0 to 12.9), 0 (0 to 39.1) | 0, 0 (0 to 20.4), 0 (0 to 61.9) |
| **Precentral** | 26, 4.1 (2.8 to 5.9), 1.1 (0.8 to 1.7) | 16, 4.2 (2.6 to 6.6), 1.2 (0.7 to 1.9) | 6, 5.5 (2.5 to 11.4), 1.5 (0.7 to 3.2) | 3, 6 (2.1 to 16.2), 1.7 (0.6 to 4.5) | 0, 0 (0 to 16.8), 0 (0 to 4.7) | 0, 0 (0 to 11.4), 0 (0 to 3.2) | 1, 3.8 (0.7 to 18.9), 1.1 (0.2 to 5.3) | 0, 0 (0 to 20.4), 0 (0 to 5.7) |
| **Postcentral** | 26, 4.1 (2.8 to 5.9), 2 (1.4 to 2.9) | 16, 4.2 (2.6 to 6.6), 2 (1.3 to 3.3) | 5, 4.5 (2 to 10.2), 2.2 (1 to 5) | 3, 6 (2.1 to 16.2), 2.9 (1 to 8) | 0, 0 (0 to 16.8), 0 (0 to 8.3) | 0, 0 (0 to 11.4), 0 (0 to 5.6) | 2, 7.7 (2.1 to 24.1), 3.8 (1 to 11.8) | 0, 0 (0 to 20.4), 0 (0 to 10) |
| **Paracentral lobule** | 19, 3 (1.9 to 4.6), 2.3 (1.5 to 3.6) | 13, 3.4 (2 to 5.7), 2.6 (1.5 to 4.4) | 6, 5.5 (2.5 to 11.4), 4.2 (2 to 8.8) | 0, 0 (0 to 7.1), 0 (0 to 5.5) | 0, 0 (0 to 16.8), 0 (0 to 13.1) | 0, 0 (0 to 11.4), 0 (0 to 8.8) | 0, 0 (0 to 12.9), 0 (0 to 10) | 0, 0 (0 to 20.4), 0 (0 to 15.8) |
| **Subcentral** | 30, 4.7 (3.3 to 6.7), 8.5 (6 to 12) | 19, 4.9 (3.2 to 7.6), 8.9 (5.7 to 13.7) | 8, 7.3 (3.7 to 13.7), 13.1 (6.7 to 24.8) | 3, 6 (2.1 to 16.2), 10.8 (3.7 to 29.3) | 0, 0 (0 to 16.8), 0 (0 to 30.4) | 0, 0 (0 to 11.4), 0 (0 to 20.5) | 0, 0 (0 to 12.9), 0 (0 to 23.3) | 0, 0 (0 to 20.4), 0 (0 to 36.8) |
| **SPL** | 24, 3.8 (2.6 to 5.6), 1.2 (0.8 to 1.8) | 19, 4.9 (3.2 to 7.6), 1.6 (1 to 2.5) | 3, 2.7 (0.9 to 7.7), 0.9 (0.3 to 2.5) | 1, 2 (0.4 to 10.5), 0.7 (0.1 to 3.4) | 1, 5.3 (0.9 to 24.6), 1.7 (0.3 to 8.1) | 0, 0 (0 to 11.4), 0 (0 to 3.7) | 0, 0 (0 to 12.9), 0 (0 to 4.2) | 0, 0 (0 to 20.4), 0 (0 to 6.7) |
| **SMG** | 55, 8.7 (6.7 to 11.1), 3.1 (2.4 to 4) | 40, 10.4 (7.7 to 13.8), 3.7 (2.8 to 4.9) | 6, 5.5 (2.5 to 11.4), 1.9 (0.9 to 4.1) | 6, 12 (5.6 to 23.8), 4.3 (2 to 8.5) | 3, 15.8 (5.5 to 37.6), 5.6 (2 to 13.4) | 0, 0 (0 to 11.4), 0 (0 to 4) | 0, 0 (0 to 12.9), 0 (0 to 4.6) | 0, 0 (0 to 20.4), 0 (0 to 7.3) |
| **ANG** | 37, 5.8 (4.3 to 7.9), 1.9 (1.4 to 2.5) | 30, 7.8 (5.5 to 10.9), 2.5 (1.8 to 3.5) | 4, 3.6 (1.4 to 9), 1.2 (0.5 to 2.9) | 1, 2 (0.4 to 10.5), 0.6 (0.1 to 3.3) | 1, 5.3 (0.9 to 24.6), 1.7 (0.3 to 7.8) | 1, 3.3 (0.6 to 16.7), 1.1 (0.2 to 5.3) | 0, 0 (0 to 12.9), 0 (0 to 4.1) | 0, 0 (0 to 20.4), 0 (0 to 6.5) |
| **Precuneus** | 38, 6 (4.4 to 8.1), 2.1 (1.6 to 2.9) | 29, 7.5 (5.3 to 10.6), 2.7 (1.9 to 3.8) | 7, 6.4 (3.1 to 12.6), 2.3 (1.1 to 4.5) | 0, 0 (0 to 7.1), 0 (0 to 2.5) | 1, 5.3 (0.9 to 24.6), 1.9 (0.3 to 8.7) | 0, 0 (0 to 11.4), 0 (0 to 4) | 1, 3.8 (0.7 to 18.9), 1.4 (0.2 to 6.7) | 0, 0 (0 to 20.4), 0 (0 to 7.2) |
| **Cuneus** | 23, 3.6 (2.4 to 5.4), 3.9 (2.6 to 5.8) | 20, 5.2 (3.4 to 7.9), 5.6 (3.7 to 8.6) | 2, 1.8 (0.5 to 6.4), 2 (0.5 to 6.9) | 0, 0 (0 to 7.1), 0 (0 to 7.7) | 1, 5.3 (0.9 to 24.6), 5.7 (1 to 26.7) | 0, 0 (0 to 11.4), 0 (0 to 12.3) | 0, 0 (0 to 12.9), 0 (0 to 14) | 0, 0 (0 to 20.4), 0 (0 to 22.1) |
| **O1** | 15, 2.4 (1.4 to 3.9), 2.8 (1.7 to 4.6) | 14, 3.6 (2.2 to 6), 4.3 (2.6 to 7.2) | 0, 0 (0 to 3.4), 0 (0 to 4) | 0, 0 (0 to 7.1), 0 (0 to 8.5) | 1, 5.3 (0.9 to 24.6), 6.3 (1.1 to 29.5) | 0, 0 (0 to 11.4), 0 (0 to 13.6) | 0, 0 (0 to 12.9), 0 (0 to 15.4) | 0, 0 (0 to 20.4), 0 (0 to 24.4) |
| **O2** | 25, 3.9 (2.7 to 5.7), 2.7 (1.9 to 4) | 22, 5.7 (3.8 to 8.5), 4 (2.6 to 5.9) | 1, 0.9 (0.2 to 5), 0.6 (0.1 to 3.4) | 1, 2 (0.4 to 10.5), 1.4 (0.2 to 7.3) | 1, 5.3 (0.9 to 24.6), 3.7 (0.6 to 17.1) | 0, 0 (0 to 11.4), 0 (0 to 7.9) | 0, 0 (0 to 12.9), 0 (0 to 8.9) | 0, 0 (0 to 20.4), 0 (0 to 14.1) |
| **O3** | 9, 1.4 (0.7 to 2.7), 1.7 (0.9 to 3.2) | 8, 2.1 (1.1 to 4), 2.5 (1.2 to 4.8) | 0, 0 (0 to 3.4), 0 (0 to 4) | 0, 0 (0 to 7.1), 0 (0 to 8.4) | 1, 5.3 (0.9 to 24.6), 6.2 (1.1 to 29.1) | 0, 0 (0 to 11.4), 0 (0 to 13.4) | 0, 0 (0 to 12.9), 0 (0 to 15.2) | 0, 0 (0 to 20.4), 0 (0 to 24.1) |
| **Occipital pole** | 2, 0.3 (0.1 to 1.1), 0.3 (0.1 to 1.1) | 2, 0.5 (0.1 to 1.9), 0.5 (0.1 to 1.8) | 0, 0 (0 to 3.4), 0 (0 to 3.2) | 0, 0 (0 to 7.1), 0 (0 to 6.9) | 0, 0 (0 to 16.8), 0 (0 to 16.2) | 0, 0 (0 to 11.4), 0 (0 to 10.9) | 0, 0 (0 to 12.9), 0 (0 to 12.4) | 0, 0 (0 to 20.4), 0 (0 to 19.6) |

| | | | | | | | | |
|---|---|---|---|---|---|---|---|---|
| **Lingual** | 19, 3 (1.9 to 4.6), 1.6 (1 to 2.5) | 17, 4.4 (2.8 to 7), 2.4 (1.5 to 3.7) | 0, 0 (0 to 3.4), 0 (0 to 1.8) | 0, 0 (0 to 7.1), 0 (0 to 3.8) | 2, 10.5 (2.9 to 31.4), 5.7 (1.6 to 16.9) | 0, 0 (0 to 11.4), 0 (0 to 6.1) | 0, 0 (0 to 12.9), 0 (0 to 6.9) | 0, 0 (0 to 20.4), 0 (0 to 11) |
| **Fusiform** | 43, 6.8 (5.1 to 9), 3.3 (2.5 to 4.4) | 33, 8.6 (6.2 to 11.8), 4.2 (3 to 5.8) | 6, 5.5 (2.5 to 11.4), 2.7 (1.2 to 5.6) | 1, 2 (0.4 to 10.5), 1 (0.2 to 5.2) | 2, 10.5 (2.9 to 31.4), 5.2 (1.4 to 15.5) | 0, 0 (0 to 11.4), 0 (0 to 5.6) | 1, 3.8 (0.7 to 18.9), 1.9 (0.3 to 9.3) | 0, 0 (0 to 20.4), 0 (0 to 10) |
| **Temporal pole** | 45, 7.1 (5.3 to 9.4), 3.1 (2.3 to 4) | 26, 6.8 (4.6 to 9.7), 2.9 (2 to 4.2) | 8, 7.3 (3.7 to 13.7), 3.1 (1.6 to 5.9) | 10, 20 (11.2 to 33), 8.6 (4.9 to 14.3) | 1, 5.3 (0.9 to 24.6), 2.3 (0.4 to 10.6) | 0, 0 (0 to 11.4), 0 (0 to 4.9) | 0, 0 (0 to 12.9), 0 (0 to 5.6) | 0, 0 (0 to 20.4), 0 (0 to 8.8) |
| **T1** | 61, 9.6 (7.6 to 12.1), 3.9 (3 to 4.9) | 49, 12.7 (9.8 to 16.4), 5.1 (3.9 to 6.6) | 7, 6.4 (3.1 to 12.6), 2.6 (1.3 to 5) | 2, 4 (1.1 to 13.5), 1.6 (0.4 to 5.4) | 2, 10.5 (2.9 to 31.4), 4.2 (1.2 to 12.6) | 0, 0 (0 to 11.4), 0 (0 to 4.6) | 1, 3.8 (0.7 to 18.9), 1.5 (0.3 to 7.6) | 0, 0 (0 to 20.4), 0 (0 to 8.2) |
| **T2** | 62, 9.8 (7.7 to 12.3), 4.5 (3.5 to 5.7) | 53, 13.8 (10.7 to 17.6), 6.3 (4.9 to 8.1) | 5, 4.5 (2 to 10.2), 2.1 (0.9 to 4.7) | 1, 2 (0.4 to 10.5), 0.9 (0.2 to 4.8) | 0, 0 (0 to 16.8), 0 (0 to 7.7) | 0, 0 (0 to 11.4), 0 (0 to 5.2) | 3, 11.5 (4 to 29), 5.3 (1.8 to 13.3) | 0, 0 (0 to 20.4), 0 (0 to 9.4) |
| **T3** | 44, 6.9 (5.2 to 9.2), 15.2 (11.4 to 20.1) | 39, 10.1 (7.5 to 13.5), 22.1 (16.4 to 29.6) | 1, 0.9 (0.2 to 5), 2 (0.4 to 10.9) | 2, 4 (1.1 to 13.5), 8.7 (2.4 to 29.4) | 1, 5.3 (0.9 to 24.6), 11.5 (2 to 53.9) | 0, 0 (0 to 11.4), 0 (0 to 24.8) | 1, 3.8 (0.7 to 18.9), 8.4 (1.5 to 41.3) | 0, 0 (0 to 20.4), 0 (0 to 44.6) |
| **Planum temporale** | 7, 1.1 (0.5 to 2.3), 3.9 (1.9 to 8) | 5, 1.3 (0.6 to 3), 4.6 (2 to 10.7) | 1, 0.9 (0.2 to 5), 3.2 (0.6 to 17.7) | 1, 2 (0.4 to 10.5), 7.1 (1.3 to 37.4) | 0, 0 (0 to 16.8), 0 (0 to 59.9) | 0, 0 (0 to 11.4), 0 (0 to 40.5) | 0, 0 (0 to 12.9), 0 (0 to 45.9) | 0, 0 (0 to 20.4), 0 (0 to 72.7) |
| **Planum polare** | 21, 3.3 (2.2 to 5), 3.6 (2.4 to 5.5) | 9, 2.3 (1.2 to 4.4), 2.6 (1.4 to 4.8) | 6, 5.5 (2.5 to 11.4), 6 (2.8 to 12.5) | 6, 12 (5.6 to 23.8), 13.2 (6.2 to 26.1) | 0, 0 (0 to 16.8), 0 (0 to 18.5) | 0, 0 (0 to 11.4), 0 (0 to 12.5) | 0, 0 (0 to 12.9), 0 (0 to 14.1) | 0, 0 (0 to 20.4), 0 (0 to 22.4) |
| **Short insular** | 49, 7.7 (5.9 to 10.1), 5.5 (4.2 to 7.1) | 22, 5.7 (3.8 to 8.5), 4 (2.7 to 6) | 15, 13.6 (8.4 to 21.3), 9.6 (6 to 15) | 12, 24 (14.3 to 37.4), 17 (10.1 to 26.4) | 0, 0 (0 to 16.8), 0 (0 to 11.9) | 0, 0 (0 to 11.4), 0 (0 to 8) | 0, 0 (0 to 12.9), 0 (0 to 9.1) | 0, 0 (0 to 20.4), 0 (0 to 14.4) |
| **Long insular** | 45, 7.1 (5.3 to 9.4), 8.5 (6.4 to 11.2) | 25, 6.5 (4.4 to 9.4), 7.8 (5.3 to 11.3) | 11, 10 (5.7 to 17), 12 (6.8 to 20.4) | 9, 18 (9.8 to 30.8), 21.6 (11.7 to 37) | 0, 0 (0 to 16.8), 0 (0 to 20.2) | 0, 0 (0 to 11.4), 0 (0 to 13.6) | 0, 0 (0 to 12.9), 0 (0 to 15.5) | 0, 0 (0 to 20.4), 0 (0 to 24.5) |
| **SCA** | 19, 3 (1.9 to 4.6), 13.8 (8.9 to 21.3) | 6, 1.6 (0.7 to 3.4), 7.2 (3.3 to 15.5) | 8, 7.3 (3.7 to 13.7), 33.5 (17.2 to 63.1) | 5, 10 (4.3 to 21.4), 46.1 (20 to 98.4) | 0, 0 (0 to 16.8), 0 (0 to 77.5) | 0, 0 (0 to 11.4), 0 (0 to 52.3) | 0, 0 (0 to 12.9), 0 (0 to 59.3) | 0, 0 (0 to 20.4), 0 (0 to 93.9) |
| **Cingulate anterior** | 38, 6 (4.4 to 8.1), 4.5 (3.3 to 6.2) | 22, 5.7 (3.8 to 8.5), 4.3 (2.9 to 6.5) | 6, 5.5 (2.5 to 11.4), 4.1 (1.9 to 8.6) | 9, 18 (9.8 to 30.8), 13.7 (7.4 to 23.4) | 0, 0 (0 to 16.8), 0 (0 to 12.8) | 1, 3.3 (0.6 to 16.7), 2.5 (0.4 to 12.7) | 0, 0 (0 to 12.9), 0 (0 to 9.8) | 0, 0 (0 to 20.4), 0 (0 to 15.5) |
| **Cingulate middle** | 28, 4.4 (3.1 to 6.3), 2.8 (2 to 4) | 18, 4.7 (3 to 7.3), 3 (1.9 to 4.6) | 4, 3.6 (1.4 to 9), 2.3 (0.9 to 5.7) | 5, 10 (4.3 to 21.4), 6.4 (2.8 to 13.7) | 1, 5.3 (0.9 to 24.6), 3.4 (0.6 to 15.8) | 0, 0 (0 to 11.4), 0 (0 to 7.3) | 0, 0 (0 to 12.9), 0 (0 to 8.2) | 0, 0 (0 to 20.4), 0 (0 to 13) |
| **Cingulate posterior** | 32, 5 (3.6 to 7), 2.7 (2 to 3.8) | 24, 6.2 (4.2 to 9.1), 3.4 (2.3 to 4.9) | 5, 4.5 (2 to 10.2), 2.5 (1.1 to 5.5) | 2, 4 (1.1 to 13.5), 2.2 (0.6 to 7.3) | 1, 5.3 (0.9 to 24.6), 2.9 (0.5 to 13.4) | 0, 0 (0 to 11.4), 0 (0 to 6.2) | 0, 0 (0 to 12.9), 0 (0 to 7) | 0, 0 (0 to 20.4), 0 (0 to 11.1) |
| **PHG** | 49, 7.7 (5.9 to 10.1), 8.1 (6.2 to 10.5) | 25, 6.5 (4.4 to 9.4), 6.8 (4.6 to 9.8) | 17, 15.5 (9.9 to 23.4), 16.2 (10.3 to 24.4) | 4, 8 (3.2 to 18.8), 8.4 (3.3 to 19.7) | 3, 15.8 (5.5 to 37.6), 16.5 (5.8 to 39.3) | 0, 0 (0 to 11.4), 0 (0 to 11.9) | 0, 0 (0 to 12.9), 0 (0 to 13.5) | 0, 0 (0 to 20.4), 0 (0 to 21.3) |

| | | | | | | | | |
|---|---|---|---|---|---|---|---|---|
| **Hippocampus** | 63, 9.9 (7.8 to 12.5), 13.3 (10.5 to 16.8) | 36, 9.4 (6.8 to 12.7), 12.6 (9.2 to 17) | 19, 17.3 (11.3 to 25.4), 23.2 (15.2 to 34.1) | 6, 12 (5.6 to 23.8), 16.1 (7.5 to 32) | 2, 10.5 (2.9 to 31.4), 14.1 (3.9 to 42.2) | 0, 0 (0 to 11.4), 0 (0 to 15.2) | 0, 0 (0 to 12.9), 0 (0 to 17.3) | 0, 0 (0 to 20.4), 0 (0 to 27.4) |
| **Amygdala** | 50, 7.9 (6 to 10.2), 27.5 (21 to 35.7) | 22, 5.7 (3.8 to 8.5), 19.9 (13.3 to 29.7) | 19, 17.3 (11.3 to 25.4), 60.3 (39.6 to 88.7) | 8, 16 (8.3 to 28.5), 55.9 (29.1 to 99.5) | 1, 5.3 (0.9 to 24.6), 18.4 (3.3 to 86) | 0, 0 (0 to 11.4), 0 (0 to 39.6) | 0, 0 (0 to 12.9), 0 (0 to 44.9) | 0, 0 (0 to 20.4), 0 (0 to 71.2) |

### Supplemental Table 11. Gyral patterns of metastases subentities

The involvement of the gyral segments by metastases in total and by organ of origin is given in absolute numbers (n), relative tumor prevalence (RTP) and relative tumor density (RTD). *ANG = angular gyrus; F1 = superior frontal gyrus; F2 = middle frontal gyrus; F3 = inferior frontal gyrus; GIT = gastrointestinal tract; O1 = superior occipital gyrus; O2 = middle occipital gyrus; O3 = inferior occipital gyrus; PHG = parahippocampal gyrus; SCA = subcallosal area; SMG = supramarginal gyrus; SPL = superior parietal lobule; T1 = superior temporal gyrus; T2 = middle temporal gyrus; T3 = inferior temporal gyrus; UGT = urogenital tract.*

| | Total (n, RTP [95% CI], RTD [95% CI]) | Lung (n, RTP [95% CI], RTD [95% CI]) | Skin (n, RTP [95% CI], RTD [95% CI]) | GIT (n, RTP [95% CI], RTD [95% CI]) | Breast (n, RTP [95% CI], RTD [95% CI]) | UGT (n, RTP [95% CI], RTD [95% CI]) | Miscellaneous (n, RTP [95% CI], RTD [95% CI]) |
|---|---|---|---|---|---|---|---|
| **Total** | 299, 100 (98.7 to 100), 1 (1 to 1) | 143, 100 (97.4 to 100), 1 (1 to 1) | 49, 100 (92.7 to 100), 1 (0.9 to 1) | 37, 100 (90.6 to 100), 1 (0.9 to 1) | 28, 100 (87.9 to 100), 1 (0.9 to 1) | 24, 100 (86.2 to 100), 1 (0.9 to 1) | 18, 100 (82.4 to 100), 1 (0.8 to 1) |
| **Frontal pole** | 4, 1.3 (0.5 to 3.4), 3.1 (1.2 to 7.9) | 1, 0.7 (0.1 to 3.9), 1.6 (0.3 to 8.9) | 1, 2 (0.4 to 10.7), 4.7 (0.8 to 24.8) | 1, 2.7 (0.5 to 13.8), 6.3 (1.1 to 32.1) | 0, 0 (0 to 12.1), 0 (0 to 28) | 0, 0 (0 to 13.8), 0 (0 to 32) | 1, 5.6 (1 to 25.8), 12.9 (2.3 to 59.8) |
| **F1** | 42, 14 (10.6 to 18.4), 2.2 (1.7 to 2.9) | 26, 18.2 (12.7 to 25.3), 2.9 (2 to 4) | 6, 12.2 (5.7 to 24.2), 1.9 (0.9 to 3.9) | 2, 5.4 (1.5 to 17.7), 0.9 (0.2 to 2.8) | 2, 7.1 (2 to 22.6), 1.1 (0.3 to 3.6) | 4, 16.7 (6.7 to 35.9), 2.7 (1.1 to 5.7) | 2, 11.1 (3.1 to 32.8), 1.8 (0.5 to 5.2) |
| **F2** | 46, 15.4 (11.7 to 19.9), 2.9 (2.2 to 3.7) | 22, 15.4 (10.4 to 22.2), 2.9 (1.9 to 4.2) | 4, 8.2 (3.2 to 19.2), 1.5 (0.6 to 3.6) | 5, 13.5 (5.9 to 28), 2.5 (1.1 to 5.2) | 3, 10.7 (3.7 to 27.2), 2 (0.7 to 5.1) | 9, 37.5 (21.2 to 57.3), 7 (4 to 10.7) | 3, 16.7 (5.8 to 39.2), 3.1 (1.1 to 7.3) |
| **F3 orbital** | 2, 0.7 (0.2 to 2.4), 1.9 (0.5 to 6.7) | 0, 0 (0 to 2.6), 0 (0 to 7.3) | 1, 2 (0.4 to 10.7), 5.7 (1 to 29.9) | 0, 0 (0 to 9.4), 0 (0 to 26.3) | 0, 0 (0 to 12.1), 0 (0 to 33.7) | 1, 4.2 (0.7 to 20.2), 11.6 (2.1 to 56.5) | 0, 0 (0 to 17.6), 0 (0 to 49.1) |
| **F3 triangular** | 7, 2.3 (1.1 to 4.8), 2.2 (1.1 to 4.4) | 2, 1.4 (0.4 to 5), 1.3 (0.4 to 4.6) | 4, 8.2 (3.2 to 19.2), 7.6 (3 to 17.9) | 1, 2.7 (0.5 to 13.8), 2.5 (0.4 to 12.9) | 0, 0 (0 to 12.1), 0 (0 to 11.3) | 0, 0 (0 to 13.8), 0 (0 to 12.9) | 0, 0 (0 to 17.6), 0 (0 to 16.4) |
| **F3 opercular** | 5, 1.7 (0.7 to 3.9), 1.3 (0.6 to 3) | 2, 1.4 (0.4 to 5), 1.1 (0.3 to 3.9) | 1, 2 (0.4 to 10.7), 1.6 (0.3 to 8.4) | 1, 2.7 (0.5 to 13.8), 2.1 (0.4 to 10.8) | 1, 3.6 (0.6 to 17.7), 2.8 (0.5 to 13.9) | 0, 0 (0 to 13.8), 0 (0 to 10.8) | 0, 0 (0 to 17.6), 0 (0 to 13.8) |
| **Anterior orbital** | 1, 0.3 (0.1 to 1.9), 1.1 (0.2 to 6.1) | 1, 0.7 (0.1 to 3.9), 2.3 (0.4 to 12.6) | 0, 0 (0 to 7.3), 0 (0 to 23.8) | 0, 0 (0 to 9.4), 0 (0 to 30.8) | 0, 0 (0 to 12.1), 0 (0 to 39.4) | 0, 0 (0 to 13.8), 0 (0 to 45.1) | 0, 0 (0 to 17.6), 0 (0 to 57.5) |
| **Medial orbital** | 2, 0.7 (0.2 to 2.4), 1 (0.3 to 3.5) | 1, 0.7 (0.1 to 3.9), 1 (0.2 to 5.6) | 1, 2 (0.4 to 10.7), 3 (0.5 to 15.5) | 0, 0 (0 to 9.4), 0 (0 to 13.6) | 0, 0 (0 to 12.1), 0 (0 to 17.5) | 0, 0 (0 to 13.8), 0 (0 to 20) | 0, 0 (0 to 17.6), 0 (0 to 25.5) |
| **Lateral orbital** | 3, 1 (0.3 to 2.9), 1.8 (0.6 to 5.3) | 1, 0.7 (0.1 to 3.9), 1.3 (0.2 to 7) | 0, 0 (0 to 7.3), 0 (0 to 13.1) | 0, 0 (0 to 9.4), 0 (0 to 17) | 1, 3.6 (0.6 to 17.7), 6.5 (1.1 to 32) | 0, 0 (0 to 13.8), 0 (0 to 24.9) | 1, 5.6 (1 to 25.8), 10 (1.8 to 46.5) |
| **Posterior orbital** | 1, 0.3 (0.1 to 1.9), 0.5 (0.1 to 2.8) | 1, 0.7 (0.1 to 3.9), 1 (0.2 to 5.7) | 0, 0 (0 to 7.3), 0 (0 to 10.7) | 0, 0 (0 to 9.4), 0 (0 to 13.9) | 0, 0 (0 to 12.1), 0 (0 to 17.8) | 0, 0 (0 to 13.8), 0 (0 to 20.4) | 0, 0 (0 to 17.6), 0 (0 to 26) |

| | | | | | | | |
|---|---|---|---|---|---|---|---|
| **Rectus** | 0, 0 (0 to 1.3), 0 (0 to 1.4) | 0, 0 (0 to 2.6), 0 (0 to 2.9) | 0, 0 (0 to 7.3), 0 (0 to 8) | 0, 0 (0 to 9.4), 0 (0 to 10.3) | 0, 0 (0 to 12.1), 0 (0 to 13.2) | 0, 0 (0 to 13.8), 0 (0 to 15.1) | 0, 0 (0 to 17.6), 0 (0 to 19.3) |
| **Rostral** | 1, 0.3 (0.1 to 1.9), 1 (0.2 to 5.7) | 0, 0 (0 to 2.6), 0 (0 to 7.9) | 1, 2 (0.4 to 10.7), 6.2 (1.1 to 32.5) | 0, 0 (0 to 9.4), 0 (0 to 28.6) | 0, 0 (0 to 12.1), 0 (0 to 36.6) | 0, 0 (0 to 13.8), 0 (0 to 41.9) | 0, 0 (0 to 17.6), 0 (0 to 53.4) |
| **Precentral** | 32, 10.7 (7.7 to 14.7), 3 (2.1 to 4.1) | 17, 11.9 (7.6 to 18.2), 3.3 (2.1 to 5.1) | 4, 8.2 (3.2 to 19.2), 2.3 (0.9 to 5.3) | 5, 13.5 (5.9 to 28), 3.8 (1.6 to 7.8) | 2, 7.1 (2 to 22.6), 2 (0.6 to 6.3) | 3, 12.5 (4.3 to 31), 3.5 (1.2 to 8.6) | 1, 5.6 (1 to 25.8), 1.5 (0.3 to 7.2) |
| **Postcentral** | 18, 6 (3.8 to 9.3), 3 (1.9 to 4.6) | 11, 7.7 (4.3 to 13.2), 3.8 (2.1 to 6.5) | 2, 4.1 (1.1 to 13.7), 2 (0.6 to 6.7) | 2, 5.4 (1.5 to 17.7), 2.7 (0.7 to 8.7) | 1, 3.6 (0.6 to 17.7), 1.8 (0.3 to 8.7) | 1, 4.2 (0.7 to 20.2), 2 (0.4 to 9.9) | 1, 5.6 (1 to 25.8), 2.7 (0.5 to 12.6) |
| **Paracentral lobule** | 10, 3.3 (1.8 to 6), 2.6 (1.4 to 4.7) | 4, 2.8 (1.1 to 7), 2.2 (0.8 to 5.4) | 1, 2 (0.4 to 10.7), 1.6 (0.3 to 8.3) | 1, 2.7 (0.5 to 13.8), 2.1 (0.4 to 10.7) | 1, 3.6 (0.6 to 17.7), 2.8 (0.5 to 13.8) | 2, 8.3 (2.3 to 25.8), 6.5 (1.8 to 20.1) | 1, 5.6 (1 to 25.8), 4.3 (0.8 to 20) |
| **Subcentral** | 6, 2 (0.9 to 4.3), 3.6 (1.7 to 7.8) | 1, 0.7 (0.1 to 3.9), 1.3 (0.2 to 7) | 3, 6.1 (2.1 to 16.5), 11.1 (3.8 to 29.9) | 1, 2.7 (0.5 to 13.8), 4.9 (0.9 to 25) | 0, 0 (0 to 12.1), 0 (0 to 21.8) | 0, 0 (0 to 13.8), 0 (0 to 24.9) | 1, 5.6 (1 to 25.8), 10 (1.8 to 46.5) |
| **SPL** | 17, 5.7 (3.6 to 8.9), 1.9 (1.2 to 2.9) | 9, 6.3 (3.3 to 11.5), 2.1 (1.1 to 3.8) | 2, 4.1 (1.1 to 13.7), 1.3 (0.4 to 4.5) | 2, 5.4 (1.5 to 17.7), 1.8 (0.5 to 5.8) | 3, 10.7 (3.7 to 27.2), 3.5 (1.2 to 8.9) | 0, 0 (0 to 13.8), 0 (0 to 4.5) | 1, 5.6 (1 to 25.8), 1.8 (0.3 to 8.4) |
| **SMG** | 13, 4.3 (2.6 to 7.3), 1.5 (0.9 to 2.6) | 3, 2.1 (0.7 to 6), 0.7 (0.3 to 2.1) | 4, 8.2 (3.2 to 19.2), 2.9 (1.1 to 6.8) | 2, 5.4 (1.5 to 17.7), 1.9 (0.5 to 6.3) | 2, 7.1 (2 to 22.6), 2.5 (0.7 to 8.1) | 1, 4.2 (0.7 to 20.2), 1.5 (0.3 to 7.2) | 1, 5.6 (1 to 25.8), 2 (0.4 to 9.2) |
| **ANG** | 11, 3.7 (2.1 to 6.5), 1.2 (0.7 to 2.1) | 6, 4.2 (1.9 to 8.9), 1.3 (0.6 to 2.8) | 1, 2 (0.4 to 10.7), 0.6 (0.1 to 3.4) | 0, 0 (0 to 9.4), 0 (0 to 3) | 1, 3.6 (0.6 to 17.7), 1.1 (0.2 to 5.6) | 2, 8.3 (2.3 to 25.8), 2.7 (0.7 to 8.2) | 1, 5.6 (1 to 25.8), 1.8 (0.3 to 8.2) |
| **Precuneus** | 20, 6.7 (4.4 to 10.1), 2.4 (1.6 to 3.6) | 6, 4.2 (1.9 to 8.9), 1.5 (0.7 to 3.1) | 6, 12.2 (5.7 to 24.2), 4.3 (2 to 8.6) | 2, 5.4 (1.5 to 17.7), 1.9 (0.5 to 6.3) | 2, 7.1 (2 to 22.6), 2.5 (0.7 to 8) | 2, 8.3 (2.3 to 25.8), 3 (0.8 to 9.2) | 2, 11.1 (3.1 to 32.8), 3.9 (1.1 to 11.6) |
| **Cuneus** | 14, 4.7 (2.8 to 7.7), 5.1 (3 to 8.4) | 6, 4.2 (1.9 to 8.9), 4.6 (2.1 to 9.6) | 3, 6.1 (2.1 to 16.5), 6.6 (2.3 to 17.9) | 3, 8.1 (2.8 to 21.3), 8.8 (3 to 23.1) | 1, 3.6 (0.6 to 17.7), 3.9 (0.7 to 19.2) | 1, 4.2 (0.7 to 20.2), 4.5 (0.8 to 22) | 0, 0 (0 to 17.6), 0 (0 to 19.1) |
| **O1** | 6, 2 (0.9 to 4.3), 2.4 (1.1 to 5.2) | 3, 2.1 (0.7 to 6), 2.5 (0.9 to 7.2) | 0, 0 (0 to 7.3), 0 (0 to 8.7) | 2, 5.4 (1.5 to 17.7), 6.5 (1.8 to 21.2) | 1, 3.6 (0.6 to 17.7), 4.3 (0.8 to 21.2) | 0, 0 (0 to 13.8), 0 (0 to 16.5) | 0, 0 (0 to 17.6), 0 (0 to 21) |
| **O2** | 27, 9 (6.3 to 12.8), 6.3 (4.4 to 8.9) | 18, 12.6 (8.1 to 19), 8.7 (5.6 to 13.2) | 5, 10.2 (4.4 to 21.8), 7.1 (3.1 to 15.1) | 3, 8.1 (2.8 to 21.3), 5.6 (1.9 to 14.8) | 0, 0 (0 to 12.1), 0 (0 to 8.4) | 1, 4.2 (0.7 to 20.2), 2.9 (0.5 to 14) | 0, 0 (0 to 17.6), 0 (0 to 12.2) |
| **O3** | 10, 3.3 (1.8 to 6), 4 (2.2 to 7.1) | 3, 2.1 (0.7 to 6), 2.5 (0.8 to 7.1) | 0, 0 (0 to 7.3), 0 (0 to 8.6) | 2, 5.4 (1.5 to 17.7), 6.4 (1.8 to 20.9) | 1, 3.6 (0.6 to 17.7), 4.2 (0.7 to 20.9) | 4, 16.7 (6.7 to 35.9), 19.7 (7.9 to 42.4) | 0, 0 (0 to 17.6), 0 (0 to 20.8) |
| **Occipital pole** | 1, 0.3 (0.1 to 1.9), 0.3 (0.1 to 1.8) | 0, 0 (0 to 2.6), 0 (0 to 2.5) | 0, 0 (0 to 7.3), 0 (0 to 7) | 0, 0 (0 to 9.4), 0 (0 to 9) | 0, 0 (0 to 12.1), 0 (0 to 11.6) | 1, 4.2 (0.7 to 20.2), 4 (0.7 to 19.5) | 0, 0 (0 to 17.6), 0 (0 to 16.9) |
| **Lingual** | 10, 3.3 (1.8 to 6), 1.8 (1 to 3.2) | 5, 3.5 (1.5 to 7.9), 1.9 (0.8 to 4.3) | 2, 4.1 (1.1 to 13.7), 2.2 (0.6 to 7.4) | 1, 2.7 (0.5 to 13.8), 1.5 (0.3 to 7.4) | 2, 7.1 (2 to 22.6), 3.8 (1.1 to 12.2) | 0, 0 (0 to 13.8), 0 (0 to 7.4) | 0, 0 (0 to 17.6), 0 (0 to 9.4) |
| **Fusiform** | 10, 3.3 (1.8 to 6), 1.6 (0.9 to 3) | 5, 3.5 (1.5 to 7.9), 1.7 (0.7 to 3.9) | 0, 0 (0 to 7.3), 0 (0 to 3.6) | 2, 5.4 (1.5 to 17.7), 2.7 (0.7 to 8.7) | 0, 0 (0 to 12.1), 0 (0 to 5.9) | 1, 4.2 (0.7 to 20.2), 2.1 (0.4 to 10) | 2, 11.1 (3.1 to 32.8), 5.5 (1.5 to 16.1) |
| **Temporal pole** | 2, 0.7 (0.2 to 2.4), 0.3 (0.1 to 1) | 2, 1.4 (0.4 to 5), 0.6 (0.2 to 2.1) | 0, 0 (0 to 7.3), 0 (0 to 3.1) | 0, 0 (0 to 9.4), 0 (0 to 4.1) | 0, 0 (0 to 12.1), 0 (0 to 5.2) | 0, 0 (0 to 13.8), 0 (0 to 6) | 0, 0 (0 to 17.6), 0 (0 to 7.6) |

| | | | | | | | |
|---|---|---|---|---|---|---|---|
| **T1** | 8, 2.7 (1.4 to 5.2), 1.1 (0.5 to 2.1) | 3, 2.1 (0.7 to 6), 0.8 (0.3 to 2.4) | 3, 6.1 (2.1 to 16.5), 2.5 (0.8 to 6.6) | 0, 0 (0 to 9.4), 0 (0 to 3.8) | 1, 3.6 (0.6 to 17.7), 1.4 (0.3 to 7.1) | 0, 0 (0 to 13.8), 0 (0 to 5.5) | 1, 5.6 (1 to 25.8), 2.2 (0.4 to 10.3) |
| **T2** | 11, 3.7 (2.1 to 6.5), 1.7 (1 to 3) | 6, 4.2 (1.9 to 8.9), 1.9 (0.9 to 4.1) | 4, 8.2 (3.2 to 19.2), 3.8 (1.5 to 8.8) | 1, 2.7 (0.5 to 13.8), 1.2 (0.2 to 6.4) | 0, 0 (0 to 12.1), 0 (0 to 5.6) | 0, 0 (0 to 13.8), 0 (0 to 6.4) | 0, 0 (0 to 17.6), 0 (0 to 8.1) |
| **T3** | 11, 3.7 (2.1 to 6.5), 8 (4.5 to 14.1) | 5, 3.5 (1.5 to 7.9), 7.6 (3.3 to 17.3) | 3, 6.1 (2.1 to 16.5), 13.4 (4.6 to 36.1) | 0, 0 (0 to 9.4), 0 (0 to 20.6) | 2, 7.1 (2 to 22.6), 15.6 (4.3 to 49.5) | 1, 4.2 (0.7 to 20.2), 9.1 (1.6 to 44.3) | 0, 0 (0 to 17.6), 0 (0 to 38.5) |
| **Planum temporale** | 1, 0.3 (0.1 to 1.9), 1.2 (0.2 to 6.7) | 0, 0 (0 to 2.6), 0 (0 to 9.3) | 0, 0 (0 to 7.3), 0 (0 to 25.9) | 0, 0 (0 to 9.4), 0 (0 to 33.5) | 0, 0 (0 to 12.1), 0 (0 to 43) | 0, 0 (0 to 13.8), 0 (0 to 49.2) | 1, 5.6 (1 to 25.8), 19.8 (3.5 to 91.8) |
| **Planum polare** | 2, 0.7 (0.2 to 2.4), 0.7 (0.2 to 2.6) | 1, 0.7 (0.1 to 3.9), 0.8 (0.1 to 4.2) | 1, 2 (0.4 to 10.7), 2.2 (0.4 to 11.7) | 0, 0 (0 to 9.4), 0 (0 to 10.3) | 0, 0 (0 to 12.1), 0 (0 to 13.2) | 0, 0 (0 to 13.8), 0 (0 to 15.1) | 0, 0 (0 to 17.6), 0 (0 to 19.3) |
| **Short insular** | 1, 0.3 (0.1 to 1.9), 0.2 (0 to 1.3) | 0, 0 (0 to 2.6), 0 (0 to 1.8) | 1, 2 (0.4 to 10.7), 1.4 (0.3 to 7.6) | 0, 0 (0 to 9.4), 0 (0 to 6.6) | 0, 0 (0 to 12.1), 0 (0 to 8.5) | 0, 0 (0 to 13.8), 0 (0 to 9.8) | 0, 0 (0 to 17.6), 0 (0 to 12.4) |
| **Long insular** | 1, 0.3 (0.1 to 1.9), 0.4 (0.1 to 2.2) | 0, 0 (0 to 2.6), 0 (0 to 3.1) | 0, 0 (0 to 7.3), 0 (0 to 8.7) | 1, 2.7 (0.5 to 13.8), 3.2 (0.6 to 16.6) | 0, 0 (0 to 12.1), 0 (0 to 14.5) | 0, 0 (0 to 13.8), 0 (0 to 16.6) | 0, 0 (0 to 17.6), 0 (0 to 21.1) |
| **SCA** | 0, 0 (0 to 1.3), 0 (0 to 5.8) | 0, 0 (0 to 2.6), 0 (0 to 12.1) | 0, 0 (0 to 7.3), 0 (0 to 33.5) | 0, 0 (0 to 9.4), 0 (0 to 43.3) | 0, 0 (0 to 12.1), 0 (0 to 55.6) | 0, 0 (0 to 13.8), 0 (0 to 63.6) | 0, 0 (0 to 17.6), 0 (0 to 81) |
| **Cingulate anterior** | 4, 1.3 (0.5 to 3.4), 1 (0.4 to 2.6) | 2, 1.4 (0.4 to 5), 1.1 (0.3 to 3.8) | 1, 2 (0.4 to 10.7), 1.5 (0.3 to 8.1) | 1, 2.7 (0.5 to 13.8), 2.1 (0.4 to 10.5) | 0, 0 (0 to 12.1), 0 (0 to 9.2) | 0, 0 (0 to 13.8), 0 (0 to 10.5) | 0, 0 (0 to 17.6), 0 (0 to 13.3) |
| **Cingulate middle** | 6, 2 (0.9 to 4.3), 1.3 (0.6 to 2.8) | 4, 2.8 (1.1 to 7), 1.8 (0.7 to 4.5) | 2, 4.1 (1.1 to 13.7), 2.6 (0.7 to 8.8) | 0, 0 (0 to 9.4), 0 (0 to 6) | 0, 0 (0 to 12.1), 0 (0 to 7.7) | 0, 0 (0 to 13.8), 0 (0 to 8.8) | 0, 0 (0 to 17.6), 0 (0 to 11.2) |
| **Cingulate posterior** | 3, 1 (0.3 to 2.9), 0.5 (0.2 to 1.6) | 2, 1.4 (0.4 to 5), 0.8 (0.2 to 2.7) | 0, 0 (0 to 7.3), 0 (0 to 3.9) | 1, 2.7 (0.5 to 13.8), 1.5 (0.3 to 7.5) | 0, 0 (0 to 12.1), 0 (0 to 6.6) | 0, 0 (0 to 13.8), 0 (0 to 7.5) | 0, 0 (0 to 17.6), 0 (0 to 9.6) |
| **PHG** | 2, 0.7 (0.2 to 2.4), 0.7 (0.2 to 2.5) | 0, 0 (0 to 2.6), 0 (0 to 2.7) | 0, 0 (0 to 7.3), 0 (0 to 7.6) | 1, 2.7 (0.5 to 13.8), 2.8 (0.5 to 14.5) | 0, 0 (0 to 12.1), 0 (0 to 12.6) | 1, 4.2 (0.7 to 20.2), 4.4 (0.8 to 21.2) | 0, 0 (0 to 17.6), 0 (0 to 18.4) |
| **Hippocampus** | 3, 1 (0.3 to 2.9), 1.3 (0.5 to 3.9) | 1, 0.7 (0.1 to 3.9), 0.9 (0.2 to 5.2) | 2, 4.1 (1.1 to 13.7), 5.5 (1.5 to 18.4) | 0, 0 (0 to 9.4), 0 (0 to 12.6) | 0, 0 (0 to 12.1), 0 (0 to 16.2) | 0, 0 (0 to 13.8), 0 (0 to 18.5) | 0, 0 (0 to 17.6), 0 (0 to 23.6) |
| **Amygdala** | 2, 0.7 (0.2 to 2.4), 2.3 (0.6 to 8.4) | 1, 0.7 (0.1 to 3.9), 2.4 (0.4 to 13.5) | 1, 2 (0.4 to 10.7), 7.1 (1.3 to 37.3) | 0, 0 (0 to 9.4), 0 (0 to 32.8) | 0, 0 (0 to 12.1), 0 (0 to 42.1) | 0, 0 (0 to 13.8), 0 (0 to 48.2) | 0, 0 (0 to 17.6), 0 (0 to 61.4) |

**Supplemental Table 12. Central supratentorial anatomical patterns of neuroepithelial tumors, primary central nervous system lymphoma and brain metastases**

The involvement of the central supratentorial structures by brain tumors in total, neuroepithelial tumors (NT), primary central nervous system lymphoma (PCNSL) or metastases is given in absolute numbers (n), relative tumor prevalence (RTP) and relative tumor density (RTD).

| | Total (n, RTP [95% CI], RTD [95% CI]) | NT (n, RTP [95% CI], RTD [95% CI]) | PCNSL (n, RTP [95% CI], RTD [95% CI]) | Metastases (n, RTP [95% CI], RTD [95% CI]) |
|---|---|---|---|---|
| **Total** | 1000, 100 (99.6 to 100), 1 (1 to 1) | 657, 100 (99.4 to 100), 1 (1 to 1) | 44, 100 (92 to 100), 1 (0.9 to 1) | 299, 100 (98.7 to 100), 1 (1 to 1) |
| **Putamen** | 14, 1.4 (0.8 to 2.3), 1.4 (0.8 to 2.3) | 5, 0.8 (0.3 to 1.8), 0.7 (0.3 to 1.7) | 6, 13.6 (6.4 to 26.7), 13.2 (6.2 to 25.8) | 3, 1 (0.3 to 2.9), 1 (0.3 to 2.8) |
| **Caudate** | 19, 1.9 (1.2 to 2.9), 2.7 (1.7 to 4.1) | 5, 0.8 (0.3 to 1.8), 1.1 (0.5 to 2.5) | 8, 18.2 (9.5 to 32), 25.5 (13.3 to 44.8) | 6, 2 (0.9 to 4.3), 2.8 (1.3 to 6) |
| **Globus pallidum** | 13, 1.3 (0.8 to 2.2), 4.6 (2.7 to 7.9) | 6, 0.9 (0.4 to 2), 3.3 (1.5 to 7.1) | 6, 13.6 (6.4 to 26.7), 48.6 (22.8 to 95.2) | 1, 0.3 (0.1 to 1.9), 1.2 (0.2 to 6.7) |
| **Internal capsule** | 24, 2.4 (1.6 to 3.5), 2.5 (1.7 to 3.6) | 4, 0.6 (0.2 to 1.6), 0.6 (0.2 to 1.6) | 17, 38.6 (25.7 to 53.4), 39.7 (26.5 to 54.9) | 3, 1 (0.3 to 2.9), 1 (0.4 to 3) |
| **Hypothalamus** | 25, 2.5 (1.7 to 3.7), 3.4 (2.3 to 5) | 19, 2.9 (1.9 to 4.5), 4 (2.6 to 6.1) | 6, 13.6 (6.4 to 26.7), 18.7 (8.8 to 36.6) | 0, 0 (0 to 1.3), 0 (0 to 1.7) |
| **Thalamus** | 54, 5.4 (4.2 to 7), 4 (3.1 to 5.2) | 41, 6.2 (4.6 to 8.4), 4.7 (3.5 to 6.2) | 6, 13.6 (6.4 to 26.7), 10.2 (4.8 to 19.9) | 7, 2.3 (1.1 to 4.8), 1.7 (0.8 to 3.5) |

**Supplemental Table 13. Central supratentorial anatomical patterns of neuroepithelial tumor subentities**

The involvement of the central supratentorial structures by neuroepithelial tumors (NT) in total, glioblastoma (GBM), WHO grade III (gIIIG) and II (gIIG) glioma, developmental tumors (DT), pilocytic astrocytoma (PA), ependymoma (EP), or medulloblastoma (MB) is given in absolute numbers (n), relative tumor prevalence (RTP) and relative tumor density (RTD).

| | Total (n, RTP [95% CI], RTD [95% CI]) | GBM (n, RTP [95% CI], RTD [95% CI]) | gIIIG (n, RTP [95% CI], RTD [95% CI]) | gIIG (n, RTP [95% CI], RTD [95% CI]) | DT (n, RTP [95% CI], RTD [95% CI]) | EP (n, RTP [95% CI], RTD [95% CI]) | PA (n, RTP [95% CI], RTD [95% CI]) | MB (n, RTP [95% CI], RTD [95% CI]) |
|---|---|---|---|---|---|---|---|---|
| **Total** | 635, 100 (99.4 to 100), 1 (1 to 1) | 385, 100 (99 to 100), 1 (1 to 1) | 110, 100 (96.6 to 100), 1 (1 to 1) | 50, 100 (92.9 to 100), 1 (0.9 to 1) | 19, 100 (83.2 to 100), 1 (0.8 to 1) | 26, 100 (87.1 to 100), 1 (0.9 to 1) | 30, 100 (88.6 to 100), 1 (0.9 to 1) | 15, 100 (79.6 to 100), 1 (0.8 to 1) |
| **Putamen** | 5, 0.8 (0.3 to 1.8), 0.8 (0.3 to 1.8) | 4, 1 (0.4 to 2.6), 1 (0.4 to 2.6) | 1, 0.9 (0.2 to 5), 0.9 (0.2 to 4.8) | 0, 0 (0 to 7.1), 0 (0 to 6.9) | 0, 0 (0 to 16.8), 0 (0 to 16.3) | 0, 0 (0 to 12.9), 0 (0 to 12.4) | 0, 0 (0 to 11.4), 0 (0 to 11) | 0, 0 (0 to 20.4), 0 (0 to 19.7) |
| **Caudate** | 5, 0.8 (0.3 to 1.8), 1.1 (0.5 to 2.6) | 3, 0.8 (0.3 to 2.3), 1.1 (0.4 to 3.2) | 1, 0.9 (0.2 to 5), 1.3 (0.2 to 7) | 1, 2 (0.4 to 10.5), 2.8 (0.5 to 14.7) | 0, 0 (0 to 16.8), 0 (0 to 23.6) | 0, 0 (0 to 12.9), 0 (0 to 18) | 0, 0 (0 to 11.4), 0 (0 to 15.9) | 0, 0 (0 to 20.4), 0 (0 to 28.6) |
| **Globus pallidum** | 6, 0.9 (0.4 to 2), 3.4 (1.5 to 7.3) | 4, 1 (0.4 to 2.6), 3.7 (1.4 to 9.4) | 2, 1.8 (0.5 to 6.4), 6.5 (1.8 to 22.8) | 0, 0 (0 to 7.1), 0 (0 to 25.4) | 0, 0 (0 to 16.8), 0 (0 to 60) | 0, 0 (0 to 12.9), 0 (0 to 45.9) | 0, 0 (0 to 11.4), 0 (0 to 40.5) | 0, 0 (0 to 20.4), 0 (0 to 72.7) |
| **Internal capsule** | 4, 0.6 (0.2 to 1.6), 0.6 (0.3 to 1.7) | 4, 1 (0.4 to 2.6), 1.1 (0.4 to 2.7) | 0, 0 (0 to 3.4), 0 (0 to 3.5) | 0, 0 (0 to 7.1), 0 (0 to 7.3) | 0, 0 (0 to 16.8), 0 (0 to 17.3) | 0, 0 (0 to 12.9), 0 (0 to 13.2) | 0, 0 (0 to 11.4), 0 (0 to 11.7) | 0, 0 (0 to 20.4), 0 (0 to 21) |
| **Hypothalamus** | 17, 2.7 (1.7 to 4.2), 3.7 (2.3 to 5.8) | 4, 1 (0.4 to 2.6), 1.4 (0.6 to 3.6) | 4, 3.6 (1.4 to 9), 5 (2 to 12.3) | 3, 6 (2.1 to 16.2), 8.2 (2.8 to 22.2) | 1, 5.3 (0.9 to 24.6), 7.2 (1.3 to 33.8) | 0, 0 (0 to 12.9), 0 (0 to 17.7) | 5, 16.7 (7.3 to 33.6), 22.9 (10.1 to 46) | 0, 0 (0 to 20.4), 0 (0 to 28) |
| **Thalamus** | 40, 6.3 (4.7 to 8.5), 4.7 (3.5 to 6.3) | 23, 6 (4 to 8.8), 4.5 (3 to 6.6) | 7, 6.4 (3.1 to 12.6), 4.7 (2.3 to 9.4) | 9, 18 (9.8 to 30.8), 13.4 (7.3 to 23) | 0, 0 (0 to 16.8), 0 (0 to 12.6) | 1, 3.8 (0.7 to 18.9), 2.9 (0.5 to 14.1) | 0, 0 (0 to 11.4), 0 (0 to 8.5) | 0, 0 (0 to 20.4), 0 (0 to 15.2) |

**Supplemental Table 14. Brainstem patterns of neuroepithelial tumors, primary central nervous system lymphoma and brain metastases**

The involvement of the brainstem by brain tumors in total, neuroepithelial tumors (NT), primary central nervous system lymphoma (PCNSL) or metastases is given in absolute numbers (n), relative tumor prevalence (RTP) and relative tumor density (RTD).

| | **Total (n, RTP [95% CI], RTD [95% CI])** | **NT (n, RTP [95% CI], RTD [95% CI])** | **PCNSL (n, RTP [95% CI], RTD [95% CI])** | **Metastases (n, RTP [95% CI], RTD [95% CI])** |
|---|---|---|---|---|
| **Total** | 1000, 100 (99.6 to 100), 1 (1 to 1) | 657, 100 (99.4 to 100), 1 (1 to 1) | 44, 100 (92 to 100), 1 (0.9 to 1) | 299, 100 (98.7 to 100), 1 (1 to 1) |
| **Mesencephalon** | 31, 3.1 (2.2 to 4.4), 3.4 (2.4 to 4.7) | 18, 2.7 (1.7 to 4.3), 3 (1.9 to 4.7) | 12, 27.3 (16.3 to 41.8), 29.6 (17.8 to 45.5) | 1, 0.3 (0.1 to 1.9), 0.4 (0.1 to 2) |
| **Pons** | 33, 3.3 (2.4 to 4.6), 2.4 (1.7 to 3.3) | 22, 3.3 (2.2 to 5), 2.4 (1.6 to 3.6) | 4, 9.1 (3.6 to 21.2), 6.5 (2.6 to 15.1) | 7, 2.3 (1.1 to 4.8), 1.7 (0.8 to 3.4) |
| **Medulla oblongata** | 13, 1.3 (0.8 to 2.2), 5.2 (3 to 8.8) | 9, 1.4 (0.7 to 2.6), 5.4 (2.9 to 10.2) | 2, 4.5 (1.3 to 15.1), 18 (5 to 60) | 2, 0.7 (0.2 to 2.4), 2.7 (0.7 to 9.5) |

**Supplemental Table 15. Cerebellar patterns of neuroepithelial tumors and brain metastases**

The involvement of the cerebellar structures by brain tumors in total, neuroepithelial tumors (NT) or metastases is given in absolute numbers (n), relative tumor prevalence (RTP) and relative tumor density (RTD).

| | Total (n, RTP [95% CI], RTD [95% CI]) | NT (n, RTP [95% CI], RTD [95% CI]) | Metastases (n, RTP [95% CI], RTD [95% CI]) |
|---|---|---|---|
| **Total** | 1000, 100 (99.6 to 100), 1 (1 to 1) | 657, 100 (99.4 to 100), 1 (1 to 1) | 299, 100 (98.7 to 100), 1 (1 to 1) |
| **Vermis** | 33, 3.3 (2.4 to 4.6), 6 (4.3 to 8.4) | 26, 4 (2.7 to 5.7), 7.2 (5 to 10.5) | 5, 1.7 (0.7 to 3.9), 3.1 (1.3 to 7.1) |
| **Hemisphere** | 122, 12.2 (10.3 to 14.4), 1.2 (1 to 1.4) | 28, 4.3 (3 to 6.1), 0.4 (0.3 to 0.6) | 89, 29.8 (24.9 to 35.2), 2.9 (2.4 to 3.5) |
| **Anterior lobe** | 33, 3.3 (2.4 to 4.6), 1.1 (0.8 to 1.5) | 16, 2.4 (1.5 to 3.9), 0.8 (0.5 to 1.3) | 15, 5 (3.1 to 8.1), 1.7 (1 to 2.7) |
| **Medial lobe** | 40, 4 (3 to 5.4), 1.5 (1.1 to 2) | 14, 2.1 (1.3 to 3.5), 0.8 (0.5 to 1.3) | 23, 7.7 (5.2 to 11.3), 2.9 (2 to 4.3) |
| **Posterior lobe** | 91, 9.1 (7.5 to 11), 1.8 (1.5 to 2.2) | 23, 3.5 (2.3 to 5.2), 0.7 (0.5 to 1) | 64, 21.4 (17.1 to 26.4), 4.3 (3.4 to 5.3) |
| **Flocculonodular lobe** | 20, 2 (1.3 to 3.1), 20.3 (13.2 to 31.2) | 18, 2.7 (1.7 to 4.3), 27.9 (17.7 to 43.6) | 1, 0.3 (0.1 to 1.9), 3.4 (0.6 to 19) |
| **Central** | 14, 1.4 (0.8 to 2.3), 31.5 (18.8 to 52.5) | 11, 1.7 (0.9 to 3), 37.6 (21.1 to 66.8) | 2, 0.7 (0.2 to 2.4), 15 (4.1 to 54.1) |
| **Culmen** | 15, 1.5 (0.9 to 2.5), 7.5 (4.5 to 12.3) | 11, 1.7 (0.9 to 3), 8.3 (4.7 to 14.8) | 3, 1 (0.3 to 2.9), 5 (1.7 to 14.5) |
| **Declive** | 13, 1.3 (0.8 to 2.2), 13.7 (8.1 to 23.4) | 10, 1.5 (0.8 to 2.8), 16.1 (8.8 to 29.4) | 1, 0.3 (0.1 to 1.9), 3.5 (0.6 to 19.8) |
| **Folium** | 11, 1.1 (0.6 to 2), 27.1 (15.2 to 48.3) | 7, 1.1 (0.5 to 2.2), 26.3 (12.8 to 53.9) | 2, 0.7 (0.2 to 2.4), 16.5 (4.5 to 59.4) |
| **Tuber** | 10, 1 (0.5 to 1.8), 28.1 (15.3 to 51.5) | 7, 1.1 (0.5 to 2.2), 30 (14.6 to 61.4) | 1, 0.3 (0.1 to 1.9), 9.4 (1.7 to 52.6) |
| **Pyramid** | 9, 0.9 (0.5 to 1.7), 23.1 (12.2 to 43.6) | 7, 1.1 (0.5 to 2.2), 27.3 (13.3 to 56) | 1, 0.3 (0.1 to 1.9), 8.6 (1.5 to 48) |
| **Uvula** | 13, 1.3 (0.8 to 2.2), 18 (10.5 to 30.6) | 11, 1.7 (0.9 to 3), 23.2 (13 to 41.2) | 1, 0.3 (0.1 to 1.9), 4.6 (0.8 to 25.9) |
| **Nodule** | 19, 1.9 (1.2 to 2.9), 96.8 (62.2 to 150.2) | 17, 2.6 (1.6 to 4.1), 131.9 (82.6 to 209.2) | 1, 0.3 (0.1 to 1.9), 17 (3 to 95.3) |
| **Ala lobuli centralis** | 12, 1.2 (0.7 to 2.1), 1.1 (0.6 to 1.9) | 8, 1.2 (0.6 to 2.4), 1.1 (0.6 to 2.2) | 3, 1 (0.3 to 2.9), 0.9 (0.3 to 2.6) |
| **AQL** | 20, 2 (1.3 to 3.1), 1.2 (0.8 to 1.9) | 10, 1.5 (0.8 to 2.8), 0.9 (0.5 to 1.7) | 8, 2.7 (1.4 to 5.2), 1.6 (0.8 to 3.2) |
| **PQL** | 16, 1.6 (1 to 2.6), 1.3 (0.8 to 2.1) | 7, 1.1 (0.5 to 2.2), 0.9 (0.4 to 1.8) | 4, 1.3 (0.5 to 3.4), 1.1 (0.4 to 2.8) |
| **SSL** | 37, 3.7 (2.7 to 5.1), 2.8 (2.1 to 3.9) | 9, 1.4 (0.7 to 2.6), 1.1 (0.6 to 2) | 24, 8 (5.5 to 11.7), 6.2 (4.2 to 8.9) |
| **ISL/gracile** | 62, 6.2 (4.9 to 7.9), 1.9 (1.5 to 2.4) | 10, 1.5 (0.8 to 2.8), 0.5 (0.3 to 0.9) | 48, 16.1 (12.3 to 20.6), 5 (3.8 to 6.4) |
| **Biventer** | 28, 2.8 (1.9 to 4), 2.5 (1.8 to 3.6) | 8, 1.2 (0.6 to 2.4), 1.1 (0.6 to 2.2) | 17, 5.7 (3.6 to 8.9), 5.1 (3.2 to 8.1) |
| **Tonsilla** | 14, 1.4 (0.8 to 2.3), 2.6 (1.5 to 4.3) | 5, 0.8 (0.3 to 1.8), 1.4 (0.6 to 3.2) | 7, 2.3 (1.1 to 4.8), 4.3 (2.1 to 8.7) |
| **Flocculus** | 8, 0.8 (0.4 to 1.6), 10.2 (5.2 to 20) | 6, 0.9 (0.4 to 2), 11.6 (5.3 to 25.1) | 1, 0.3 (0.1 to 1.9), 4.2 (0.8 to 23.8) |

### Supplemental Table 16. Cerebellar patterns of neuroepithelial tumor subentities

The involvement of the cerebellar structures by neuroepithelial tumors (NT) in total, glioblastoma (GBM), WHO grade III (gIIIG) and II (gIIG) glioma, developmental tumors (DT), pilocytic astrocytoma (PA), ependymoma (EP), or medulloblastoma (MB) is given in absolute numbers (n), relative tumor prevalence (RTP) and relative tumor density (RTD).

| | Total (n, RTP [95% CI], RTD [95% CI]) | GBM (n, RTP [95% CI], RTD [95% CI]) | gIIIG (n, RTP [95% CI], RTD [95% CI]) | gIIG (n, RTP [95% CI], RTD [95% CI]) | DT (n, RTP [95% CI], RTD [95% CI]) | EP (n, RTP [95% CI], RTD [95% CI]) | PA (n, RTP [95% CI], RTD [95% CI]) | MB (n, RTP [95% CI], RTD [95% CI]) |
|---|---|---|---|---|---|---|---|---|
| **Total** | 635, 100 (99.4 to 100), 1 (1 to 1) | 385, 100 (99 to 100), 1 (1 to 1) | 110, 100 (96.6 to 100), 1 (1 to 1) | 50, 100 (92.9 to 100), 1 (0.9 to 1) | 19, 100 (83.2 to 100), 1 (0.8 to 1) | 26, 100 (87.1 to 100), 1 (0.9 to 1) | 30, 100 (88.6 to 100), 1 (0.9 to 1) | 15, 100 (79.6 to 100), 1 (0.8 to 1) |
| **Vermis** | 25, 3.9 (2.7 to 5.7), 7.2 (4.9 to 10.5) | 3, 0.8 (0.3 to 2.3), 1.4 (0.5 to 4.1) | 3, 2.7 (0.9 to 7.7), 5 (1.7 to 14.1) | 0, 0 (0 to 7.1), 0 (0 to 13.1) | 1, 5.3 (0.9 to 24.6), 9.6 (1.7 to 45.1) | 1, 3.8 (0.7 to 18.9), 7 (1.2 to 34.6) | 6, 20 (9.5 to 37.3), 36.6 (17.4 to 68.3) | 11, 73.3 (48 to 89.1), 134.2 (87.9 to 163) |
| **Hemisphere** | 27, 4.3 (2.9 to 6.1), 0.4 (0.3 to 0.6) | 3, 0.8 (0.3 to 2.3), 0.1 (0 to 0.2) | 4, 3.6 (1.4 to 9), 0.4 (0.1 to 0.9) | 0, 0 (0 to 7.1), 0 (0 to 0.7) | 1, 5.3 (0.9 to 24.6), 0.5 (0.1 to 2.4) | 3, 11.5 (4 to 29), 1.1 (0.4 to 2.8) | 13, 43.3 (27.4 to 60.8), 4.3 (2.7 to 6) | 3, 20 (7 to 45.2), 2 (0.7 to 4.4) |
| **Anterior lobe** | 15, 2.4 (1.4 to 3.9), 0.8 (0.5 to 1.3) | 3, 0.8 (0.3 to 2.3), 0.3 (0.1 to 0.8) | 4, 3.6 (1.4 to 9), 1.2 (0.5 to 3) | 0, 0 (0 to 7.1), 0 (0 to 2.4) | 1, 5.3 (0.9 to 24.6), 1.8 (0.3 to 8.3) | 1, 3.8 (0.7 to 18.9), 1.3 (0.2 to 6.4) | 4, 13.3 (5.3 to 29.7), 4.5 (1.8 to 10) | 2, 13.3 (3.7 to 37.9), 4.5 (1.3 to 12.8) |
| **Medial lobe** | 14, 2.2 (1.3 to 3.7), 0.8 (0.5 to 1.4) | 3, 0.8 (0.3 to 2.3), 0.3 (0.1 to 0.9) | 4, 3.6 (1.4 to 9), 1.4 (0.5 to 3.4) | 0, 0 (0 to 7.1), 0 (0 to 2.7) | 0, 0 (0 to 16.8), 0 (0 to 6.4) | 0, 0 (0 to 12.9), 0 (0 to 4.9) | 5, 16.7 (7.3 to 33.6), 6.3 (2.8 to 12.7) | 2, 13.3 (3.7 to 37.9), 5 (1.4 to 14.3) |
| **Posterior lobe** | 23, 3.6 (2.4 to 5.4), 0.7 (0.5 to 1.1) | 3, 0.8 (0.3 to 2.3), 0.2 (0.1 to 0.5) | 2, 1.8 (0.5 to 6.4), 0.4 (0.1 to 1.3) | 0, 0 (0 to 7.1), 0 (0 to 1.4) | 0, 0 (0 to 16.8), 0 (0 to 3.3) | 1, 3.8 (0.7 to 18.9), 0.8 (0.1 to 3.8) | 11, 36.7 (21.9 to 54.5), 7.3 (4.4 to 10.9) | 6, 40 (19.8 to 64.3), 8 (3.9 to 12.8) |
| **Flocculonodular lobe** | 18, 2.8 (1.8 to 4.4), 28.8 (18.3 to 45.1) | 1, 0.3 (0 to 1.5), 2.6 (0.5 to 14.8) | 2, 1.8 (0.5 to 6.4), 18.5 (5.1 to 65) | 0, 0 (0 to 7.1), 0 (0 to 72.6) | 0, 0 (0 to 16.8), 0 (0 to 171.1) | 1, 3.8 (0.7 to 18.9), 39.1 (6.9 to 192.2) | 2, 6.7 (1.8 to 21.3), 67.8 (18.8 to 216.9) | 12, 80 (54.8 to 93), 813.7 (557.5 to 945.4) |
| **Central** | 10, 1.6 (0.9 to 2.9), 35.4 (19.3 to 64.6) | 3, 0.8 (0.3 to 2.3), 17.5 (6 to 50.9) | 3, 2.7 (0.9 to 7.7), 61.3 (20.9 to 173.3) | 0, 0 (0 to 7.1), 0 (0 to 160.3) | 0, 0 (0 to 16.8), 0 (0 to 377.9) | 0, 0 (0 to 12.9), 0 (0 to 289.3) | 2, 6.7 (1.8 to 21.3), 149.8 (41.5 to 479.2) | 2, 13.3 (3.7 to 37.9), 299.6 (84 to 851.3) |
| **Culmen** | 10, 1.6 (0.9 to 2.9), 7.8 (4.3 to 14.3) | 3, 0.8 (0.3 to 2.3), 3.9 (1.3 to 11.3) | 3, 2.7 (0.9 to 7.7), 13.6 (4.6 to 38.4) | 0, 0 (0 to 7.1), 0 (0 to 35.6) | 1, 5.3 (0.9 to 24.6), 26.2 (4.7 to 122.8) | 0, 0 (0 to 12.9), 0 (0 to 64.2) | 2, 6.7 (1.8 to 21.3), 33.2 (9.2 to 106.3) | 1, 6.7 (1.2 to 29.8), 33.2 (5.9 to 148.6) |
| **Declive** | 10, 1.6 (0.9 to 2.9), 16.7 (9.1 to 30.4) | 3, 0.8 (0.3 to 2.3), 8.2 (2.8 to 24) | 3, 2.7 (0.9 to 7.7), 28.8 (9.9 to 81.6) | 0, 0 (0 to 7.1), 0 (0 to 75.5) | 0, 0 (0 to 16.8), 0 (0 to 177.9) | 0, 0 (0 to 12.9), 0 (0 to 136.1) | 3, 10 (3.5 to 25.6), 105.8 (36.6 to 271) | 1, 6.7 (1.2 to 29.8), 70.5 (12.6 to 315.3) |
| **Folium** | 7, 1.1 (0.5 to 2.3), 27.2 (13.2 to 55.7) | 2, 0.5 (0.1 to 1.9), 12.8 (3.5 to 46.3) | 2, 1.8 (0.5 to 6.4), 44.9 (12.3 to 157.7) | 0, 0 (0 to 7.1), 0 (0 to 176.1) | 0, 0 (0 to 16.8), 0 (0 to 415.1) | 0, 0 (0 to 12.9), 0 (0 to 317.7) | 2, 6.7 (1.8 to 21.3), 164.5 (45.6 to 526.3) | 1, 6.7 (1.2 to 29.8), 164.5 (29.3 to 735.9) |

| | | | | | | | | |
|---|---|---|---|---|---|---|---|---|
| **Tuber** | 7, 1.1 (0.5 to 2.3), 31 (15.1 to 63.5) | 2, 0.5 (0.1 to 1.9), 14.6 (4 to 52.7) | 2, 1.8 (0.5 to 6.4), 51.2 (14.1 to 179.8) | 0, 0 (0 to 7.1), 0 (0 to 200.8) | 0, 0 (0 to 16.8), 0 (0 to 473.3) | 0, 0 (0 to 12.9), 0 (0 to 362.3) | 2, 6.7 (1.8 to 21.3), 187.6 (52 to 600.1) | 1, 6.7 (1.2 to 29.8), 187.6 (33.4 to 839.1) |
| **Pyramid** | 7, 1.1 (0.5 to 2.3), 28.3 (13.7 to 57.9) | 2, 0.5 (0.1 to 1.9), 13.3 (3.7 to 48.1) | 2, 1.8 (0.5 to 6.4), 46.6 (12.8 to 163.9) | 0, 0 (0 to 7.1), 0 (0 to 183) | 0, 0 (0 to 16.8), 0 (0 to 431.4) | 0, 0 (0 to 12.9), 0 (0 to 330.2) | 1, 3.3 (0.6 to 16.7), 85.5 (15.2 to 427.6) | 2, 13.3 (3.7 to 37.9), 342 (95.8 to 971.7) |
| **Uvula** | 11, 1.7 (1 to 3.1), 24 (13.4 to 42.6) | 2, 0.5 (0.1 to 1.9), 7.2 (2 to 25.9) | 2, 1.8 (0.5 to 6.4), 25.2 (6.9 to 88.4) | 0, 0 (0 to 7.1), 0 (0 to 98.8) | 0, 0 (0 to 16.8), 0 (0 to 232.9) | 0, 0 (0 to 12.9), 0 (0 to 178.2) | 1, 3.3 (0.6 to 16.7), 46.2 (8.2 to 230.8) | 6, 40 (19.8 to 64.3), 553.8 (274.5 to 889.7) |
| **Nodule** | 17, 2.7 (1.7 to 4.2), 136.4 (85.5 to 216.3) | 1, 0.3 (0 to 1.5), 13.2 (2.3 to 74.2) | 2, 1.8 (0.5 to 6.4), 92.7 (25.5 to 325.5) | 0, 0 (0 to 7.1), 0 (0 to 363.6) | 0, 0 (0 to 16.8), 0 (0 to 857) | 1, 3.8 (0.7 to 18.9), 196 (34.8 to 962.7) | 2, 6.7 (1.8 to 21.3), 339.7 (94.2 to 1086.6) | 11, 73.3 (48 to 89.1), 3736.9 (2448.5 to 4540.5) |
| **Ala lobuli centralis** | 7, 1.1 (0.5 to 2.3), 1 (0.5 to 2) | 3, 0.8 (0.3 to 2.3), 0.7 (0.2 to 2.1) | 3, 2.7 (0.9 to 7.7), 2.5 (0.8 to 7) | 0, 0 (0 to 7.1), 0 (0 to 6.5) | 0, 0 (0 to 16.8), 0 (0 to 15.2) | 0, 0 (0 to 12.9), 0 (0 to 11.7) | 1, 3.3 (0.6 to 16.7), 3 (0.5 to 15.1) | 0, 0 (0 to 20.4), 0 (0 to 18.5) |
| **AQL** | 9, 1.4 (0.7 to 2.7), 0.9 (0.5 to 1.6) | 3, 0.8 (0.3 to 2.3), 0.5 (0.2 to 1.4) | 4, 3.6 (1.4 to 9), 2.2 (0.9 to 5.5) | 0, 0 (0 to 7.1), 0 (0 to 4.4) | 1, 5.3 (0.9 to 24.6), 3.2 (0.6 to 15.2) | 1, 3.8 (0.7 to 18.9), 2.4 (0.4 to 11.6) | 0, 0 (0 to 11.4), 0 (0 to 7) | 0, 0 (0 to 20.4), 0 (0 to 12.6) |
| **PQL** | 7, 1.1 (0.5 to 2.3), 0.9 (0.4 to 1.9) | 3, 0.8 (0.3 to 2.3), 0.6 (0.2 to 1.9) | 4, 3.6 (1.4 to 9), 3 (1.2 to 7.5) | 0, 0 (0 to 7.1), 0 (0 to 5.9) | 0, 0 (0 to 16.8), 0 (0 to 14) | 0, 0 (0 to 12.9), 0 (0 to 10.7) | 0, 0 (0 to 11.4), 0 (0 to 9.4) | 0, 0 (0 to 20.4), 0 (0 to 17) |
| **SSL** | 9, 1.4 (0.7 to 2.7), 1.1 (0.6 to 2) | 3, 0.8 (0.3 to 2.3), 0.6 (0.2 to 1.7) | 2, 1.8 (0.5 to 6.4), 1.4 (0.4 to 4.9) | 0, 0 (0 to 7.1), 0 (0 to 5.5) | 0, 0 (0 to 16.8), 0 (0 to 12.9) | 0, 0 (0 to 12.9), 0 (0 to 9.9) | 3, 10 (3.5 to 25.6), 7.7 (2.7 to 19.6) | 1, 6.7 (1.2 to 29.8), 5.1 (0.9 to 22.9) |
| **ISL/gracile** | 10, 1.6 (0.9 to 2.9), 0.5 (0.3 to 0.9) | 3, 0.8 (0.3 to 2.3), 0.2 (0.1 to 0.7) | 2, 1.8 (0.5 to 6.4), 0.6 (0.2 to 2) | 0, 0 (0 to 7.1), 0 (0 to 2.2) | 0, 0 (0 to 16.8), 0 (0 to 5.2) | 1, 3.8 (0.7 to 18.9), 1.2 (0.2 to 5.9) | 4, 13.3 (5.3 to 29.7), 4.1 (1.6 to 9.2) | 0, 0 (0 to 20.4), 0 (0 to 6.3) |
| **Biventer** | 8, 1.3 (0.6 to 2.5), 1.1 (0.6 to 2.2) | 2, 0.5 (0.1 to 1.9), 0.5 (0.1 to 1.7) | 2, 1.8 (0.5 to 6.4), 1.6 (0.5 to 5.8) | 0, 0 (0 to 7.1), 0 (0 to 6.4) | 0, 0 (0 to 16.8), 0 (0 to 15.2) | 0, 0 (0 to 12.9), 0 (0 to 11.6) | 4, 13.3 (5.3 to 29.7), 12 (4.8 to 26.8) | 0, 0 (0 to 20.4), 0 (0 to 18.4) |
| **Tonsilla** | 5, 0.8 (0.3 to 1.8), 1.4 (0.6 to 3.3) | 2, 0.5 (0.1 to 1.9), 1 (0.3 to 3.4) | 2, 1.8 (0.5 to 6.4), 3.3 (0.9 to 11.7) | 0, 0 (0 to 7.1), 0 (0 to 13.1) | 0, 0 (0 to 16.8), 0 (0 to 30.8) | 0, 0 (0 to 12.9), 0 (0 to 23.6) | 1, 3.3 (0.6 to 16.7), 6.1 (1.1 to 30.5) | 0, 0 (0 to 20.4), 0 (0 to 37.3) |
| **Flocculus** | 6, 0.9 (0.4 to 2), 12 (5.5 to 26) | 1, 0.3 (0 to 1.5), 3.3 (0.6 to 18.5) | 2, 1.8 (0.5 to 6.4), 23.1 (6.4 to 81.2) | 0, 0 (0 to 7.1), 0 (0 to 90.7) | 0, 0 (0 to 16.8), 0 (0 to 213.7) | 1, 3.8 (0.7 to 18.9), 48.9 (8.7 to 240.1) | 0, 0 (0 to 11.4), 0 (0 to 144.2) | 2, 13.3 (3.7 to 37.9), 169.4 (47.5 to 481.4) |

### Supplemental Table 17. Cerebellar patterns of metastases subentities

The involvement of the cerebellar structures by metastases in total and by organ of origin is given in absolute numbers (n), relative tumor prevalence (RTP) and relative tumor density (RTD). *GIT = gastrointestinal tract; UGT = urogenital tract.*

| | Total (n, RTP [95% CI], RTD [95% CI]) | Lung (n, RTP [95% CI], RTD [95% CI]) | Skin (n, RTP [95% CI], RTD [95% CI]) | GIT (n, RTP [95% CI], RTD [95% CI]) | Breast (n, RTP [95% CI], RTD [95% CI]) | UGT (n, RTP [95% CI], RTD [95% CI]) | Miscellaneous (n, RTP [95% CI], RTD [95% CI]) |
|---|---|---|---|---|---|---|---|
| **Total** | 299, 100 (98.7 to 100), 1 (1 to 1) | 143, 100 (97.4 to 100), 1 (1 to 1) | 49, 100 (92.7 to 100), 1 (0.9 to 1) | 37, 100 (90.6 to 100), 1 (0.9 to 1) | 28, 100 (87.9 to 100), 1 (0.9 to 1) | 24, 100 (86.2 to 100), 1 (0.9 to 1) | 18, 100 (82.4 to 100), 1 (0.8 to 1) |
| **Vermis** | 5, 1.7 (0.7 to 3.9), 3.1 (1.3 to 7.1) | 2, 1.4 (0.4 to 5), 2.6 (0.7 to 9.1) | 0, 0 (0 to 7.3), 0 (0 to 13.3) | 0, 0 (0 to 9.4), 0 (0 to 17.2) | 0, 0 (0 to 12.1), 0 (0 to 22.1) | 0, 0 (0 to 13.8), 0 (0 to 25.2) | 3, 16.7 (5.8 to 39.2), 30.5 (10.7 to 71.8) |
| **Hemisphere** | 89, 29.8 (24.9 to 35.2), 2.9 (2.4 to 3.5) | 50, 35 (27.6 to 43.1), 3.4 (2.7 to 4.2) | 8, 16.3 (8.5 to 29), 1.6 (0.8 to 2.9) | 12, 32.4 (19.6 to 48.5), 3.2 (1.9 to 4.8) | 11, 39.3 (23.6 to 57.6), 3.9 (2.3 to 5.7) | 2, 8.3 (2.3 to 25.8), 0.8 (0.2 to 2.5) | 6, 33.3 (16.3 to 56.3), 3.3 (1.6 to 5.5) |
| **Anterior lobe** | 15, 5 (3.1 to 8.1), 1.7 (1 to 2.7) | 5, 3.5 (1.5 to 7.9), 1.2 (0.5 to 2.7) | 3, 6.1 (2.1 to 16.5), 2.1 (0.7 to 5.6) | 1, 2.7 (0.5 to 13.8), 0.9 (0.2 to 4.7) | 0, 0 (0 to 12.1), 0 (0 to 4.1) | 2, 8.3 (2.3 to 25.8), 2.8 (0.8 to 8.7) | 4, 22.2 (9 to 45.2), 7.5 (3 to 15.2) |
| **Medial lobe** | 23, 7.7 (5.2 to 11.3), 2.9 (2 to 4.3) | 14, 9.8 (5.9 to 15.8), 3.7 (2.2 to 6) | 2, 4.1 (1.1 to 13.7), 1.5 (0.4 to 5.2) | 3, 8.1 (2.8 to 21.3), 3.1 (1.1 to 8.1) | 2, 7.1 (2 to 22.6), 2.7 (0.7 to 8.6) | 0, 0 (0 to 13.8), 0 (0 to 5.2) | 2, 11.1 (3.1 to 32.8), 4.2 (1.2 to 12.4) |
| **Posterior lobe** | 64, 21.4 (17.1 to 26.4), 4.3 (3.4 to 5.3) | 36, 25.2 (18.8 to 32.9), 5 (3.7 to 6.5) | 4, 8.2 (3.2 to 19.2), 1.6 (0.6 to 3.8) | 10, 27 (15.4 to 43), 5.4 (3.1 to 8.6) | 10, 35.7 (20.7 to 54.2), 7.1 (4.1 to 10.8) | 0, 0 (0 to 13.8), 0 (0 to 2.7) | 4, 22.2 (9 to 45.2), 4.4 (1.8 to 9) |
| **Flocculonodular lobe** | 1, 0.3 (0.1 to 1.9), 3.4 (0.6 to 19) | 0, 0 (0 to 2.6), 0 (0 to 26.6) | 0, 0 (0 to 7.3), 0 (0 to 73.9) | 0, 0 (0 to 9.4), 0 (0 to 95.7) | 0, 0 (0 to 12.1), 0 (0 to 122.7) | 0, 0 (0 to 13.8), 0 (0 to 140.3) | 1, 5.6 (1 to 25.8), 56.5 (10 to 262) |
| **Central** | 2, 0.7 (0.2 to 2.4), 15 (4.1 to 54.1) | 0, 0 (0 to 2.6), 0 (0 to 58.8) | 0, 0 (0 to 7.3), 0 (0 to 163.4) | 0, 0 (0 to 9.4), 0 (0 to 211.4) | 0, 0 (0 to 12.1), 0 (0 to 271.1) | 0, 0 (0 to 13.8), 0 (0 to 310.1) | 2, 11.1 (3.1 to 32.8), 249.7 (69.7 to 737.1) |
| **Culmen** | 3, 1 (0.3 to 2.9), 5 (1.7 to 14.5) | 1, 0.7 (0.1 to 3.9), 3.5 (0.6 to 19.2) | 0, 0 (0 to 7.3), 0 (0 to 36.2) | 0, 0 (0 to 9.4), 0 (0 to 46.9) | 0, 0 (0 to 12.1), 0 (0 to 60.1) | 0, 0 (0 to 13.8), 0 (0 to 68.8) | 2, 11.1 (3.1 to 32.8), 55.4 (15.5 to 163.5) |
| **Declive** | 1, 0.3 (0.1 to 1.9), 3.5 (0.6 to 19.8) | 0, 0 (0 to 2.6), 0 (0 to 27.7) | 0, 0 (0 to 7.3), 0 (0 to 76.9) | 0, 0 (0 to 9.4), 0 (0 to 99.5) | 0, 0 (0 to 12.1), 0 (0 to 127.6) | 0, 0 (0 to 13.8), 0 (0 to 145.9) | 1, 5.6 (1 to 25.8), 58.8 (10.4 to 272.4) |
| **Folium** | 2, 0.7 (0.2 to 2.4), 16.5 (4.5 to 59.4) | 1, 0.7 (0.1 to 3.9), 17.3 (3 to 95.1) | 0, 0 (0 to 7.3), 0 (0 to 179.4) | 0, 0 (0 to 9.4), 0 (0 to 232.1) | 0, 0 (0 to 12.1), 0 (0 to 297.8) | 0, 0 (0 to 13.8), 0 (0 to 340.5) | 1, 5.6 (1 to 25.8), 137.1 (24.4 to 635.7) |
| **Tuber** | 1, 0.3 (0.1 to 1.9), 9.4 (1.7 to 52.6) | 0, 0 (0 to 2.6), 0 (0 to 73.6) | 0, 0 (0 to 7.3), 0 (0 to 204.6) | 0, 0 (0 to 9.4), 0 (0 to 264.7) | 0, 0 (0 to 12.1), 0 (0 to 339.5) | 0, 0 (0 to 13.8), 0 (0 to 388.3) | 1, 5.6 (1 to 25.8), 156.3 (27.8 to 724.8) |
| **Pyramid** | 1, 0.3 (0.1 to 1.9), 8.6 (1.5 to 48) | 0, 0 (0 to 2.6), 0 (0 to 67.1) | 0, 0 (0 to 7.3), 0 (0 to 186.5) | 0, 0 (0 to 9.4), 0 (0 to 241.3) | 0, 0 (0 to 12.1), 0 (0 to 309.5) | 0, 0 (0 to 13.8), 0 (0 to 353.9) | 1, 5.6 (1 to 25.8), 142.5 (25.3 to 660.7) |
| **Uvula** | 1, 0.3 (0.1 to 1.9), 4.6 (0.8 to 25.9) | 0, 0 (0 to 2.6), 0 (0 to 36.2) | 0, 0 (0 to 7.3), 0 (0 to 100.7) | 0, 0 (0 to 9.4), 0 (0 to 130.2) | 0, 0 (0 to 12.1), 0 (0 to 167) | 0, 0 (0 to 13.8), 0 (0 to 191) | 1, 5.6 (1 to 25.8), 76.9 (13.7 to 356.6) |

| | | | | | | | |
|---|---|---|---|---|---|---|---|
| **Nodule** | 1, 0.3 (0.1 to 1.9), 17 (3 to 95.3) | 0, 0 (0 to 2.6), 0 (0 to 133.3) | 0, 0 (0 to 7.3), 0 (0 to 370.5) | 0, 0 (0 to 9.4), 0 (0 to 479.3) | 0, 0 (0 to 12.1), 0 (0 to 614.8) | 0, 0 (0 to 13.8), 0 (0 to 703.1) | 1, 5.6 (1 to 25.8), 283.1 (50.3 to 1312.6) |
| **Ala lobuli centralis** | 3, 1 (0.3 to 2.9), 0.9 (0.3 to 2.6) | 0, 0 (0 to 2.6), 0 (0 to 2.4) | 0, 0 (0 to 7.3), 0 (0 to 6.6) | 1, 2.7 (0.5 to 13.8), 2.4 (0.4 to 12.5) | 0, 0 (0 to 12.1), 0 (0 to 10.9) | 1, 4.2 (0.7 to 20.2), 3.8 (0.7 to 18.3) | 1, 5.6 (1 to 25.8), 5 (0.9 to 23.3) |
| **AQL** | 8, 2.7 (1.4 to 5.2), 1.6 (0.8 to 3.2) | 4, 2.8 (1.1 to 7), 1.7 (0.7 to 4.3) | 1, 2 (0.4 to 10.7), 1.3 (0.2 to 6.6) | 0, 0 (0 to 9.4), 0 (0 to 5.8) | 0, 0 (0 to 12.1), 0 (0 to 7.4) | 0, 0 (0 to 13.8), 0 (0 to 8.5) | 3, 16.7 (5.8 to 39.2), 10.3 (3.6 to 24.2) |
| **PQL** | 4, 1.3 (0.5 to 3.4), 1.1 (0.4 to 2.8) | 2, 1.4 (0.4 to 5), 1.2 (0.3 to 4.1) | 0, 0 (0 to 7.3), 0 (0 to 6) | 0, 0 (0 to 9.4), 0 (0 to 7.8) | 0, 0 (0 to 12.1), 0 (0 to 10) | 1, 4.2 (0.7 to 20.2), 3.5 (0.6 to 16.8) | 1, 5.6 (1 to 25.8), 4.6 (0.8 to 21.4) |
| **SSL** | 24, 8 (5.5 to 11.7), 6.2 (4.2 to 8.9) | 13, 9.1 (5.4 to 14.9), 7 (4.1 to 11.4) | 4, 8.2 (3.2 to 19.2), 6.3 (2.5 to 14.7) | 2, 5.4 (1.5 to 17.7), 4.1 (1.1 to 13.6) | 3, 10.7 (3.7 to 27.2), 8.2 (2.8 to 20.8) | 0, 0 (0 to 13.8), 0 (0 to 10.6) | 2, 11.1 (3.1 to 32.8), 8.5 (2.4 to 25.1) |
| **ISL/gracile** | 48, 16.1 (12.3 to 20.6), 5 (3.8 to 6.4) | 26, 18.2 (12.7 to 25.3), 5.6 (3.9 to 7.9) | 4, 8.2 (3.2 to 19.2), 2.5 (1 to 6) | 7, 18.9 (9.5 to 34.2), 5.9 (2.9 to 10.6) | 9, 32.1 (17.9 to 50.7), 10 (5.6 to 15.7) | 0, 0 (0 to 13.8), 0 (0 to 4.3) | 2, 11.1 (3.1 to 32.8), 3.5 (1 to 10.2) |
| **Biventer** | 17, 5.7 (3.6 to 8.9), 5.1 (3.2 to 8.1) | 8, 5.6 (2.9 to 10.7), 5.1 (2.6 to 9.6) | 0, 0 (0 to 7.3), 0 (0 to 6.6) | 5, 13.5 (5.9 to 28), 12.2 (5.3 to 25.3) | 0, 0 (0 to 12.1), 0 (0 to 10.9) | 0, 0 (0 to 13.8), 0 (0 to 12.5) | 4, 22.2 (9 to 45.2), 20.1 (8.1 to 40.8) |
| **Tonsilla** | 7, 2.3 (1.1 to 4.8), 4.3 (2.1 to 8.7) | 3, 2.1 (0.7 to 6), 3.8 (1.3 to 11) | 1, 2 (0.4 to 10.7), 3.7 (0.7 to 19.6) | 0, 0 (0 to 9.4), 0 (0 to 17.2) | 1, 3.6 (0.6 to 17.7), 6.5 (1.2 to 32.4) | 0, 0 (0 to 13.8), 0 (0 to 25.3) | 2, 11.1 (3.1 to 32.8), 20.3 (5.7 to 60) |
| **Flocculus** | 1, 0.3 (0.1 to 1.9), 4.2 (0.8 to 23.8) | 0, 0 (0 to 2.6), 0 (0 to 33.2) | 0, 0 (0 to 7.3), 0 (0 to 92.4) | 0, 0 (0 to 9.4), 0 (0 to 119.5) | 0, 0 (0 to 12.1), 0 (0 to 153.3) | 0, 0 (0 to 13.8), 0 (0 to 175.3) | 1, 5.6 (1 to 25.8), 70.6 (12.5 to 327.3) |

**Supplemental Table 18. Ventricular segmental patterns of neuroepithelial tumors, primary central nervous system lymphoma and brain metastases**

The involvement of the ventricular segments by brain tumors in total, neuroepithelial tumors (NT), primary central nervous system lymphoma (PCNSL) or metastases is given in absolute numbers (n), relative tumor prevalence (RTP) and relative tumor density (RTD).

| | Total (n, RTP [95% CI], RTD [95% CI]) | NT (n, RTP [95% CI], RTD [95% CI]) | PCNSL (n, RTP [95% CI], RTD [95% CI]) | Metastases (n, RTP [95% CI], RTD [95% CI]) |
|---|---|---|---|---|
| **Total** | 1000, 100 (99.6 to 100), 1 (1 to 1) | 657, 100 (99.4 to 100), 1 (1 to 1) | 44, 100 (92 to 100), 1 (0.9 to 1) | 299, 100 (98.7 to 100), 1 (1 to 1) |
| **Lateral ventricle total** | 556, 55.6 (52.5 to 58.7), 0.7 (0.6 to 0.7) | 506, 77 (73.6 to 80.1), 0.9 (0.9 to 1) | 25, 56.8 (42.2 to 70.3), 0.7 (0.5 to 0.8) | 25, 8.4 (5.7 to 12.1), 0.1 (0.1 to 0.1) |
| **Lateral ventricle frontal horn** | 198, 19.8 (17.4 to 22.4), 0.7 (0.6 to 0.8) | 176, 26.8 (23.5 to 30.3), 1 (0.8 to 1.1) | 16, 36.4 (23.8 to 51.1), 1.3 (0.8 to 1.8) | 6, 2 (0.9 to 4.3), 0.1 (0 to 0.2) |
| **Lateral ventricle body** | 160, 16 (13.9 to 18.4), 0.8 (0.7 to 0.9) | 134, 20.4 (17.5 to 23.6), 1 (0.8 to 1.1) | 21, 47.7 (33.8 to 62.1), 2.3 (1.6 to 2.9) | 5, 1.7 (0.7 to 3.9), 0.1 (0 to 0.2) |
| **Lateral ventricle atrium** | 275, 27.5 (24.8 to 30.3), 1.1 (1 to 1.3) | 245, 37.3 (33.7 to 41.1), 1.5 (1.4 to 1.7) | 20, 45.5 (31.7 to 59.9), 1.9 (1.3 to 2.5) | 10, 3.3 (1.8 to 6), 0.1 (0.1 to 0.2) |
| **Lateral ventricle occipital horn** | 102, 10.2 (8.5 to 12.2), 2.1 (1.7 to 2.5) | 85, 12.9 (10.6 to 15.7), 2.6 (2.1 to 3.2) | 10, 22.7 (12.8 to 37), 4.6 (2.6 to 7.5) | 7, 2.3 (1.1 to 4.8), 0.5 (0.2 to 1) |
| **Lateral ventricle temporal horn** | 184, 18.4 (16.1 to 20.9), 3.8 (3.3 to 4.3) | 167, 25.4 (22.2 to 28.9), 5.3 (4.6 to 6) | 11, 25 (14.6 to 39.4), 5.2 (3 to 8.2) | 6, 2 (0.9 to 4.3), 0.4 (0.2 to 0.9) |
| **Third ventricle** | 44, 4.4 (3.3 to 5.9), 0.7 (0.5 to 1) | 34, 5.2 (3.7 to 7.1), 0.9 (0.6 to 1.2) | 8, 18.2 (9.5 to 32), 3 (1.6 to 5.3) | 2, 0.7 (0.2 to 2.4), 0.1 (0 to 0.4) |
| **Fourth ventricle total** | 83, 8.3 (6.7 to 10.2), 0.8 (0.6 to 0.9) | 74, 11.3 (9.1 to 13.9), 1 (0.8 to 1.3) | 5, 11.4 (5 to 24), 1.1 (0.5 to 2.2) | 4, 1.3 (0.5 to 3.4), 0.1 (0 to 0.3) |
| **Fourth ventricle apex** | 15, 1.5 (0.9 to 2.5), 1.4 (0.8 to 2.2) | 12, 1.8 (1 to 3.2), 1.6 (0.9 to 2.9) | 2, 4.5 (1.3 to 15.1), 4.1 (1.1 to 13.6) | 1, 0.3 (0.1 to 1.9), 0.3 (0.1 to 1.7) |
| **Fourth ventricle lateral recess** | 70, 7 (5.6 to 8.8), 5.4 (4.3 to 6.7) | 65, 9.9 (7.8 to 12.4), 7.6 (6 to 9.5) | 3, 6.8 (2.3 to 18.2), 5.2 (1.8 to 14) | 2, 0.7 (0.2 to 2.4), 0.5 (0.1 to 1.8) |
| **Fourth ventricle obex** | 20, 2 (1.3 to 3.1), 1.8 (1.1 to 2.7) | 17, 2.6 (1.6 to 4.1), 2.3 (1.4 to 3.6) | 2, 4.5 (1.3 to 15.1), 4 (1.1 to 13.3) | 1, 0.3 (0.1 to 1.9), 0.3 (0.1 to 1.6) |
| **Fourth ventricle fastigium** | 26, 2.6 (1.8 to 3.8), 2 (1.4 to 2.9) | 19, 2.9 (1.9 to 4.5), 2.2 (1.4 to 3.4) | 5, 11.4 (5 to 24), 8.7 (3.8 to 18.3) | 2, 0.7 (0.2 to 2.4), 0.5 (0.1 to 1.8) |

**Supplemental Table 19. Ventricular segmental patterns of neuroepithelial tumor subentities**

The involvement of the ventricular segments by neuroepithelial tumors (NT) in total, glioblastoma (GBM), WHO grade III (gIIIG) and II (gIIG) glioma, developmental tumors (DT), pilocytic astrocytoma (PA), ependymoma (EP), or medulloblastoma (MB) is given in absolute numbers (n), relative tumor prevalence (RTP) and relative tumor density (RTD).

| | Total (n, RTP [95% CI], RTD [95% CI]) | GBM (n, RTP [95% CI], RTD [95% CI]) | gIIIG (n, RTP [95% CI], RTD [95% CI]) | gIIG (n, RTP [95% CI], RTD [95% CI]) | DT (n, RTP [95% CI], RTD [95% CI]) | EP (n, RTP [95% CI], RTD [95% CI]) | MB (n, RTP [95% CI], RTD [95% CI]) | PA (n, RTP [95% CI], RTD [95% CI]) |
|---|---|---|---|---|---|---|---|---|
| **Total** | 635, 100 (99.4 to 100), 1 (1 to 1) | 385, 100 (99 to 100), 1 (1 to 1) | 110, 100 (96.6 to 100), 1 (1 to 1) | 50, 100 (92.9 to 100), 1 (0.9 to 1) | 19, 100 (83.2 to 100), 1 (0.8 to 1) | 26, 100 (87.1 to 100), 1 (0.9 to 1) | 15, 100 (79.6 to 100), 1 (0.8 to 1) | 30, 100 (88.6 to 100), 1 (0.9 to 1) |
| **Lateral ventricle total** | 493, 77.6 (74.2 to 80.7), 0.9 (0.9 to 1) | 357, 92.7 (89.7 to 94.9), 1.1 (1.1 to 1.1) | 87, 79.1 (70.6 to 85.6), 1 (0.8 to 1) | 34, 68 (54.2 to 79.2), 0.8 (0.7 to 1) | 8, 42.1 (23.1 to 63.7), 0.5 (0.3 to 0.8) | 5, 19.2 (8.5 to 37.9), 0.2 (0.1 to 0.5) | 0, 0 (0 to 20.4), 0 (0 to 0.2) | 2, 6.7 (1.8 to 21.3), 0.1 (0 to 0.3) |
| **Lateral ventricle frontal horn** | 168, 26.5 (23.2 to 30), 0.9 (0.8 to 1.1) | 101, 26.2 (22.1 to 30.8), 0.9 (0.8 to 1.1) | 43, 39.1 (30.5 to 48.4), 1.4 (1.1 to 1.7) | 21, 42 (29.4 to 55.8), 1.5 (1 to 2) | 1, 5.3 (0.9 to 24.6), 0.2 (0 to 0.9) | 1, 3.8 (0.7 to 18.9), 0.1 (0 to 0.7) | 0, 0 (0 to 20.4), 0 (0 to 0.7) | 1, 3.3 (0.6 to 16.7), 0.1 (0 to 0.6) |
| **Lateral ventricle body** | 130, 20.5 (17.5 to 23.8), 1 (0.8 to 1.1) | 90, 23.4 (19.4 to 27.9), 1.1 (0.9 to 1.3) | 27, 24.5 (17.5 to 33.4), 1.2 (0.8 to 1.6) | 13, 26 (15.9 to 39.6), 1.2 (0.8 to 1.9) | 0, 0 (0 to 16.8), 0 (0 to 0.8) | 0, 0 (0 to 12.9), 0 (0 to 0.6) | 0, 0 (0 to 20.4), 0 (0 to 1) | 0, 0 (0 to 11.4), 0 (0 to 0.5) |
| **Lateral ventricle atrium** | 241, 38 (34.3 to 41.8), 1.6 (1.4 to 1.7) | 197, 51.2 (46.2 to 56.1), 2.1 (1.9 to 2.3) | 25, 22.7 (15.9 to 31.4), 0.9 (0.7 to 1.3) | 10, 20 (11.2 to 33), 0.8 (0.5 to 1.4) | 4, 21.1 (8.5 to 43.3), 0.9 (0.4 to 1.8) | 4, 15.4 (6.2 to 33.5), 0.6 (0.3 to 1.4) | 0, 0 (0 to 20.4), 0 (0 to 0.8) | 1, 3.3 (0.6 to 16.7), 0.1 (0 to 0.7) |
| **Lateral ventricle occipital horn** | 83, 13.1 (10.7 to 15.9), 2.6 (2.2 to 3.2) | 73, 19 (15.4 to 23.2), 3.8 (3.1 to 4.7) | 7, 6.4 (3.1 to 12.6), 1.3 (0.6 to 2.5) | 3, 6 (2.1 to 16.2), 1.2 (0.4 to 3.3) | 0, 0 (0 to 16.8), 0 (0 to 3.4) | 0, 0 (0 to 12.9), 0 (0 to 2.6) | 0, 0 (0 to 20.4), 0 (0 to 4.1) | 0, 0 (0 to 11.4), 0 (0 to 2.3) |
| **Lateral ventricle temporal horn** | 166, 26.1 (22.9 to 29.7), 5.4 (4.7 to 6.1) | 121, 31.4 (27 to 36.2), 6.5 (5.6 to 7.5) | 30, 27.3 (19.8 to 36.3), 5.6 (4.1 to 7.5) | 10, 20 (11.2 to 33), 4.1 (2.3 to 6.8) | 4, 21.1 (8.5 to 43.3), 4.4 (1.8 to 9) | 1, 3.8 (0.7 to 18.9), 0.8 (0.1 to 3.9) | 0, 0 (0 to 20.4), 0 (0 to 4.2) | 0, 0 (0 to 11.4), 0 (0 to 2.4) |
| **Third ventricle** | 28, 4.4 (3.1 to 6.3), 0.7 (0.5 to 1) | 5, 1.3 (0.6 to 3), 0.2 (0.1 to 0.5) | 11, 10 (5.7 to 17), 1.7 (0.9 to 2.8) | 5, 10 (4.3 to 21.4), 1.7 (0.7 to 3.5) | 1, 5.3 (0.9 to 24.6), 0.9 (0.2 to 4.1) | 1, 3.8 (0.7 to 18.9), 0.6 (0.1 to 3.1) | 0, 0 (0 to 20.4), 0 (0 to 3.4) | 5, 16.7 (7.3 to 33.6), 2.8 (1.2 to 5.6) |
| **Fourth ventricle total** | 70, 11 (8.8 to 13.7), 1 (0.8 to 1.3) | 6, 1.6 (0.7 to 3.4), 0.1 (0.1 to 0.3) | 9, 8.2 (4.4 to 14.8), 0.8 (0.4 to 1.4) | 2, 4 (1.1 to 13.5), 0.4 (0.1 to 1.2) | 2, 10.5 (2.9 to 31.4), 1 (0.3 to 2.9) | 15, 57.7 (38.9 to 74.5), 5.4 (3.6 to 6.9) | 14, 93.3 (70.2 to 98.8), 8.7 (6.5 to 9.2) | 22, 73.3 (55.6 to 85.8), 6.8 (5.2 to 8) |
| **Fourth ventricle apex** | 11, 1.7 (1 to 3.1), 1.6 (0.9 to 2.8) | 3, 0.8 (0.3 to 2.3), 0.7 (0.2 to 2) | 6, 5.5 (2.5 to 11.4), 4.9 (2.3 to 10.3) | 1, 2 (0.4 to 10.5), 1.8 (0.3 to 9.5) | 0, 0 (0 to 16.8), 0 (0 to 15.2) | 0, 0 (0 to 12.9), 0 (0 to 11.6) | 1, 6.7 (1.2 to 29.8), 6 (1.1 to 26.9) | 0, 0 (0 to 11.4), 0 (0 to 10.2) |

| | | | | | | | | |
|---|---|---|---|---|---|---|---|---|
| **Fourth ventricle lateral recess** | 61, 9.6 (7.6 to 12.1), 7.4 (5.8 to 9.3) | 5, 1.3 (0.6 to 3), 1 (0.4 to 2.3) | 9, 8.2 (4.4 to 14.8), 6.3 (3.3 to 11.3) | 2, 4 (1.1 to 13.5), 3.1 (0.8 to 10.3) | 1, 5.3 (0.9 to 24.6), 4 (0.7 to 18.9) | 14, 53.8 (35.5 to 71.2), 41.2 (27.1 to 54.5) | 11, 73.3 (48 to 89.1), 56.1 (36.8 to 68.2) | 19, 63.3 (45.5 to 78.1), 48.5 (34.8 to 59.8) |
| **Fourth ventricle obex** | 15, 2.4 (1.4 to 3.9), 2.1 (1.3 to 3.4) | 3, 0.8 (0.3 to 2.3), 0.7 (0.2 to 2) | 6, 5.5 (2.5 to 11.4), 4.8 (2.2 to 10) | 1, 2 (0.4 to 10.5), 1.8 (0.3 to 9.2) | 1, 5.3 (0.9 to 24.6), 4.6 (0.8 to 21.6) | 1, 3.8 (0.7 to 18.9), 3.4 (0.6 to 16.6) | 2, 13.3 (3.7 to 37.9), 11.7 (3.3 to 33.3) | 1, 3.3 (0.6 to 16.7), 2.9 (0.5 to 14.6) |
| **Fourth ventricle fastigium** | 18, 2.8 (1.8 to 4.4), 2.2 (1.4 to 3.4) | 4, 1 (0.4 to 2.6), 0.8 (0.3 to 2) | 6, 5.5 (2.5 to 11.4), 4.2 (1.9 to 8.7) | 0, 0 (0 to 7.1), 0 (0 to 5.4) | 0, 0 (0 to 16.8), 0 (0 to 12.8) | 0, 0 (0 to 12.9), 0 (0 to 9.8) | 5, 33.3 (15.2 to 58.3), 25.4 (11.6 to 44.4) | 3, 10 (3.5 to 25.6), 7.6 (2.6 to 19.5) |

**Supplemental Table 20. Ventricular segmentality of neuroepithelial tumors**

The involvement of the ventricular segments by univentriculosegmental (uni) or mutiventriculosegmental (multi) neuroepithelial tumors (NT) is given in absolute numbers (n), relative tumor prevalence (RTP) and relative tumor density (RTD).

| | Total (n, RTP [95% CI], RTD [95% CI]) | uni (n, RTP [95% CI], RTD [95% CI]) | multi (n, RTP [95% CI], RTD [95% CI]) |
|---|---|---|---|
| **Total** | 588, 100 (99.4 to 100), 1 (1 to 1) | 388, 100 (99 to 100), 1 (1 to 1) | 200, 100 (98.1 to 100), 1 (1 to 1) |
| **Lateral ventricle total** | 506, 86.1 (83 to 88.6), 1 (1 to 1.1) | 308, 79.4 (75.1 to 83.1), 1 (0.9 to 1) | 198, 99 (96.4 to 99.7), 1.2 (1.2 to 1.2) |
| **Lateral ventricle frontal horn** | 176, 29.9 (26.4 to 33.8), 1.1 (0.9 to 1.2) | 101, 26 (21.9 to 30.6), 0.9 (0.8 to 1.1) | 75, 37.5 (31.1 to 44.4), 1.3 (1.1 to 1.6) |
| **Lateral ventricle body** | 134, 22.8 (19.6 to 26.3), 1.1 (0.9 to 1.2) | 46, 11.9 (9 to 15.5), 0.6 (0.4 to 0.7) | 88, 44 (37.3 to 50.9), 2.1 (1.8 to 2.4) |
| **Lateral ventricle atrium** | 245, 41.7 (37.7 to 45.7), 1.7 (1.6 to 1.9) | 89, 22.9 (19 to 27.4), 0.9 (0.8 to 1.1) | 156, 78 (71.8 to 83.2), 3.2 (3 to 3.4) |
| **Lateral ventricle occipital horn** | 85, 14.5 (11.8 to 17.5), 2.9 (2.4 to 3.5) | 14, 3.6 (2.2 to 6), 0.7 (0.4 to 1.2) | 71, 35.5 (29.2 to 42.3), 7.2 (5.9 to 8.5) |
| **Lateral ventricle temporal horn** | 167, 28.4 (24.9 to 32.2), 5.9 (5.2 to 6.7) | 59, 15.2 (12 to 19.1), 3.1 (2.5 to 4) | 108, 54 (47.1 to 60.8), 11.2 (9.7 to 12.6) |
| **Third ventricle** | 34, 5.8 (4.2 to 8), 1 (0.7 to 1.3) | 14, 3.6 (2.2 to 6), 0.6 (0.4 to 1) | 20, 10 (6.6 to 14.9), 1.7 (1.1 to 2.5) |
| **Fourth ventricle total** | 74, 12.6 (10.1 to 15.5), 1.2 (0.9 to 1.4) | 65, 16.8 (13.4 to 20.8), 1.6 (1.2 to 1.9) | 9, 4.5 (2.4 to 8.3), 0.4 (0.2 to 0.8) |
| **Fourth ventricle apex** | 12, 2 (1.2 to 3.5), 1.8 (1.1 to 3.2) | 3, 0.8 (0.3 to 2.2), 0.7 (0.2 to 2) | 9, 4.5 (2.4 to 8.3), 4.1 (2.1 to 7.5) |
| **Fourth ventricle lateral recess** | 65, 11.1 (8.8 to 13.8), 8.5 (6.7 to 10.6) | 56, 14.4 (11.3 to 18.3), 11.1 (8.6 to 14) | 9, 4.5 (2.4 to 8.3), 3.4 (1.8 to 6.4) |
| **Fourth ventricle obex** | 17, 2.9 (1.8 to 4.6), 2.5 (1.6 to 4) | 9, 2.3 (1.2 to 4.3), 2 (1.1 to 3.8) | 8, 4 (2 to 7.7), 3.5 (1.8 to 6.8) |
| **Fourth ventricle fastigium** | 19, 3.2 (2.1 to 5), 2.5 (1.6 to 3.8) | 11, 2.8 (1.6 to 5), 2.2 (1.2 to 3.8) | 8, 4 (2 to 7.7), 3 (1.6 to 5.9) |

**Supplemental Table 21. Ventriculo-parenchymal topology of neuroepithelial tumors**

The co-involvement of ventricular and parenchymal segments by neuroepithelial tumors (NT) is given in absolute numbers (n), relative tumor prevalence (RTP) and relative tumor density (RTD).

| | FH (n, RTP [95% CI], RTD [95% CI]) | Body (n, RTP [95% CI], RTD [95% CI]) | Atrium (n, RTP [95% CI], RTD [95% CI]) | OH (n, RTP [95% CI], RTD [95% CI]) | TH (n, RTP [95% CI], RTD [95% CI]) | 3rd (n, RTP [95% CI], RTD [95% CI]) | 4th (n, RTP [95% CI], RTD [95% CI]) | multi (n, RTP [95% CI], RTD [95% CI]) | none (n, RTP [95% CI], RTD [95% CI]) | Total (n, RTP [95% CI], RTD [95% CI]) |
|---|---|---|---|---|---|---|---|---|---|---|
| **Total** | 101, 100 (96.3 to 100), 1 (1 to 1) | 46, 100 (92.3 to 100), 1 (0.9 to 1) | 89, 100 (95.9 to 100), 1 (1 to 1) | 14, 100 (78.5 to 100), 1 (0.8 to 1) | 59, 100 (93.9 to 100), 1 (0.9 to 1) | 14, 100 (78.5 to 100), 1 (0.8 to 1) | 65, 100 (94.4 to 100), 1 (0.9 to 1) | 200, 100 (98.1 to 100), 1 (1 to 1) | 69, 100 (94.7 to 100), 1 (0.9 to 1) | 657, 100 (99.4 to 100), 1 (1 to 1) |
| **Frontal pole** | 9, 8.9 (4.8 to 16.1), 20.7 (11 to 37.3) | 0, 0 (0 to 7.7), 0 (0 to 17.9) | 0, 0 (0 to 4.1), 0 (0 to 9.6) | 0, 0 (0 to 21.5), 0 (0 to 50) | 0, 0 (0 to 6.1), 0 (0 to 14.2) | 0, 0 (0 to 21.5), 0 (0 to 50) | 0, 0 (0 to 5.6), 0 (0 to 13) | 2, 1 (0.3 to 3.6), 2.3 (0.6 to 8.3) | 0, 0 (0 to 5.3), 0 (0 to 12.2) | 11, 1.7 (0.9 to 3), 3.9 (2.2 to 6.9) |
| **F1** | 36, 35.6 (27 to 45.4), 5.7 (4.3 to 7.2) | 15, 32.6 (20.9 to 47), 5.2 (3.3 to 7.5) | 0, 0 (0 to 4.1), 0 (0 to 0.7) | 0, 0 (0 to 21.5), 0 (0 to 3.4) | 0, 0 (0 to 6.1), 0 (0 to 1) | 0, 0 (0 to 21.5), 0 (0 to 3.4) | 0, 0 (0 to 5.6), 0 (0 to 0.9) | 23, 11.5 (7.8 to 16.7), 1.8 (1.2 to 2.7) | 7, 10.1 (5 to 19.5), 1.6 (0.8 to 3.1) | 81, 12.3 (10 to 15.1), 2 (1.6 to 2.4) |
| **F2** | 28, 27.7 (19.9 to 37.1), 5.2 (3.7 to 7) | 4, 8.7 (3.4 to 20.3), 1.6 (0.6 to 3.8) | 1, 1.1 (0.2 to 6.1), 0.2 (0 to 1.1) | 0, 0 (0 to 21.5), 0 (0 to 4) | 0, 0 (0 to 6.1), 0 (0 to 1.1) | 0, 0 (0 to 21.5), 0 (0 to 4) | 0, 0 (0 to 5.6), 0 (0 to 1) | 21, 10.5 (7 to 15.5), 2 (1.3 to 2.9) | 3, 4.3 (1.5 to 12), 0.8 (0.3 to 2.3) | 57, 8.7 (6.8 to 11.1), 1.6 (1.3 to 2.1) |
| **F3 orbital** | 12, 11.9 (6.9 to 19.6), 33.2 (19.4 to 54.8) | 0, 0 (0 to 7.7), 0 (0 to 21.5) | 0, 0 (0 to 4.1), 0 (0 to 11.6) | 0, 0 (0 to 21.5), 0 (0 to 60.1) | 0, 0 (0 to 6.1), 0 (0 to 17.1) | 0, 0 (0 to 21.5), 0 (0 to 60.1) | 0, 0 (0 to 5.6), 0 (0 to 15.6) | 11, 5.5 (3.1 to 9.6), 15.4 (8.7 to 26.8) | 0, 0 (0 to 5.3), 0 (0 to 14.7) | 23, 3.5 (2.3 to 5.2), 9.8 (6.5 to 14.5) |
| **F3 triangular** | 13, 12.9 (7.7 to 20.8), 12 (7.2 to 19.4) | 0, 0 (0 to 7.7), 0 (0 to 7.2) | 0, 0 (0 to 4.1), 0 (0 to 3.9) | 0, 0 (0 to 21.5), 0 (0 to 20.1) | 0, 0 (0 to 6.1), 0 (0 to 5.7) | 0, 0 (0 to 21.5), 0 (0 to 20.1) | 0, 0 (0 to 5.6), 0 (0 to 5.2) | 11, 5.5 (3.1 to 9.6), 5.1 (2.9 to 8.9) | 1, 1.4 (0.3 to 7.8), 1.4 (0.2 to 7.2) | 25, 3.8 (2.6 to 5.6), 3.6 (2.4 to 5.2) |
| **F3 opercular** | 11, 10.9 (6.2 to 18.5), 8.5 (4.8 to 14.4) | 4, 8.7 (3.4 to 20.3), 6.8 (2.7 to 15.9) | 0, 0 (0 to 4.1), 0 (0 to 3.2) | 0, 0 (0 to 21.5), 0 (0 to 16.8) | 0, 0 (0 to 6.1), 0 (0 to 4.8) | 0, 0 (0 to 21.5), 0 (0 to 16.8) | 0, 0 (0 to 5.6), 0 (0 to 4.4) | 12, 6 (3.5 to 10.2), 4.7 (2.7 to 8) | 4, 5.8 (2.3 to 14), 4.5 (1.8 to 10.9) | 31, 4.7 (3.3 to 6.6), 3.7 (2.6 to 5.2) |
| **Anterior orbital** | 9, 8.9 (4.8 to 16.1), 29.1 (15.6 to 52.6) | 0, 0 (0 to 7.7), 0 (0 to 25.2) | 0, 0 (0 to 4.1), 0 (0 to 13.5) | 0, 0 (0 to 21.5), 0 (0 to 70.4) | 0, 0 (0 to 6.1), 0 (0 to 20) | 0, 0 (0 to 21.5), 0 (0 to 70.4) | 0, 0 (0 to 5.6), 0 (0 to 18.2) | 0, 0 (0 to 1.9), 0 (0 to 6.2) | 0, 0 (0 to 5.3), 0 (0 to 17.2) | 9, 1.4 (0.7 to 2.6), 4.5 (2.4 to 8.4) |
| **Medial orbital** | 11, 10.9 (6.2 to 18.5), 15.8 (9 to 26.7) | 0, 0 (0 to 7.7), 0 (0 to 11.2) | 0, 0 (0 to 4.1), 0 (0 to 6) | 0, 0 (0 to 21.5), 0 (0 to 31.2) | 0, 0 (0 to 6.1), 0 (0 to 8.8) | 0, 0 (0 to 21.5), 0 (0 to 31.2) | 0, 0 (0 to 5.6), 0 (0 to 8.1) | 3, 1.5 (0.5 to 4.3), 2.2 (0.7 to 6.2) | 0, 0 (0 to 5.3), 0 (0 to 7.6) | 14, 2.1 (1.3 to 3.5), 3.1 (1.8 to 5.1) |

| | | | | | | | | | | |
|---|---|---|---|---|---|---|---|---|---|---|
| **Lateral orbital** | 11, 10.9 (6.2 to 18.5), 19.7 (11.2 to 33.3) | 0, 0 (0 to 7.7), 0 (0 to 13.9) | 0, 0 (0 to 4.1), 0 (0 to 7.5) | 0, 0 (0 to 21.5), 0 (0 to 38.9) | 0, 0 (0 to 6.1), 0 (0 to 11) | 0, 0 (0 to 21.5), 0 (0 to 38.9) | 0, 0 (0 to 5.6), 0 (0 to 10.1) | 0, 0 (0 to 1.9), 0 (0 to 3.4) | 0, 0 (0 to 5.3), 0 (0 to 9.5) | 11, 1.7 (0.9 to 3), 3 (1.7 to 5.4) |
| **Posterior orbital** | 20, 19.8 (13.2 to 28.6), 29.2 (19.5 to 42.2) | 0, 0 (0 to 7.7), 0 (0 to 11.4) | 0, 0 (0 to 4.1), 0 (0 to 6.1) | 0, 0 (0 to 21.5), 0 (0 to 31.8) | 1, 1.7 (0.3 to 9), 2.5 (0.4 to 13.3) | 0, 0 (0 to 21.5), 0 (0 to 31.8) | 0, 0 (0 to 5.6), 0 (0 to 8.2) | 11, 5.5 (3.1 to 9.6), 8.1 (4.6 to 14.1) | 1, 1.4 (0.3 to 7.8), 2.1 (0.4 to 11.5) | 33, 5 (3.6 to 7), 7.4 (5.3 to 10.3) |
| **Rectus** | 10, 9.9 (5.5 to 17.3), 10.8 (6 to 18.9) | 0, 0 (0 to 7.7), 0 (0 to 8.4) | 0, 0 (0 to 4.1), 0 (0 to 4.5) | 0, 0 (0 to 21.5), 0 (0 to 23.6) | 0, 0 (0 to 6.1), 0 (0 to 6.7) | 0, 0 (0 to 21.5), 0 (0 to 23.6) | 0, 0 (0 to 5.6), 0 (0 to 6.1) | 5, 2.5 (1.1 to 5.7), 2.7 (1.2 to 6.3) | 0, 0 (0 to 5.3), 0 (0 to 5.8) | 15, 2.3 (1.4 to 3.7), 2.5 (1.5 to 4.1) |
| **Rostral** | 8, 7.9 (4.1 to 14.9), 24 (12.4 to 45.1) | 0, 0 (0 to 7.7), 0 (0 to 23.4) | 0, 0 (0 to 4.1), 0 (0 to 12.6) | 0, 0 (0 to 21.5), 0 (0 to 65.4) | 0, 0 (0 to 6.1), 0 (0 to 18.6) | 0, 0 (0 to 21.5), 0 (0 to 65.4) | 0, 0 (0 to 5.6), 0 (0 to 16.9) | 4, 2 (0.8 to 5), 6.1 (2.4 to 15.3) | 0, 0 (0 to 5.3), 0 (0 to 16) | 12, 1.8 (1 to 3.2), 5.5 (3.2 to 9.6) |
| **Precentral** | 0, 0 (0 to 3.7), 0 (0 to 1) | 9, 19.6 (10.7 to 33.2), 5.5 (3 to 9.2) | 3, 3.4 (1.2 to 9.4), 0.9 (0.3 to 2.6) | 0, 0 (0 to 21.5), 0 (0 to 6) | 0, 0 (0 to 6.1), 0 (0 to 1.7) | 0, 0 (0 to 21.5), 0 (0 to 6) | 0, 0 (0 to 5.6), 0 (0 to 1.6) | 9, 4.5 (2.4 to 8.3), 1.3 (0.7 to 2.3) | 5, 7.2 (3.1 to 15.9), 2 (0.9 to 4.4) | 26, 4 (2.7 to 5.7), 1.1 (0.8 to 1.6) |
| **Postcentral** | 0, 0 (0 to 3.7), 0 (0 to 1.8) | 6, 13 (6.1 to 25.7), 6.4 (3 to 12.6) | 3, 3.4 (1.2 to 9.4), 1.7 (0.6 to 4.6) | 0, 0 (0 to 21.5), 0 (0 to 10.6) | 0, 0 (0 to 6.1), 0 (0 to 3) | 0, 0 (0 to 21.5), 0 (0 to 10.6) | 0, 0 (0 to 5.6), 0 (0 to 2.7) | 11, 5.5 (3.1 to 9.6), 2.7 (1.5 to 4.7) | 6, 8.7 (4 to 17.7), 4.3 (2 to 8.7) | 26, 4 (2.7 to 5.7), 1.9 (1.3 to 2.8) |
| **Paracentral lobule** | 0, 0 (0 to 3.7), 0 (0 to 2.8) | 4, 8.7 (3.4 to 20.3), 6.8 (2.7 to 15.8) | 2, 2.2 (0.6 to 7.8), 1.7 (0.5 to 6.1) | 0, 0 (0 to 21.5), 0 (0 to 16.7) | 0, 0 (0 to 6.1), 0 (0 to 4.7) | 0, 0 (0 to 21.5), 0 (0 to 16.7) | 0, 0 (0 to 5.6), 0 (0 to 4.3) | 9, 4.5 (2.4 to 8.3), 3.5 (1.9 to 6.5) | 4, 5.8 (2.3 to 14), 4.5 (1.8 to 10.9) | 19, 2.9 (1.9 to 4.5), 2.2 (1.4 to 3.5) |
| **Subcentral** | 0, 0 (0 to 3.7), 0 (0 to 6.6) | 8, 17.4 (9.1 to 30.7), 31.4 (16.4 to 55.5) | 3, 3.4 (1.2 to 9.4), 6.1 (2.1 to 17.1) | 0, 0 (0 to 21.5), 0 (0 to 38.9) | 0, 0 (0 to 6.1), 0 (0 to 11) | 0, 0 (0 to 21.5), 0 (0 to 38.9) | 0, 0 (0 to 5.6), 0 (0 to 10.1) | 12, 6 (3.5 to 10.2), 10.8 (6.3 to 18.4) | 7, 10.1 (5 to 19.5), 18.3 (9 to 35.2) | 30, 4.6 (3.2 to 6.4), 8.3 (5.8 to 11.6) |
| **SPL** | 0, 0 (0 to 3.7), 0 (0 to 1.2) | 0, 0 (0 to 7.7), 0 (0 to 2.5) | 13, 14.6 (8.7 to 23.4), 4.8 (2.9 to 7.7) | 0, 0 (0 to 21.5), 0 (0 to 7.1) | 0, 0 (0 to 6.1), 0 (0 to 2) | 0, 0 (0 to 21.5), 0 (0 to 7.1) | 0, 0 (0 to 5.6), 0 (0 to 1.8) | 11, 5.5 (3.1 to 9.6), 1.8 (1 to 3.1) | 0, 0 (0 to 5.3), 0 (0 to 1.7) | 24, 3.7 (2.5 to 5.4), 1.2 (0.8 to 1.8) |
| **SMG** | 0, 0 (0 to 3.7), 0 (0 to 1.3) | 2, 4.3 (1.2 to 14.5), 1.5 (0.4 to 5.2) | 22, 24.7 (16.9 to 34.6), 8.8 (6 to 12.3) | 1, 7.1 (1.3 to 31.5), 2.5 (0.5 to 11.2) | 0, 0 (0 to 6.1), 0 (0 to 2.2) | 0, 0 (0 to 21.5), 0 (0 to 7.7) | 0, 0 (0 to 5.6), 0 (0 to 2) | 22, 11 (7.4 to 16.1), 3.9 (2.6 to 5.7) | 8, 11.6 (6 to 21.2), 4.1 (2.1 to 7.6) | 55, 8.4 (6.5 to 10.7), 3 (2.3 to 3.8) |
| **ANG** | 0, 0 (0 to 3.7), 0 (0 to 1.2) | 0, 0 (0 to 7.7), 0 (0 to 2.5) | 20, 22.5 (15 to 32.2), 7.2 (4.8 to 10.2) | 1, 7.1 (1.3 to 31.5), 2.3 (0.4 to 10) | 0, 0 (0 to 6.1), 0 (0 to 1.9) | 0, 0 (0 to 21.5), 0 (0 to 6.9) | 0, 0 (0 to 5.6), 0 (0 to 1.8) | 12, 6 (3.5 to 10.2), 1.9 (1.1 to 3.2) | 4, 5.8 (2.3 to 14), 1.8 (0.7 to 4.5) | 37, 5.6 (4.1 to 7.7), 1.8 (1.3 to 2.4) |
| **Precuneus** | 0, 0 (0 to 3.7), 0 (0 to 1.3) | 0, 0 (0 to 7.7), 0 (0 to 2.7) | 15, 16.9 (10.5 to 26), 6 (3.7 to 9.2) | 2, 14.3 (4 to 39.9), 5.1 (1.4 to 14.2) | 0, 0 (0 to 6.1), 0 (0 to 2.2) | 0, 0 (0 to 21.5), 0 (0 to 7.6) | 0, 0 (0 to 5.6), 0 (0 to 2) | 17, 8.5 (5.4 to 13.2), 3 (1.9 to 4.7) | 4, 5.8 (2.3 to 14), 2.1 (0.8 to 5) | 38, 5.8 (4.2 to 7.8), 2.1 (1.5 to 2.8) |
| **Cuneus** | 0, 0 (0 to 3.7), 0 (0 to 4) | 0, 0 (0 to 7.7), 0 (0 to 8.4) | 3, 3.4 (1.2 to 9.4), 3.7 (1.3 to 10.2) | 5, 35.7 (16.3 to 61.2), 38.7 (17.7 to 66.4) | 0, 0 (0 to 6.1), 0 (0 to 6.6) | 0, 0 (0 to 21.5), 0 (0 to 23.4) | 0, 0 (0 to 5.6), 0 (0 to 6.1) | 14, 7 (4.2 to 11.4), 7.6 (4.6 to 12.4) | 1, 1.4 (0.3 to 7.8), 1.6 (0.3 to 8.4) | 23, 3.5 (2.3 to 5.2), 3.8 (2.5 to 5.6) |

| | | | | | | | | | | |
|---|---|---|---|---|---|---|---|---|---|---|
| **O1** | 0, 0 (0 to 3.7), 0 (0 to 4.4) | 0, 0 (0 to 7.7), 0 (0 to 9.2) | 3, 3.4 (1.2 to 9.4), 4 (1.4 to 11.3) | 3, 21.4 (7.6 to 47.6), 25.6 (9.1 to 56.9) | 0, 0 (0 to 6.1), 0 (0 to 7.3) | 0, 0 (0 to 21.5), 0 (0 to 25.7) | 0, 0 (0 to 5.6), 0 (0 to 6.7) | 9, 4.5 (2.4 to 8.3), 5.4 (2.9 to 10) | 1, 1.4 (0.3 to 7.8), 1.7 (0.3 to 9.3) | 16, 2.4 (1.5 to 3.9), 2.9 (1.8 to 4.7) |
| **O2** | 0, 0 (0 to 3.7), 0 (0 to 2.5) | 0, 0 (0 to 7.7), 0 (0 to 5.3) | 2, 2.2 (0.6 to 7.8), 1.6 (0.4 to 5.4) | 5, 35.7 (16.3 to 61.2), 24.8 (11.3 to 42.5) | 0, 0 (0 to 6.1), 0 (0 to 4.2) | 0, 0 (0 to 21.5), 0 (0 to 14.9) | 0, 0 (0 to 5.6), 0 (0 to 3.9) | 16, 8 (5 to 12.6), 5.6 (3.5 to 8.7) | 2, 2.9 (0.8 to 10), 2 (0.6 to 6.9) | 25, 3.8 (2.6 to 5.6), 2.6 (1.8 to 3.9) |
| **O3** | 0, 0 (0 to 3.7), 0 (0 to 4.3) | 0, 0 (0 to 7.7), 0 (0 to 9.1) | 0, 0 (0 to 4.1), 0 (0 to 4.9) | 1, 7.1 (1.3 to 31.5), 8.4 (1.5 to 37.2) | 0, 0 (0 to 6.1), 0 (0 to 7.2) | 0, 0 (0 to 21.5), 0 (0 to 25.5) | 0, 0 (0 to 5.6), 0 (0 to 6.6) | 7, 3.5 (1.7 to 7), 4.1 (2 to 8.3) | 1, 1.4 (0.3 to 7.8), 1.7 (0.3 to 9.2) | 9, 1.4 (0.7 to 2.6), 1.6 (0.9 to 3.1) |
| **Occipital pole** | 0, 0 (0 to 3.7), 0 (0 to 3.5) | 0, 0 (0 to 7.7), 0 (0 to 7.4) | 0, 0 (0 to 4.1), 0 (0 to 4) | 0, 0 (0 to 21.5), 0 (0 to 20.7) | 0, 0 (0 to 6.1), 0 (0 to 5.9) | 0, 0 (0 to 21.5), 0 (0 to 20.7) | 0, 0 (0 to 5.6), 0 (0 to 5.4) | 2, 1 (0.3 to 3.6), 1 (0.3 to 3.4) | 0, 0 (0 to 5.3), 0 (0 to 5.1) | 2, 0.3 (0.1 to 1.1), 0.3 (0.1 to 1.1) |
| **Lingual** | 0, 0 (0 to 3.7), 0 (0 to 2) | 0, 0 (0 to 7.7), 0 (0 to 4.1) | 2, 2.2 (0.6 to 7.8), 1.2 (0.3 to 4.2) | 4, 28.6 (11.7 to 54.6), 15.4 (6.3 to 29.4) | 0, 0 (0 to 6.1), 0 (0 to 3.3) | 0, 0 (0 to 21.5), 0 (0 to 11.6) | 0, 0 (0 to 5.6), 0 (0 to 3) | 12, 6 (3.5 to 10.2), 3.2 (1.9 to 5.5) | 2, 2.9 (0.8 to 10), 1.6 (0.4 to 5.4) | 20, 3 (2 to 4.7), 1.6 (1.1 to 2.5) |
| **Fusiform** | 0, 0 (0 to 3.7), 0 (0 to 1.8) | 0, 0 (0 to 7.7), 0 (0 to 3.8) | 3, 3.4 (1.2 to 9.4), 1.7 (0.6 to 4.7) | 0, 0 (0 to 21.5), 0 (0 to 10.6) | 10, 16.9 (9.5 to 28.5), 8.3 (4.7 to 14) | 0, 0 (0 to 21.5), 0 (0 to 10.6) | 0, 0 (0 to 5.6), 0 (0 to 2.7) | 29, 14.5 (10.3 to 20), 7.1 (5.1 to 9.9) | 1, 1.4 (0.3 to 7.8), 0.7 (0.1 to 3.8) | 43, 6.5 (4.9 to 8.7), 3.2 (2.4 to 4.3) |
| **Temporal pole** | 5, 5 (2.1 to 11.1), 2.1 (0.9 to 4.8) | 0, 0 (0 to 7.7), 0 (0 to 3.3) | 2, 2.2 (0.6 to 7.8), 1 (0.3 to 3.4) | 0, 0 (0 to 21.5), 0 (0 to 9.3) | 12, 20.3 (12 to 32.3), 8.8 (5.2 to 13.9) | 0, 0 (0 to 21.5), 0 (0 to 9.3) | 0, 0 (0 to 5.6), 0 (0 to 2.4) | 24, 12 (8.2 to 17.2), 5.2 (3.5 to 7.4) | 2, 2.9 (0.8 to 10), 1.3 (0.3 to 4.3) | 45, 6.8 (5.2 to 9), 3 (2.2 to 3.9) |
| **T1** | 0, 0 (0 to 3.7), 0 (0 to 1.5) | 0, 0 (0 to 7.7), 0 (0 to 3.1) | 10, 11.2 (6.2 to 19.5), 4.5 (2.5 to 7.8) | 1, 7.1 (1.3 to 31.5), 2.9 (0.5 to 12.6) | 12, 20.3 (12 to 32.3), 8.2 (4.8 to 13) | 0, 0 (0 to 21.5), 0 (0 to 8.6) | 0, 0 (0 to 5.6), 0 (0 to 2.2) | 35, 17.5 (12.9 to 23.4), 7 (5.2 to 9.4) | 4, 5.8 (2.3 to 14), 2.3 (0.9 to 5.6) | 62, 9.4 (7.4 to 11.9), 3.8 (3 to 4.8) |
| **T2** | 0, 0 (0 to 3.7), 0 (0 to 1.7) | 0, 0 (0 to 7.7), 0 (0 to 3.6) | 11, 12.4 (7 to 20.8), 5.7 (3.2 to 9.6) | 1, 7.1 (1.3 to 31.5), 3.3 (0.6 to 14.5) | 17, 28.8 (18.8 to 41.4), 13.3 (8.7 to 19.1) | 0, 0 (0 to 21.5), 0 (0 to 9.9) | 0, 0 (0 to 5.6), 0 (0 to 2.6) | 30, 15 (10.7 to 20.6), 6.9 (4.9 to 9.5) | 3, 4.3 (1.5 to 12), 2 (0.7 to 5.5) | 62, 9.4 (7.4 to 11.9), 4.3 (3.4 to 5.5) |
| **T3** | 1, 1 (0.2 to 5.4), 2.2 (0.4 to 11.8) | 0, 0 (0 to 7.7), 0 (0 to 16.9) | 7, 7.9 (3.9 to 15.4), 17.2 (8.4 to 33.6) | 1, 7.1 (1.3 to 31.5), 15.6 (2.8 to 68.8) | 13, 22 (13.4 to 34.1), 48.2 (29.2 to 74.6) | 0, 0 (0 to 21.5), 0 (0 to 47.1) | 0, 0 (0 to 5.6), 0 (0 to 12.2) | 22, 11 (7.4 to 16.1), 24.1 (16.1 to 35.2) | 1, 1.4 (0.3 to 7.8), 3.2 (0.6 to 17) | 45, 6.8 (5.2 to 9), 15 (11.3 to 19.8) |
| **Planum temporale** | 0, 0 (0 to 3.7), 0 (0 to 13.1) | 0, 0 (0 to 7.7), 0 (0 to 27.5) | 1, 1.1 (0.2 to 6.1), 4 (0.7 to 21.7) | 0, 0 (0 to 21.5), 0 (0 to 76.7) | 0, 0 (0 to 6.1), 0 (0 to 21.8) | 0, 0 (0 to 21.5), 0 (0 to 76.7) | 0, 0 (0 to 5.6), 0 (0 to 19.9) | 6, 3 (1.4 to 6.4), 10.7 (4.9 to 22.8) | 0, 0 (0 to 5.3), 0 (0 to 18.8) | 7, 1.1 (0.5 to 2.2), 3.8 (1.8 to 7.8) |
| **Planum polare** | 6, 5.9 (2.8 to 12.4), 6.5 (3 to 13.6) | 0, 0 (0 to 7.7), 0 (0 to 8.5) | 0, 0 (0 to 4.1), 0 (0 to 4.5) | 0, 0 (0 to 21.5), 0 (0 to 23.6) | 6, 10.2 (4.7 to 20.5), 11.2 (5.2 to 22.5) | 0, 0 (0 to 21.5), 0 (0 to 23.6) | 0, 0 (0 to 5.6), 0 (0 to 6.1) | 9, 4.5 (2.4 to 8.3), 4.9 (2.6 to 9.1) | 0, 0 (0 to 5.3), 0 (0 to 5.8) | 21, 3.2 (2.1 to 4.8), 3.5 (2.3 to 5.3) |
| **Short insular** | 15, 14.9 (9.2 to 23.1), 10.5 (6.5 to 16.3) | 1, 2.2 (0.4 to 11.3), 1.5 (0.3 to 8) | 5, 5.6 (2.4 to 12.5), 4 (1.7 to 8.8) | 1, 7.1 (1.3 to 31.5), 5 (0.9 to 22.2) | 4, 6.8 (2.7 to 16.2), 4.8 (1.9 to 11.4) | 0, 0 (0 to 21.5), 0 (0 to 15.2) | 0, 0 (0 to 5.6), 0 (0 to 3.9) | 22, 11 (7.4 to 16.1), 7.8 (5.2 to 11.4) | 1, 1.4 (0.3 to 7.8), 1 (0.2 to 5.5) | 49, 7.5 (5.7 to 9.7), 5.3 (4 to 6.9) |

| | | | | | | | | | | |
|---|---|---|---|---|---|---|---|---|---|---|
| **Long insular** | 6, 5.9 (2.8 to 12.4), 7.1 (3.3 to 14.8) | 1, 2.2 (0.4 to 11.3), 2.6 (0.5 to 13.6) | 7, 7.9 (3.9 to 15.4), 9.4 (4.6 to 18.4) | 1, 7.1 (1.3 to 31.5), 8.6 (1.5 to 37.8) | 5, 8.5 (3.7 to 18.4), 10.2 (4.4 to 22) | 0, 0 (0 to 21.5), 0 (0 to 25.9) | 0, 0 (0 to 5.6), 0 (0 to 6.7) | 24, 12 (8.2 to 17.2), 14.4 (9.8 to 20.7) | 1, 1.4 (0.3 to 7.8), 1.7 (0.3 to 9.3) | 45, 6.8 (5.2 to 9), 8.2 (6.2 to 10.9) |
| **SCA** | 10, 9.9 (5.5 to 17.3), 45.6 (25.2 to 79.6) | 0, 0 (0 to 7.7), 0 (0 to 35.5) | 0, 0 (0 to 4.1), 0 (0 to 19.1) | 0, 0 (0 to 21.5), 0 (0 to 99.2) | 0, 0 (0 to 6.1), 0 (0 to 28.2) | 0, 0 (0 to 21.5), 0 (0 to 99.2) | 0, 0 (0 to 5.6), 0 (0 to 25.7) | 9, 4.5 (2.4 to 8.3), 20.7 (11 to 38.4) | 0, 0 (0 to 5.3), 0 (0 to 24.3) | 19, 2.9 (1.9 to 4.5), 13.3 (8.6 to 20.6) |
| **Cingulate anterior** | 20, 19.8 (13.2 to 28.6), 15 (10 to 21.7) | 2, 4.3 (1.2 to 14.5), 3.3 (0.9 to 11) | 0, 0 (0 to 4.1), 0 (0 to 3.1) | 0, 0 (0 to 21.5), 0 (0 to 16.3) | 0, 0 (0 to 6.1), 0 (0 to 4.6) | 0, 0 (0 to 21.5), 0 (0 to 16.3) | 0, 0 (0 to 5.6), 0 (0 to 4.2) | 15, 7.5 (4.6 to 12), 5.7 (3.5 to 9.1) | 1, 1.4 (0.3 to 7.8), 1.1 (0.2 to 5.9) | 38, 5.8 (4.2 to 7.8), 4.4 (3.2 to 5.9) |
| **Cingulate middle** | 4, 4 (1.6 to 9.7), 2.5 (1 to 6.2) | 10, 21.7 (12.3 to 35.6), 13.9 (7.8 to 22.7) | 2, 2.2 (0.6 to 7.8), 1.4 (0.4 to 5) | 0, 0 (0 to 21.5), 0 (0 to 13.8) | 0, 0 (0 to 6.1), 0 (0 to 3.9) | 0, 0 (0 to 21.5), 0 (0 to 13.8) | 0, 0 (0 to 5.6), 0 (0 to 3.6) | 11, 5.5 (3.1 to 9.6), 3.5 (2 to 6.1) | 1, 1.4 (0.3 to 7.8), 0.9 (0.2 to 5) | 28, 4.3 (3 to 6.1), 2.7 (1.9 to 3.9) |
| **Cingulate posterior** | 0, 0 (0 to 3.7), 0 (0 to 2) | 0, 0 (0 to 7.7), 0 (0 to 4.2) | 14, 15.7 (9.6 to 24.7), 8.5 (5.2 to 13.4) | 2, 14.3 (4 to 39.9), 7.8 (2.2 to 21.7) | 0, 0 (0 to 6.1), 0 (0 to 3.3) | 0, 0 (0 to 21.5), 0 (0 to 11.7) | 0, 0 (0 to 5.6), 0 (0 to 3) | 15, 7.5 (4.6 to 12), 4.1 (2.5 to 6.5) | 1, 1.4 (0.3 to 7.8), 0.8 (0.1 to 4.2) | 32, 4.9 (3.5 to 6.8), 2.6 (1.9 to 3.7) |
| **PHG** | 0, 0 (0 to 3.7), 0 (0 to 3.8) | 0, 0 (0 to 7.7), 0 (0 to 8.1) | 1, 1.1 (0.2 to 6.1), 1.2 (0.2 to 6.4) | 1, 7.1 (1.3 to 31.5), 7.5 (1.3 to 32.9) | 17, 28.8 (18.8 to 41.4), 30.1 (19.7 to 43.3) | 0, 0 (0 to 21.5), 0 (0 to 22.5) | 0, 0 (0 to 5.6), 0 (0 to 5.8) | 29, 14.5 (10.3 to 20), 15.2 (10.8 to 21) | 1, 1.4 (0.3 to 7.8), 1.5 (0.3 to 8.1) | 49, 7.5 (5.7 to 9.7), 7.8 (5.9 to 10.2) |
| **Hippocampus** | 0, 0 (0 to 3.7), 0 (0 to 4.9) | 0, 0 (0 to 7.7), 0 (0 to 10.3) | 1, 1.1 (0.2 to 6.1), 1.5 (0.3 to 8.2) | 0, 0 (0 to 21.5), 0 (0 to 28.9) | 13, 22 (13.4 to 34.1), 29.6 (17.9 to 45.8) | 0, 0 (0 to 21.5), 0 (0 to 28.9) | 0, 0 (0 to 5.6), 0 (0 to 7.5) | 49, 24.5 (19.1 to 30.9), 32.9 (25.6 to 41.5) | 0, 0 (0 to 5.3), 0 (0 to 7.1) | 63, 9.6 (7.6 to 12.1), 12.9 (10.2 to 16.2) |
| **Amygdala** | 0, 0 (0 to 3.7), 0 (0 to 12.8) | 0, 0 (0 to 7.7), 0 (0 to 26.9) | 0, 0 (0 to 4.1), 0 (0 to 14.4) | 0, 0 (0 to 21.5), 0 (0 to 75.2) | 12, 20.3 (12 to 32.3), 71 (42 to 112.6) | 1, 7.1 (1.3 to 31.5), 24.9 (4.4 to 109.9) | 0, 0 (0 to 5.6), 0 (0 to 19.5) | 37, 18.5 (13.7 to 24.5), 64.6 (47.9 to 85.4) | 0, 0 (0 to 5.3), 0 (0 to 18.4) | 50, 7.6 (5.8 to 9.9), 26.6 (20.3 to 34.5) |
| **Putamen** | 0, 0 (0 to 3.7), 0 (0 to 3.5) | 0, 0 (0 to 7.7), 0 (0 to 7.5) | 1, 1.1 (0.2 to 6.1), 1.1 (0.2 to 5.9) | 0, 0 (0 to 21.5), 0 (0 to 20.8) | 1, 1.7 (0.3 to 9), 1.6 (0.3 to 8.7) | 0, 0 (0 to 21.5), 0 (0 to 20.8) | 0, 0 (0 to 5.6), 0 (0 to 5.4) | 3, 1.5 (0.5 to 4.3), 1.4 (0.5 to 4.2) | 0, 0 (0 to 5.3), 0 (0 to 5.1) | 5, 0.8 (0.3 to 1.8), 0.7 (0.3 to 1.7) |
| **Caudate** | 0, 0 (0 to 3.7), 0 (0 to 5.1) | 0, 0 (0 to 7.7), 0 (0 to 10.8) | 0, 0 (0 to 4.1), 0 (0 to 5.8) | 0, 0 (0 to 21.5), 0 (0 to 30.2) | 1, 1.7 (0.3 to 9), 2.4 (0.4 to 12.6) | 0, 0 (0 to 21.5), 0 (0 to 30.2) | 0, 0 (0 to 5.6), 0 (0 to 7.8) | 4, 2 (0.8 to 5), 2.8 (1.1 to 7) | 0, 0 (0 to 5.3), 0 (0 to 7.4) | 5, 0.8 (0.3 to 1.8), 1.1 (0.5 to 2.5) |
| **Globus pallidum** | 0, 0 (0 to 3.7), 0 (0 to 13.1) | 0, 0 (0 to 7.7), 0 (0 to 27.5) | 1, 1.1 (0.2 to 6.1), 4 (0.7 to 21.7) | 0, 0 (0 to 21.5), 0 (0 to 76.8) | 1, 1.7 (0.3 to 9), 6 (1.1 to 32.1) | 0, 0 (0 to 21.5), 0 (0 to 76.8) | 0, 0 (0 to 5.6), 0 (0 to 19.9) | 4, 2 (0.8 to 5), 7.1 (2.8 to 17.9) | 0, 0 (0 to 5.3), 0 (0 to 18.8) | 6, 0.9 (0.4 to 2), 3.3 (1.5 to 7.1) |
| **Internal capsule** | 0, 0 (0 to 3.7), 0 (0 to 3.8) | 0, 0 (0 to 7.7), 0 (0 to 7.9) | 0, 0 (0 to 4.1), 0 (0 to 4.3) | 0, 0 (0 to 21.5), 0 (0 to 22.1) | 0, 0 (0 to 6.1), 0 (0 to 6.3) | 0, 0 (0 to 21.5), 0 (0 to 22.1) | 0, 0 (0 to 5.6), 0 (0 to 5.7) | 4, 2 (0.8 to 5), 2.1 (0.8 to 5.2) | 0, 0 (0 to 5.3), 0 (0 to 5.4) | 4, 0.6 (0.2 to 1.6), 0.6 (0.2 to 1.6) |
| **Hypothalamus** | 0, 0 (0 to 3.7), 0 (0 to 5) | 0, 0 (0 to 7.7), 0 (0 to 10.6) | 0, 0 (0 to 4.1), 0 (0 to 5.7) | 0, 0 (0 to 21.5), 0 (0 to 29.5) | 0, 0 (0 to 6.1), 0 (0 to 8.4) | 9, 64.3 (38.8 to 83.7), 88.2 (53.2 to 114.8) | 0, 0 (0 to 5.6), 0 (0 to 7.7) | 10, 5 (2.7 to 9), 6.9 (3.8 to 12.3) | 0, 0 (0 to 5.3), 0 (0 to 7.2) | 19, 2.9 (1.9 to 4.5), 4 (2.6 to 6.1) |

| | | | | | | | | | | |
|---|---|---|---|---|---|---|---|---|---|---|
| **Thalamus** | 0, 0 (0 to 3.7), 0 (0 to 2.7) | 2, 4.3 (1.2 to 14.5), 3.2 (0.9 to 10.8) | 0, 0 (0 to 4.1), 0 (0 to 3.1) | 0, 0 (0 to 21.5), 0 (0 to 16.1) | 2, 3.4 (0.9 to 11.5), 2.5 (0.7 to 8.6) | 5, 35.7 (16.3 to 61.2), 26.7 (12.2 to 45.7) | 0, 0 (0 to 5.6), 0 (0 to 4.2) | 31, 15.5 (11.1 to 21.2), 11.6 (8.3 to 15.8) | 1, 1.4 (0.3 to 7.8), 1.1 (0.2 to 5.8) | 41, 6.2 (4.6 to 8.4), 4.7 (3.5 to 6.2) |
| **Mesencephalon** | 0, 0 (0 to 3.7), 0 (0 to 4) | 0, 0 (0 to 7.7), 0 (0 to 8.4) | 0, 0 (0 to 4.1), 0 (0 to 4.5) | 0, 0 (0 to 21.5), 0 (0 to 23.4) | 0, 0 (0 to 6.1), 0 (0 to 6.6) | 3, 21.4 (7.6 to 47.6), 23.3 (8.2 to 51.7) | 1, 1.5 (0.3 to 8.2), 1.7 (0.3 to 8.9) | 12, 6 (3.5 to 10.2), 6.5 (3.8 to 11.1) | 2, 2.9 (0.8 to 10), 3.1 (0.9 to 10.8) | 18, 2.7 (1.7 to 4.3), 3 (1.9 to 4.7) |
| **Pons** | 0, 0 (0 to 3.7), 0 (0 to 2.6) | 0, 0 (0 to 7.7), 0 (0 to 5.5) | 0, 0 (0 to 4.1), 0 (0 to 3) | 0, 0 (0 to 21.5), 0 (0 to 15.4) | 0, 0 (0 to 6.1), 0 (0 to 4.4) | 0, 0 (0 to 21.5), 0 (0 to 15.4) | 18, 27.7 (18.3 to 39.6), 19.8 (13.1 to 28.3) | 3, 1.5 (0.5 to 4.3), 1.1 (0.4 to 3.1) | 1, 1.4 (0.3 to 7.8), 1 (0.2 to 5.5) | 22, 3.3 (2.2 to 5), 2.4 (1.6 to 3.6) |
| **Medulla oblongata** | 0, 0 (0 to 3.7), 0 (0 to 14.5) | 0, 0 (0 to 7.7), 0 (0 to 30.5) | 0, 0 (0 to 4.1), 0 (0 to 16.4) | 0, 0 (0 to 21.5), 0 (0 to 85.3) | 0, 0 (0 to 6.1), 0 (0 to 24.2) | 0, 0 (0 to 21.5), 0 (0 to 85.3) | 7, 10.8 (5.3 to 20.6), 42.7 (21.1 to 81.6) | 2, 1 (0.3 to 3.6), 4 (1.1 to 14.2) | 0, 0 (0 to 5.3), 0 (0 to 20.9) | 9, 1.4 (0.7 to 2.6), 5.4 (2.9 to 10.2) |
| **Central** | 0, 0 (0 to 3.7), 0 (0 to 82.3) | 0, 0 (0 to 7.7), 0 (0 to 173.2) | 0, 0 (0 to 4.1), 0 (0 to 93) | 0, 0 (0 to 21.5), 0 (0 to 483.9) | 0, 0 (0 to 6.1), 0 (0 to 137.4) | 0, 0 (0 to 21.5), 0 (0 to 483.9) | 8, 12.3 (6.4 to 22.5), 276.6 (143.2 to 504.6) | 3, 1.5 (0.5 to 4.3), 33.7 (11.5 to 97) | 0, 0 (0 to 5.3), 0 (0 to 118.5) | 11, 1.7 (0.9 to 3), 37.6 (21.1 to 66.8) |
| **Culmen** | 0, 0 (0 to 3.7), 0 (0 to 18.3) | 0, 0 (0 to 7.7), 0 (0 to 38.4) | 0, 0 (0 to 4.1), 0 (0 to 20.6) | 0, 0 (0 to 21.5), 0 (0 to 107.3) | 0, 0 (0 to 6.1), 0 (0 to 30.5) | 0, 0 (0 to 21.5), 0 (0 to 107.3) | 8, 12.3 (6.4 to 22.5), 61.3 (31.7 to 111.9) | 3, 1.5 (0.5 to 4.3), 7.5 (2.5 to 21.5) | 0, 0 (0 to 5.3), 0 (0 to 26.3) | 11, 1.7 (0.9 to 3), 8.3 (4.7 to 14.8) |
| **Declive** | 0, 0 (0 to 3.7), 0 (0 to 38.8) | 0, 0 (0 to 7.7), 0 (0 to 81.5) | 0, 0 (0 to 4.1), 0 (0 to 43.8) | 0, 0 (0 to 21.5), 0 (0 to 227.7) | 0, 0 (0 to 6.1), 0 (0 to 64.6) | 0, 0 (0 to 21.5), 0 (0 to 227.7) | 7, 10.8 (5.3 to 20.6), 113.9 (56.2 to 217.9) | 3, 1.5 (0.5 to 4.3), 15.9 (5.4 to 45.7) | 0, 0 (0 to 5.3), 0 (0 to 55.8) | 10, 1.5 (0.8 to 2.8), 16.1 (8.8 to 29.4) |
| **Folium** | 0, 0 (0 to 3.7), 0 (0 to 90.4) | 0, 0 (0 to 7.7), 0 (0 to 190.2) | 0, 0 (0 to 4.1), 0 (0 to 102.1) | 0, 0 (0 to 21.5), 0 (0 to 531.4) | 0, 0 (0 to 6.1), 0 (0 to 150.9) | 0, 0 (0 to 21.5), 0 (0 to 531.4) | 4, 6.2 (2.4 to 14.8), 151.9 (59.7 to 364.8) | 3, 1.5 (0.5 to 4.3), 37 (12.6 to 106.5) | 0, 0 (0 to 5.3), 0 (0 to 130.2) | 7, 1.1 (0.5 to 2.2), 26.3 (12.8 to 53.9) |
| **Tuber** | 0, 0 (0 to 3.7), 0 (0 to 103.1) | 0, 0 (0 to 7.7), 0 (0 to 216.9) | 0, 0 (0 to 4.1), 0 (0 to 116.4) | 0, 0 (0 to 21.5), 0 (0 to 605.9) | 0, 0 (0 to 6.1), 0 (0 to 172) | 0, 0 (0 to 21.5), 0 (0 to 605.9) | 4, 6.2 (2.4 to 14.8), 173.2 (68.1 to 416) | 3, 1.5 (0.5 to 4.3), 42.2 (14.4 to 121.5) | 0, 0 (0 to 5.3), 0 (0 to 148.4) | 7, 1.1 (0.5 to 2.2), 30 (14.6 to 61.4) |
| **Pyramid** | 0, 0 (0 to 3.7), 0 (0 to 94) | 0, 0 (0 to 7.7), 0 (0 to 197.7) | 0, 0 (0 to 4.1), 0 (0 to 106.1) | 0, 0 (0 to 21.5), 0 (0 to 552.3) | 0, 0 (0 to 6.1), 0 (0 to 156.8) | 0, 0 (0 to 21.5), 0 (0 to 552.3) | 4, 6.2 (2.4 to 14.8), 157.8 (62 to 379.2) | 3, 1.5 (0.5 to 4.3), 38.5 (13.1 to 110.7) | 0, 0 (0 to 5.3), 0 (0 to 135.3) | 7, 1.1 (0.5 to 2.2), 27.3 (13.3 to 56) |
| **Uvula** | 0, 0 (0 to 3.7), 0 (0 to 50.7) | 0, 0 (0 to 7.7), 0 (0 to 106.7) | 0, 0 (0 to 4.1), 0 (0 to 57.3) | 0, 0 (0 to 21.5), 0 (0 to 298.1) | 0, 0 (0 to 6.1), 0 (0 to 84.6) | 0, 0 (0 to 21.5), 0 (0 to 298.1) | 8, 12.3 (6.4 to 22.5), 170.4 (88.2 to 310.9) | 3, 1.5 (0.5 to 4.3), 20.8 (7.1 to 59.8) | 0, 0 (0 to 5.3), 0 (0 to 73) | 11, 1.7 (0.9 to 3), 23.2 (13 to 41.2) |
| **Nodule** | 0, 0 (0 to 3.7), 0 (0 to 186.7) | 0, 0 (0 to 7.7), 0 (0 to 392.8) | 0, 0 (0 to 4.1), 0 (0 to 210.8) | 0, 0 (0 to 21.5), 0 (0 to 1097.2) | 0, 0 (0 to 6.1), 0 (0 to 311.5) | 0, 0 (0 to 21.5), 0 (0 to 1097.2) | 14, 21.5 (13.3 to 33), 1097.6 (677.1 to 1679.9) | 3, 1.5 (0.5 to 4.3), 76.4 (26.1 to 220) | 0, 0 (0 to 5.3), 0 (0 to 268.7) | 17, 2.6 (1.6 to 4.1), 131.9 (82.6 to 209.2) |
| **Ala lobuli centralis** | 0, 0 (0 to 3.7), 0 (0 to 3.3) | 0, 0 (0 to 7.7), 0 (0 to 7) | 0, 0 (0 to 4.1), 0 (0 to 3.7) | 0, 0 (0 to 21.5), 0 (0 to 19.5) | 0, 0 (0 to 6.1), 0 (0 to 5.5) | 0, 0 (0 to 21.5), 0 (0 to 19.5) | 5, 7.7 (3.3 to 16.8), 7 (3 to 15.2) | 3, 1.5 (0.5 to 4.3), 1.4 (0.5 to 3.9) | 0, 0 (0 to 5.3), 0 (0 to 4.8) | 8, 1.2 (0.6 to 2.4), 1.1 (0.6 to 2.2) |

| | | | | | | | | | | |
|---|---|---|---|---|---|---|---|---|---|---|
| **AQL** | 0, 0 (0 to 3.7), 0 (0 to 2.3) | 0, 0 (0 to 7.7), 0 (0 to 4.8) | 0, 0 (0 to 4.1), 0 (0 to 2.6) | 0, 0 (0 to 21.5), 0 (0 to 13.3) | 0, 0 (0 to 6.1), 0 (0 to 3.8) | 0, 0 (0 to 21.5), 0 (0 to 13.3) | 5, 7.7 (3.3 to 16.8), 4.7 (2.1 to 10.3) | 4, 2 (0.8 to 5), 1.2 (0.5 to 3.1) | 1, 1.4 (0.3 to 7.8), 0.9 (0.2 to 4.8) | 10, 1.5 (0.8 to 2.8), 0.9 (0.5 to 1.7) |
| **PQL** | 0, 0 (0 to 3.7), 0 (0 to 3) | 0, 0 (0 to 7.7), 0 (0 to 6.4) | 0, 0 (0 to 4.1), 0 (0 to 3.4) | 0, 0 (0 to 21.5), 0 (0 to 17.9) | 0, 0 (0 to 6.1), 0 (0 to 5.1) | 0, 0 (0 to 21.5), 0 (0 to 17.9) | 3, 4.6 (1.6 to 12.7), 3.8 (1.3 to 10.6) | 4, 2 (0.8 to 5), 1.7 (0.6 to 4.2) | 0, 0 (0 to 5.3), 0 (0 to 4.4) | 7, 1.1 (0.5 to 2.2), 0.9 (0.4 to 1.8) |
| **SSL** | 0, 0 (0 to 3.7), 0 (0 to 2.8) | 0, 0 (0 to 7.7), 0 (0 to 5.9) | 0, 0 (0 to 4.1), 0 (0 to 3.2) | 0, 0 (0 to 21.5), 0 (0 to 16.5) | 0, 0 (0 to 6.1), 0 (0 to 4.7) | 0, 0 (0 to 21.5), 0 (0 to 16.5) | 5, 7.7 (3.3 to 16.8), 5.9 (2.6 to 12.9) | 3, 1.5 (0.5 to 4.3), 1.1 (0.4 to 3.3) | 1, 1.4 (0.3 to 7.8), 1.1 (0.2 to 6) | 9, 1.4 (0.7 to 2.6), 1.1 (0.6 to 2) |
| **ISL/gracile** | 0, 0 (0 to 3.7), 0 (0 to 1.1) | 0, 0 (0 to 7.7), 0 (0 to 2.4) | 0, 0 (0 to 4.1), 0 (0 to 1.3) | 0, 0 (0 to 21.5), 0 (0 to 6.7) | 0, 0 (0 to 6.1), 0 (0 to 1.9) | 0, 0 (0 to 21.5), 0 (0 to 6.7) | 7, 10.8 (5.3 to 20.6), 3.3 (1.7 to 6.4) | 3, 1.5 (0.5 to 4.3), 0.5 (0.2 to 1.3) | 0, 0 (0 to 5.3), 0 (0 to 1.6) | 10, 1.5 (0.8 to 2.8), 0.5 (0.3 to 0.9) |
| **Biventer** | 0, 0 (0 to 3.7), 0 (0 to 3.3) | 0, 0 (0 to 7.7), 0 (0 to 7) | 0, 0 (0 to 4.1), 0 (0 to 3.7) | 0, 0 (0 to 21.5), 0 (0 to 19.4) | 0, 0 (0 to 6.1), 0 (0 to 5.5) | 0, 0 (0 to 21.5), 0 (0 to 19.4) | 5, 7.7 (3.3 to 16.8), 6.9 (3 to 15.2) | 3, 1.5 (0.5 to 4.3), 1.4 (0.5 to 3.9) | 0, 0 (0 to 5.3), 0 (0 to 4.8) | 8, 1.2 (0.6 to 2.4), 1.1 (0.6 to 2.2) |
| **Tonsilla** | 0, 0 (0 to 3.7), 0 (0 to 6.7) | 0, 0 (0 to 7.7), 0 (0 to 14.1) | 0, 0 (0 to 4.1), 0 (0 to 7.6) | 0, 0 (0 to 21.5), 0 (0 to 39.4) | 0, 0 (0 to 6.1), 0 (0 to 11.2) | 0, 0 (0 to 21.5), 0 (0 to 39.4) | 2, 3.1 (0.8 to 10.5), 5.6 (1.6 to 19.3) | 3, 1.5 (0.5 to 4.3), 2.7 (0.9 to 7.9) | 0, 0 (0 to 5.3), 0 (0 to 9.7) | 5, 0.8 (0.3 to 1.8), 1.4 (0.6 to 3.2) |
| **Flocculus** | 0, 0 (0 to 3.7), 0 (0 to 46.6) | 0, 0 (0 to 7.7), 0 (0 to 97.9) | 0, 0 (0 to 4.1), 0 (0 to 52.6) | 0, 0 (0 to 21.5), 0 (0 to 273.6) | 0, 0 (0 to 6.1), 0 (0 to 77.7) | 0, 0 (0 to 21.5), 0 (0 to 273.6) | 3, 4.6 (1.6 to 12.7), 58.6 (20.1 to 161.6) | 3, 1.5 (0.5 to 4.3), 19.1 (6.5 to 54.9) | 0, 0 (0 to 5.3), 0 (0 to 67) | 6, 0.9 (0.4 to 2), 11.6 (5.3 to 25.1) |

**Supplemental Table 22. Radial cerebral anatomical patterns of neuroepithelial tumors, primary central nervous system lymphoma and brain metastases**

The involvement of the radial cerebral sectors by brain tumors in total, neuroepithelial tumors (NT), primary central nervous system lymphoma (PCNSL) or metastases is given in absolute numbers (n), relative tumor prevalence (RTP) and relative tumor density (RTD).

| | Total (n, % [95% CI]) | NT (n, % [95% CI]) | PCNSL (n, % [95% CI]) | Metastases (n, % [95% CI]) |
|---|---|---|---|---|
| **Total** | 815, 100 (99.5 to 100) | 540, 100 (99.3 to 100) | 30, 100 (88.6 to 100) | 245, 100 (98.5 to 100) |
| **Cortex** | 772, 94.7 (93 to 96.1) | 521, 96.5 (94.6 to 97.7) | 8, 26.7 (14.2 to 44.4) | 243, 99.2 (97.1 to 99.8) |
| **Cerebral subcortical WMS** | 780, 95.7 (94.1 to 96.9) | 509, 94.3 (92 to 95.9) | 28, 93.3 (78.7 to 98.2) | 243, 99.2 (97.1 to 99.8) |
| **Cerebral subgyral white matter** | 714, 87.6 (85.2 to 89.7) | 509, 94.3 (92 to 95.9) | 26, 86.7 (70.3 to 94.7) | 179, 73.1 (67.2 to 78.2) |
| **Cerebral gyral white matter** | 595, 73 (69.9 to 75.9) | 498, 92.2 (89.7 to 94.2) | 24, 80 (62.7 to 90.5) | 73, 29.8 (24.4 to 35.8) |
| **Cerebral lobar white matter** | 508, 62.3 (59 to 65.6) | 461, 85.4 (82.1 to 88.1) | 21, 70 (52.1 to 83.3) | 26, 10.6 (7.3 to 15.1) |
| **Internal capsule** | 19, 2.3 (1.5 to 3.6) | 4, 0.7 (0.3 to 1.9) | 14, 46.7 (30.2 to 63.9) | 1, 0.4 (0.1 to 2.3) |
| **LV ependyma** | 519, 63.7 (60.3 to 66.9) | 478, 88.5 (85.6 to 90.9) | 18, 60 (42.3 to 75.4) | 23, 9.4 (6.3 to 13.7) |
| **LV ependyma - diffuse** | 355, 43.6 (40.2 to 47) | 332, 61.5 (57.3 to 65.5) | 18, 60 (42.3 to 75.4) | 5, 2 (0.9 to 4.7) |
| **Corpus callosum - subependymal** | 100, 12.3 (10.2 to 14.7) | 89, 16.5 (13.6 to 19.8) | 11, 36.7 (21.9 to 54.5) | 0, 0 (0 to 1.5) |
| **Corpus callosum - central** | 44, 5.4 (4 to 7.2) | 24, 4.4 (3 to 6.5) | 17, 56.7 (39.2 to 72.6) | 3, 1.2 (0.4 to 3.5) |

**Supplemental Table 23. Radial cerebral anatomical patterns of neuroepithelial tumor subentities**

The involvement of the radial cerebral sectors by neuroepithelial tumors (NT) in total, glioblastoma (GBM), WHO grade III (gIIIG) and II (gIIG) glioma is given in absolute numbers (n), relative tumor prevalence (RTP) and relative tumor density (RTD).

| | Total (n, % [95% CI]) | GBM (n, % [95% CI]) | gIIIG (n, % [95% CI]) | gIIG (n, % [95% CI]) |
|---|---|---|---|---|
| **Total** | 510, 100 (99.3 to 100) | 368, 100 (99 to 100) | 99, 100 (96.3 to 100) | 43, 100 (91.8 to 100) |
| **Cortex** | 493, 96.7 (94.7 to 97.9) | 352, 95.7 (93.1 to 97.3) | 98, 99 (94.5 to 99.8) | 43, 100 (91.8 to 100) |
| **Cerebral subcortical WMS** | 487, 95.5 (93.3 to 97) | 351, 95.4 (92.7 to 97.1) | 96, 97 (91.5 to 99) | 40, 93 (81.4 to 97.6) |
| **Cerebral subgyral white matter** | 487, 95.5 (93.3 to 97) | 353, 95.9 (93.4 to 97.5) | 94, 94.9 (88.7 to 97.8) | 40, 93 (81.4 to 97.6) |
| **Cerebral gyral white matter** | 478, 93.7 (91.3 to 95.5) | 355, 96.5 (94.1 to 97.9) | 89, 89.9 (82.4 to 94.4) | 34, 79.1 (64.8 to 88.6) |
| **Cerebral lobar white matter** | 445, 87.3 (84.1 to 89.9) | 340, 92.4 (89.2 to 94.7) | 77, 77.8 (68.6 to 84.8) | 28, 65.1 (50.2 to 77.6) |
| **Internal capsule** | 4, 0.8 (0.3 to 2) | 4, 1.1 (0.4 to 2.8) | 0, 0 (0 to 3.7) | 0, 0 (0 to 8.2) |
| **LV ependyma** | 460, 90.2 (87.3 to 92.5) | 347, 94.3 (91.4 to 96.2) | 83, 83.8 (75.3 to 89.8) | 30, 69.8 (54.9 to 81.4) |
| **LV ependyma - diffuse** | 324, 63.5 (59.3 to 67.6) | 269, 73.1 (68.3 to 77.4) | 41, 41.4 (32.2 to 51.3) | 14, 32.6 (20.5 to 47.5) |
| **Corpus callosum - subependymal** | 89, 17.5 (14.4 to 21) | 73, 19.8 (16.1 to 24.2) | 11, 11.1 (6.3 to 18.8) | 5, 11.6 (5.1 to 24.5) |
| **Corpus callosum - central** | 24, 4.7 (3.2 to 6.9) | 23, 6.2 (4.2 to 9.2) | 1, 1 (0.2 to 5.5) | 0, 0 (0 to 8.2) |

**Supplemental Table 24. Radial cerebellar anatomical patterns of neuroepithelial tumors, primary central nervous system lymphoma and brain metastases**

The involvement of the radial cerebellar sectors by brain tumors in total, neuroepithelial tumors (NT), primary central nervous system lymphoma (PCNSL) or metastases is given in absolute numbers (n), relative tumor prevalence (RTP) and relative tumor density (RTD).

| | Total (n, % [95% CI]) | NT (n, % [95% CI]) | PCNSL (n, % [95% CI]) | Metastases (n, % [95% CI]) |
|---|---|---|---|---|
| **Total** | 136, 100 (97.3 to 100) | 43, 100 (91.8 to 100) | 5, 100 (56.6 to 100) | 88, 100 (95.8 to 100) |
| **Cerebellar cortex** | 133, 97.8 (93.7 to 99.2) | 43, 100 (91.8 to 100) | 3, 60 (23.1 to 88.2) | 87, 98.9 (93.8 to 99.8) |
| **Cerebellar subcortical WMS** | 131, 96.3 (91.7 to 98.4) | 40, 93 (81.4 to 97.6) | 5, 100 (56.6 to 100) | 86, 97.7 (92.1 to 99.4) |
| **Cerebellar sublobular white matter** | 116, 85.3 (78.4 to 90.3) | 40, 93 (81.4 to 97.6) | 5, 100 (56.6 to 100) | 71, 80.7 (71.2 to 87.6) |
| **Cerebellar lobular white matter** | 94, 69.1 (60.9 to 76.3) | 39, 90.7 (78.4 to 96.3) | 4, 80 (37.6 to 96.4) | 51, 58 (47.5 to 67.7) |
| **Cerebellar lobar white matter** | 45, 33.1 (25.7 to 41.4) | 37, 86 (72.7 to 93.4) | 4, 80 (37.6 to 96.4) | 4, 4.5 (1.8 to 11.1) |
| **Cerebellar nuclei** | 8, 5.9 (3 to 11.2) | 3, 7 (2.4 to 18.6) | 4, 80 (37.6 to 96.4) | 1, 1.1 (0.2 to 6.2) |
| **Cerebellar peduncles** | 13, 9.6 (5.7 to 15.7) | 8, 18.6 (9.7 to 32.6) | 4, 80 (37.6 to 96.4) | 1, 1.1 (0.2 to 6.2) |
| **4th ventricle** | 47, 34.6 (27.1 to 42.9) | 41, 95.3 (84.5 to 98.7) | 2, 40 (11.8 to 76.9) | 4, 4.5 (1.8 to 11.1) |

**Supplemental Table 25. Pre- and postoperative growth patterns of neuroepithelial tumors, primary central nervous system lymphoma and brain metastases**

The pre- and postoperative growth patterns of brain tumors in total, neuroepithelial tumors (NT), primary central nervous system lymphoma (PCNSL) or metastases is given in absolute (n) and relative (%) numbers.

| Growth pattern | Total (n, % [95% CI]) | NT (n, % [95% CI]) | PCNSL (n, % [95% CI]) | Metastases (n, % [95% CI]) |
|---|---|---|---|---|
| **Total** | 1000, 100 (99.6 to 100) | 657, 100 (99.4 to 100) | 44, 100 (92 to 100) | 299, 100 (98.7 to 100) |
| **PREOPERATIVE** | | | | |
| **spherical** | 40, 4 (3 to 5.4) | 9, 1.4 (0.7 to 2.6) | 0, 0 (0 to 8) | 31, 10.4 (7.4 to 14.3) |
| **ventriculofugal** | 73, 7.3 (5.8 to 9.1) | 72, 11 (8.8 to 13.6) | 1, 2.3 (0.4 to 11.8) | 0, 0 (0 to 1.3) |
| **ventriculopetal** | 5, 0.5 (0.2 to 1.2) | 5, 0.8 (0.3 to 1.8) | 0, 0 (0 to 8) | 0, 0 (0 to 1.3) |
| **along WM tracts** | 8, 0.8 (0.4 to 1.6) | 0, 0 (0 to 0.6) | 8, 18.2 (9.5 to 32) | 0, 0 (0 to 1.3) |
| **subependymal** | 34, 3.4 (2.4 to 4.7) | 32, 4.9 (3.5 to 6.8) | 2, 4.5 (1.3 to 15.1) | 0, 0 (0 to 1.3) |
| **leptomeningeal** | 6, 0.6 (0.3 to 1.3) | 6, 0.9 (0.4 to 2) | 0, 0 (0 to 8) | 0, 0 (0 to 1.3) |
| **CSF spread** | 1, 0.1 (0 to 0.6) | 0, 0 (0 to 0.6) | 1, 2.3 (0.4 to 11.8) | 0, 0 (0 to 1.3) |
| **new independent lesion** | 8, 0.8 (0.4 to 1.6) | 0, 0 (0 to 0.6) | 0, 0 (0 to 8) | 8, 2.7 (1.4 to 5.2) |
| **ND** | 72, 7.2 (5.8 to 9) | 37, 5.6 (4.1 to 7.7) | 1, 2.3 (0.4 to 11.8) | 34, 11.4 (8.3 to 15.5) |
| **POSTOPERATIVE** | | | | |
| **Total** | 1000, 100 (99.6 to 100) | 657, 100 (99.4 to 100) | 44, 100 (92 to 100) | 299, 100 (98.7 to 100) |
| **spherical** | 111, 11.1 (9.3 to 13.2) | 18, 2.7 (1.7 to 4.3) | 2, 4.5 (1.3 to 15.1) | 91, 30.4 (25.5 to 35.9) |
| **ventriculofugal** | 408, 40.8 (37.8 to 43.9) | 408, 62.1 (58.3 to 65.7) | 0, 0 (0 to 8) | 0, 0 (0 to 1.3) |
| **ventriculopetal** | 21, 2.1 (1.4 to 3.2) | 21, 3.2 (2.1 to 4.8) | 0, 0 (0 to 8) | 0, 0 (0 to 1.3) |
| **along WM tracts** | 13, 1.3 (0.8 to 2.2) | 1, 0.2 (0 to 0.9) | 12, 27.3 (16.3 to 41.8) | 0, 0 (0 to 1.3) |
| **subependymal** | 370, 37 (34.1 to 40) | 365, 55.6 (51.7 to 59.3) | 4, 9.1 (3.6 to 21.2) | 1, 0.3 (0.1 to 1.9) |
| **leptomeningeal** | 52, 5.2 (4 to 6.8) | 48, 7.3 (5.6 to 9.6) | 1, 2.3 (0.4 to 11.8) | 3, 1 (0.3 to 2.9) |
| **CSF spread** | 24, 2.4 (1.6 to 3.5) | 21, 3.2 (2.1 to 4.8) | 0, 0 (0 to 8) | 3, 1 (0.3 to 2.9) |
| **new independent lesion** | 116, 11.6 (9.8 to 13.7) | 0, 0 (0 to 0.6) | 0, 0 (0 to 8) | 116, 38.8 (33.4 to 44.4) |
| **ND** | 194, 19.4 (17.1 to 22) | 87, 13.2 (10.9 to 16) | 19, 43.2 (29.7 to 57.8) | 88, 29.4 (24.6 to 34.8) |

### Supplemental Table 26. Pre- and postoperative growth patterns of neuroepithelial tumor subentities

The pre- and postoperative growth patterns of neuroepithelial tumors (NT) in total, glioblastoma (GBM), WHO grade III (gIIIG) and II (gIIG) glioma, developmental tumors (DT), pilocytic astrocytoma (PA), ependymoma (EP), or medulloblastoma (MB) is given in absolute (n) and relative (%) numbers.

| Growth pattern | Total (n, % [95% CI]) | GBM (n, % [95% CI]) | gIIIG (n, % [95% CI]) | gIIG (n, % [95% CI]) | DT (n, % [95% CI]) | EP (n, % [95% CI]) | MB (n, % [95% CI]) | PA (n, % [95% CI]) |
|---|---|---|---|---|---|---|---|---|
| **Total** | 635, 100 (99.4 to 100) | 385, 100 (99 to 100) | 110, 100 (96.6 to 100) | 50, 100 (92.9 to 100) | 19, 100 (83.2 to 100) | 26, 100 (87.1 to 100) | 15, 100 (79.6 to 100) | 30, 100 (88.6 to 100) |
| **PREOPERATIVE** | | | | | | | | |
| **spherical** | 9, 1.4 (0.7 to 2.7) | 1, 0.3 (0 to 1.5) | 1, 0.9 (0.2 to 5) | 0, 0 (0 to 7.1) | 4, 21.1 (8.5 to 43.3) | 2, 7.7 (2.1 to 24.1) | 0, 0 (0 to 20.4) | 1, 3.3 (0.6 to 16.7) |
| **ventriculofugal** | 71, 11.2 (9 to 13.9) | 37, 9.6 (7.1 to 13) | 25, 22.7 (15.9 to 31.4) | 4, 8 (3.2 to 18.8) | 2, 10.5 (2.9 to 31.4) | 1, 3.8 (0.7 to 18.9) | 1, 6.7 (1.2 to 29.8) | 1, 3.3 (0.6 to 16.7) |
| **ventriculopetal** | 4, 0.6 (0.2 to 1.6) | 1, 0.3 (0 to 1.5) | 2, 1.8 (0.5 to 6.4) | 1, 2 (0.4 to 10.5) | 0, 0 (0 to 16.8) | 0, 0 (0 to 12.9) | 0, 0 (0 to 20.4) | 0, 0 (0 to 11.4) |
| **along WM tracts** | 0, 0 (0 to 0.6) | 0, 0 (0 to 1) | 0, 0 (0 to 3.4) | 0, 0 (0 to 7.1) | 0, 0 (0 to 16.8) | 0, 0 (0 to 12.9) | 0, 0 (0 to 20.4) | 0, 0 (0 to 11.4) |
| **subependymal** | 32, 5 (3.6 to 7) | 20, 5.2 (3.4 to 7.9) | 8, 7.3 (3.7 to 13.7) | 1, 2 (0.4 to 10.5) | 2, 10.5 (2.9 to 31.4) | 1, 3.8 (0.7 to 18.9) | 0, 0 (0 to 20.4) | 0, 0 (0 to 11.4) |
| **leptomeningeal** | 6, 0.9 (0.4 to 2) | 3, 0.8 (0.3 to 2.3) | 2, 1.8 (0.5 to 6.4) | 0, 0 (0 to 7.1) | 1, 5.3 (0.9 to 24.6) | 0, 0 (0 to 12.9) | 0, 0 (0 to 20.4) | 0, 0 (0 to 11.4) |
| **CSF spread** | 0, 0 (0 to 0.6) | 0, 0 (0 to 1) | 0, 0 (0 to 3.4) | 0, 0 (0 to 7.1) | 0, 0 (0 to 16.8) | 0, 0 (0 to 12.9) | 0, 0 (0 to 20.4) | 0, 0 (0 to 11.4) |
| **new independent lesion** | 0, 0 (0 to 0.6) | 0, 0 (0 to 1) | 0, 0 (0 to 3.4) | 0, 0 (0 to 7.1) | 0, 0 (0 to 16.8) | 0, 0 (0 to 12.9) | 0, 0 (0 to 20.4) | 0, 0 (0 to 11.4) |
| **ND** | 31, 4.9 (3.5 to 6.8) | 5, 1.3 (0.6 to 3) | 11, 10 (5.7 to 17) | 11, 22 (12.8 to 35.2) | 3, 15.8 (5.5 to 37.6) | 1, 3.8 (0.7 to 18.9) | 0, 0 (0 to 20.4) | 0, 0 (0 to 11.4) |
| **POSTOPERATIVE** | | | | | | | | |
| **spherical** | 15, 2.4 (1.4 to 3.9) | 1, 0.3 (0 to 1.5) | 3, 2.7 (0.9 to 7.7) | 2, 4 (1.1 to 13.5) | 3, 15.8 (5.5 to 37.6) | 2, 7.7 (2.1 to 24.1) | 1, 6.7 (1.2 to 29.8) | 3, 10 (3.5 to 25.6) |
| **ventriculofugal** | 403, 63.5 (59.6 to 67.1) | 267, 69.4 (64.6 to 73.7) | 84, 76.4 (67.6 to 83.3) | 36, 72 (58.3 to 82.5) | 2, 10.5 (2.9 to 31.4) | 7, 26.9 (13.7 to 46.1) | 2, 13.3 (3.7 to 37.9) | 5, 16.7 (7.3 to 33.6) |
| **ventriculopetal** | 19, 3 (1.9 to 4.6) | 8, 2.1 (1.1 to 4) | 5, 4.5 (2 to 10.2) | 5, 10 (4.3 to 21.4) | 0, 0 (0 to 16.8) | 1, 3.8 (0.7 to 18.9) | 0, 0 (0 to 20.4) | 0, 0 (0 to 11.4) |
| **along WM tracts** | 1, 0.2 (0 to 0.9) | 1, 0.3 (0 to 1.5) | 0, 0 (0 to 3.4) | 0, 0 (0 to 7.1) | 0, 0 (0 to 16.8) | 0, 0 (0 to 12.9) | 0, 0 (0 to 20.4) | 0, 0 (0 to 11.4) |
| **subependymal** | 362, 57 (53.1 to 60.8) | 246, 63.9 (59 to 68.5) | 78, 70.9 (61.8 to 78.6) | 29, 58 (44.2 to 70.6) | 2, 10.5 (2.9 to 31.4) | 6, 23.1 (11 to 42.1) | 0, 0 (0 to 20.4) | 1, 3.3 (0.6 to 16.7) |
| **leptomeningeal** | 47, 7.4 (5.6 to 9.7) | 37, 9.6 (7.1 to 13) | 7, 6.4 (3.1 to 12.6) | 0, 0 (0 to 7.1) | 0, 0 (0 to 16.8) | 2, 7.7 (2.1 to 24.1) | 0, 0 (0 to 20.4) | 1, 3.3 (0.6 to 16.7) |
| **CSF spread** | 19, 3 (1.9 to 4.6) | 8, 2.1 (1.1 to 4) | 3, 2.7 (0.9 to 7.7) | 1, 2 (0.4 to 10.5) | 0, 0 (0 to 16.8) | 3, 11.5 (4 to 29) | 4, 26.7 (10.9 to 52) | 0, 0 (0 to 11.4) |

| | | | | | | | | |
|---|---|---|---|---|---|---|---|---|
| **new independent lesion** | 0, 0 (0 to 0.6) | 0, 0 (0 to 1) | 0, 0 (0 to 3.4) | 0, 0 (0 to 7.1) | 0, 0 (0 to 16.8) | 0, 0 (0 to 12.9) | 0, 0 (0 to 20.4) | 0, 0 (0 to 11.4) |
| **ND** | 79, 12.4 (10.1 to 15.2) | 22, 5.7 (3.8 to 8.5) | 12, 10.9 (6.4 to 18.1) | 7, 14 (7 to 26.2) | 12, 63.2 (41 to 80.9) | 10, 38.5 (22.4 to 57.5) | 5, 33.3 (15.2 to 58.3) | 11, 36.7 (21.9 to 54.5) |

**Supplemental Table 27. Pre- and postoperative growth patterns of metastases subentities**

The pre- and postoperative growth patterns of metastases in total and by organ of origin is given in in absolute (n) and relative (%) numbers. *GIT = gastrointestinal tract; UGT = urogenital tract.*

| Growth pattern | Total (n, % [95% CI]) | Lung (n, % [95% CI]) | Skin (n, % [95% CI]) | GIT (n, % [95% CI]) | Breast (n, % [95% CI]) | UGT (n, % [95% CI]) | Miscellaneous (n, % [95% CI]) |
|---|---|---|---|---|---|---|---|
| **Total** | 299, 100 (98.7 to 100) | 143, 100 (97.4 to 100) | 49, 100 (92.7 to 100) | 37, 100 (90.6 to 100) | 28, 100 (87.9 to 100) | 24, 100 (86.2 to 100) | 18, 100 (82.4 to 100) |
| **PREOPERATIVE** | | | | | | | |
| **spherical** | 31, 10.4 (7.4 to 14.3) | 12, 8.4 (4.9 to 14.1) | 9, 18.4 (10 to 31.4) | 4, 10.8 (4.3 to 24.7) | 2, 7.1 (2 to 22.6) | 3, 12.5 (4.3 to 31) | 1, 5.6 (1 to 25.8) |
| **ventriculofugal** | 0, 0 (0 to 1.3) | 0, 0 (0 to 2.6) | 0, 0 (0 to 7.3) | 0, 0 (0 to 9.4) | 0, 0 (0 to 12.1) | 0, 0 (0 to 13.8) | 0, 0 (0 to 17.6) |
| **ventriculopetal** | 0, 0 (0 to 1.3) | 0, 0 (0 to 2.6) | 0, 0 (0 to 7.3) | 0, 0 (0 to 9.4) | 0, 0 (0 to 12.1) | 0, 0 (0 to 13.8) | 0, 0 (0 to 17.6) |
| **along WM tracts** | 0, 0 (0 to 1.3) | 0, 0 (0 to 2.6) | 0, 0 (0 to 7.3) | 0, 0 (0 to 9.4) | 0, 0 (0 to 12.1) | 0, 0 (0 to 13.8) | 0, 0 (0 to 17.6) |
| **subependymal** | 0, 0 (0 to 1.3) | 0, 0 (0 to 2.6) | 0, 0 (0 to 7.3) | 0, 0 (0 to 9.4) | 0, 0 (0 to 12.1) | 0, 0 (0 to 13.8) | 0, 0 (0 to 17.6) |
| **leptomeningeal** | 0, 0 (0 to 1.3) | 0, 0 (0 to 2.6) | 0, 0 (0 to 7.3) | 0, 0 (0 to 9.4) | 0, 0 (0 to 12.1) | 0, 0 (0 to 13.8) | 0, 0 (0 to 17.6) |
| **CSF spread** | 0, 0 (0 to 1.3) | 0, 0 (0 to 2.6) | 0, 0 (0 to 7.3) | 0, 0 (0 to 9.4) | 0, 0 (0 to 12.1) | 0, 0 (0 to 13.8) | 0, 0 (0 to 17.6) |
| **new independent lesion** | 8, 2.7 (1.4 to 5.2) | 3, 2.1 (0.7 to 6) | 3, 6.1 (2.1 to 16.5) | 0, 0 (0 to 9.4) | 1, 3.6 (0.6 to 17.7) | 1, 4.2 (0.7 to 20.2) | 0, 0 (0 to 17.6) |
| **ND** | 34, 11.4 (8.3 to 15.5) | 15, 10.5 (6.5 to 16.6) | 6, 12.2 (5.7 to 24.2) | 5, 13.5 (5.9 to 28) | 3, 10.7 (3.7 to 27.2) | 3, 12.5 (4.3 to 31) | 2, 11.1 (3.1 to 32.8) |
| **POSTOPERATIVE** | | | | | | | |
| **spherical** | 91, 30.4 (25.5 to 35.9) | 40, 28 (21.3 to 35.8) | 17, 34.7 (22.9 to 48.7) | 14, 37.8 (24.1 to 53.9) | 11, 39.3 (23.6 to 57.6) | 5, 20.8 (9.2 to 40.5) | 4, 22.2 (9 to 45.2) |
| **ventriculofugal** | 0, 0 (0 to 1.3) | 0, 0 (0 to 2.6) | 0, 0 (0 to 7.3) | 0, 0 (0 to 9.4) | 0, 0 (0 to 12.1) | 0, 0 (0 to 13.8) | 0, 0 (0 to 17.6) |
| **ventriculopetal** | 0, 0 (0 to 1.3) | 0, 0 (0 to 2.6) | 0, 0 (0 to 7.3) | 0, 0 (0 to 9.4) | 0, 0 (0 to 12.1) | 0, 0 (0 to 13.8) | 0, 0 (0 to 17.6) |
| **along WM tracts** | 0, 0 (0 to 1.3) | 0, 0 (0 to 2.6) | 0, 0 (0 to 7.3) | 0, 0 (0 to 9.4) | 0, 0 (0 to 12.1) | 0, 0 (0 to 13.8) | 0, 0 (0 to 17.6) |
| **subependymal** | 1, 0.3 (0.1 to 1.9) | 0, 0 (0 to 2.6) | 1, 2 (0.4 to 10.7) | 0, 0 (0 to 9.4) | 0, 0 (0 to 12.1) | 0, 0 (0 to 13.8) | 0, 0 (0 to 17.6) |
| **leptomeningeal** | 3, 1 (0.3 to 2.9) | 0, 0 (0 to 2.6) | 1, 2 (0.4 to 10.7) | 0, 0 (0 to 9.4) | 1, 3.6 (0.6 to 17.7) | 1, 4.2 (0.7 to 20.2) | 0, 0 (0 to 17.6) |
| **CSF spread** | 3, 1 (0.3 to 2.9) | 0, 0 (0 to 2.6) | 1, 2 (0.4 to 10.7) | 0, 0 (0 to 9.4) | 1, 3.6 (0.6 to 17.7) | 1, 4.2 (0.7 to 20.2) | 0, 0 (0 to 17.6) |
| **new independent lesion** | 116, 38.8 (33.4 to 44.4) | 55, 38.5 (30.9 to 46.6) | 22, 44.9 (31.9 to 58.7) | 12, 32.4 (19.6 to 48.5) | 9, 32.1 (17.9 to 50.7) | 10, 41.7 (24.5 to 61.2) | 8, 44.4 (24.6 to 66.3) |
| **ND** | 88, 29.4 (24.6 to 34.8) | 42, 29.4 (22.5 to 37.3) | 16, 32.7 (21.2 to 46.6) | 12, 32.4 (19.6 to 48.5) | 8, 28.6 (15.3 to 47.1) | 5, 20.8 (9.2 to 40.5) | 5, 27.8 (12.5 to 50.9) |

**Supplemental Table 28. Survival analysis stratified by general topographic tumor anatomy**

Evidence for a difference between the survival curves in neuroepithelial tumors and glioblastoma stratified by general topographic anatomy according to pairwise comparison with the log-rank test and Bonferoni-Holm correction for multiple testing. ****very strong evidence (p < 0.001), **strong evidence (p < 0.01), *evidence (p < 0.05), +weak evidence (p < 0.1).*

| | Frontal lobe | Central lobe | Parietal lobe | Occipital lobe | Temporal lobe | Insular lobe | Limbic lobe | Basal ganglia | Diencephalon | Brainstem |
|---|---|---|---|---|---|---|---|---|---|---|
| **NEUROEPITHELIAL TUMORS** | | | | | | | | | | |
| **Central lobe** | 0.34 | - | - | - | - | - | - | - | - | - |
| **Parietal lobe** | 0.032* | 0.42 | - | - | - | - | - | - | - | - |
| **Occipital lobe** | 0.0002*** | 0.054+ | 0.15 | - | - | - | - | - | - | - |
| **Temporal lobe** | 0.0017** | 0.21 | 0.73 | 0.2 | - | - | - | - | - | - |
| **Insular lobe** | 0.34 | 0.94 | 0.53 | 0.089+ | 0.3 | - | - | - | - | - |
| **Limbic lobe** | 0.087+ | 0.65 | 0.72 | 0.087+ | 0.34 | 0.73 | - | - | - | - |
| **Basal ganglia** | 0.12 | 0.19 | 0.34 | 0.72 | 0.31 | 0.34 | 0.34 | - | - | - |
| **Diencephalon** | 0.34 | 0.88 | 0.62 | 0.14 | 0.34 | 0.94 | 0.74 | 0.31 | - | - |
| **Brainstem** | 0.31 | 0.14 | 0.032* | 0.0015** | 0.0053** | 0.15 | 0.056+ | 0.15 | 0.15 | - |
| **Cerebellum** | 0.0004*** | <0.0001*** | <0.0001*** | <0.0001*** | <0.0001*** | 0.00015*** | <0.0001*** | 0.0014** | 0.0001*** | 0.08+ |
| **GLIOBLASTOMA** | | | | | | | | | | |
| **Central lobe** | 0.89 | - | - | - | - | - | - | - | - | - |
| **Parietal lobe** | 0.75 | 0.79 | - | - | - | - | - | - | - | - |
| **Occipital lobe** | 0.6 | 0.6 | 0.36 | - | - | - | - | - | - | - |
| **Temporal lobe** | 0.6 | 0.6 | 0.36 | 0.81 | - | - | - | - | - | - |
| **Insular lobe** | 0.31 | 0.36 | 0.31 | 0.6 | 0.6 | - | - | - | - | - |
| **Limbic lobe** | 0.31 | 0.36 | 0.3 | 0.72 | 0.57 | 0.81 | - | - | - | - |
| **Basal ganglia** | 0.3 | 0.3 | 0.3 | 0.31 | 0.3 | 0.54 | 0.36 | - | - | - |
| **Diencephalon** | 0.3 | 0.3 | 0.3 | 0.36 | 0.31 | 0.77 | 0.58 | 0.57 | - | - |
| **Brainstem** | 0.5 | 0.5 | 0.36 | 0.58 | 0.38 | 0.79 | 0.68 | 0.81 | 0.85 | - |
| **Cerebellum** | 0.31 | 0.3 | 0.3 | 0.36 | 0.3 | 0.52 | 0.36 | 0.72 | 0.54 | 0.79 |

### Supplemental Table 29. Survival analysis stratified by the ventricular tumor topography

Evidence for a difference between the survival curves in unisegmental neuroepithelial tumors and glioblastoma stratified by their ventricular contact according to pairwise comparison with the log-rank test and Bonferoni-Holm correction for multiple testing. ****very strong evidence (p < 0.001), **strong evidence (p < 0.01), *evidence (p < 0.05), +weak evidence (p < 0.1).*

| | Frontal horn | Body | Atrium | Occipital horn | Temporal horn | 3rd ventricle |
|---|---|---|---|---|---|---|
| **NEUROEPITHELIAL TUMORS** | | | | | | |
| **Body** | 0.066+ | - | - | - | - | - |
| **Atrium** | 0.016* | 0.85 | - | - | - | - |
| **Occipital horn** | 0.36 | 0.95 | 0.85 | - | - | - |
| **Temporal horn** | 0.008** | 0.7 | 0.85 | 0.74 | - | - |
| **3rd ventricle** | 0.12 | 0.022* | 0.013* | 0.066+ | 0.0061** | - |
| **4th ventricle** | <0.0001*** | <0.0001*** | <0.0001*** | <0.0001*** | <0.0001*** | 0.85 |
| **GLIOBLASTOMA** | | | | | | |
| **Body** | 0.78 | - | - | - | - | - |
| **Atrium** | 0.75 | 0.78 | - | - | - | - |
| **Occipital horn** | 0.78 | 0.78 | 1 | - | - | - |
| **Temporal horn** | 0.78 | 0.78 | 0.78 | 0.78 | - | - |
| **3rd ventricle** | 0.95 | 0.78 | 0.78 | 0.78 | 0.78 | - |
| **4th ventricle** | 0.043* | 0.1 | 0.016* | 0.036* | 0.017* | 0.5 |

**Supplemental Table 30. Survival analysis stratified by tumor ventriculosegmentality**

Evidence for a difference between the survival curves in neuroepithelial tumors and glioblastoma stratified by their ventriculosegmentality according to pairwise comparison with the log-rank test and Bonferoni-Holm correction for multiple testing. ****very strong evidence (p < 0.001), **strong evidence (p < 0.01), *evidence (p < 0.05), +weak evidence (p < 0.1).*

| | Unisegmental | Multisegmental |
|---|---|---|
| **NEUROEPITHELIAL TUMORS** | | |
| **Multisegmental** | <0.0001*** | - |
| **No contact to the ventricle** | 0.002** | <0.0001*** |
| **GLIOBLASTOMA** | | |
| **Multisegmental** | 0.0003*** | - |
| **No contact to the ventricle** | 0.19 | 0.0045** |

**Supplemental Table 31. Survival analysis stratified by the radial ventriculo-cortical tumor anatomy**

Evidence for a difference between the survival curves in neuroepithelial tumors and glioblastoma stratified by the involvement of the radial ventriculo-cortical sectors according to pairwise comparison with the log-rank test and Bonferoni-Holm correction for multiple testing. *WMS = white matter sector. ***very strong evidence (p < 0.001), **strong evidence (p < 0.01), *evidence (p < 0.05), +weak evidence (p < 0.1).*

| | Cortex | Cerebral subcortical WMS | Cerebral subgyral WMS | Cerebral gyral WMS |
|---|---|---|---|---|
| **NEUROEPITHELIAL TUMORS** | | | | |
| **Cerebral subcortical WMS** | 0.73 | - | - | - |
| **Cerebral subgyral WMS** | 0.73 | 1 | - | - |
| **Cerebral gyral WMS** | 0.57 | 0.72 | 0.72 | - |
| **Cerebral lobar WMS** | 0.28 | 0.28 | 0.28 | 0.57 |
| **GLIOBLASTOMA** | | | | |
| **Cerebral subcortical WMS** | 0.98 | - | - | - |
| **Cerebral subgyral WMS** | 0.98 | 0.98 | - | - |
| **Cerebral gyral WMS** | 0.98 | 0.98 | 0.98 | - |
| **Cerebral lobar WMS** | 0.98 | 0.98 | 0.98 | 0.98 |

**Supplemental Table 32. Survival analysis stratified by capsular, ependymal and corpus callosum tumor anatomy**

Evidence for a difference between the survival curves in neuroepithelial tumors and glioblastoma stratified by the invasion of the internal capsule, focal or diffuse involvement of the lateral ventricular (LV) ependyma and subependymal or central involvement of the corpus callosum (CC), according to pairwise comparison with the log-rank test and Bonferoni-Holm correction for multiple testing. ****very strong evidence (p < 0.001), **strong evidence (p < 0.01), *evidence (p < 0.05), +weak evidence (p < 0.1).*

| | Internal capsule | LV ependyma - focal | LV ependyma - diffuse | Corpus callosum - subependymal |
|---|---|---|---|---|
| **NEUROEPITHELIAL TUMORS** | | | | |
| **LV ependyma - focal** | <0.0001*** | - | - | - |
| **LV ependyma - diffuse** | 0.22 | <0.0001*** | - | - |
| **Corpus callosum - subependymal** | 0.51 | <0.0001*** | 0.22 | - |
| **Corpus callosum - central** | 0.7 | <0.0001*** | 0.014* | 0.22 |
| **GLIOBLASTOMA** | | | | |
| **LV ependyma - focal** | 0.0096** | - | - | - |
| **LV ependyma - diffuse** | 0.4 | 0.0003*** | - | - |
| **Corpus callosum - subependymal** | 0.85 | <0.0001*** | 0.096+ | - |
| **Corpus callosum - central** | 0.85 | 0.0002*** | 0.16 | 0.85 |

**Supplemental Table 33. Staging of lobar WHO grade II-III glioma and glioblastoma**

The staging of lobar glioblastoma (GBM), WHO grade III (gIIIG) and II (gIIG) glioma is given in absolute (n) and relative (%) numbers.

| | GBM (n, % [95% CI]) | gIIIG (n, % [95% CI]) | gIIG (n, % [95% CI]) |
|---|---|---|---|
| **Total** | 368, 100 (99 to 100) | 99, 100 (96.3 to 100) | 43, 100 (91.8 to 100) |
| **Stage I** | 96, 26.1 (21.9 to 30.8) | 58, 58.6 (48.7 to 67.8) | 28, 65.1 (50.2 to 77.6) |
| **Stage II** | 246, 66.8 (61.9 to 71.5) | 40, 40.4 (31.3 to 50.3) | 15, 34.9 (22.4 to 49.8) |
| **Stage III** | 26, 7.1 (4.9 to 10.2) | 1, 1 (0.2 to 5.5) | 0, 0 (0 to 8.2) |

### References


1. Louis DN, Ohgaki H, Wiestler OD, et al. The 2007 WHO Classification of Tumours of the Central Nervous System. Acta Neuropathol 2007;114(2):97–109.
2. Louis DN, Perry A, Reifenberger G, et al. The 2016 World Health Organization Classification of Tumors of the Central Nervous System: a summary. Acta Neuropathol 2016;131(6):803–20.
3. Akeret K, van Niftrik BCH, Sebök M, et al. Topographic volume-standardization atlas of the human brain. medRxiv 2021.
4. Puelles L, Harrison M, Paxinos G, Watson C. A developmental ontology for the mammalian brain based on the prosomeric model. Trends Neurosci 2013;36(10):570–8.
5. Mai JK, Paxinos G. The Human Nervous System. Elsevier; 2012.
6. Puelles L. Brain segmentation and forebrain development in amniotes. Brain Res Bull 2001;55(6):695–710.
7. de Graaf-Peters VB, Hadders-Algra M. Ontogeny of the human central nervous system: What is happening when? Early Hum Dev 2006;82(4):257–66.
8. Nieuwenhuys R, Voogd J, Van Huijzen C. The human central nervous system. Springer; 2008.
9. Nieuwenhuys R. Principles of Current Vertebrate Neuromorphology. Brain Behav Evol 2017;90(2):117–30.
10. Roland PE, Zilles K. Structural divisions and functional fields in the human cerebral cortex. Brain Res Rev 1998;26(2–3):87–105.
